# Evaluation of Stroke Risk Following COVID-19 mRNA Bivalent Vaccines Among U.S. Adults Aged ≥65 Years

**DOI:** 10.1101/2023.10.10.23296624

**Authors:** Yun Lu, Kathryn Matuska, Gita Nadimpalli, Yuxin Ma, Nathan Duma, Henry Zhang, Yiyun Chiang, Hai Lyu, Yoganand Chillarige, Jeffrey Kelman, Richard Forshee, Steven A. Anderson

**Affiliations:** Center for Biologics Evaluation and Research, Food and Drug Administration, Silver Spring, MD, USA; Acumen LLC, Burlingame, CA, USA; Centers for Medicare & Medicaid Services, Washington, DC, USA

**Keywords:** COVID-19 mRNA bivalent vaccine, booster, hemorrhagic stroke, non-hemorrhagic or ischemic stroke, transient ischemic attack, Medicare beneficiaries, high-dose or adjuvanted influenza vaccine

## Abstract

In January 2023, the United States Food and Drug Administration and the Centers for Disease Control and Prevention noted a safety concern for ischemic stroke in adults ≥65 years receiving the BNT162b2; WT/OMI BA.4/BA.5 COVID-19 bivalent vaccine. This self-controlled case series analysis evaluated stroke risk among Medicare fee-for-service beneficiaries aged ≥65 years receiving: 1) a Pfizer-BioNTech (BNT162b2; WT/OMI BA.4/BA.5) or Moderna (mRNA-1273.222) COVID-19 bivalent vaccine, 2) high-dose/adjuvanted influenza vaccines, and 3) concomitant COVID-19 bivalent vaccines and influenza vaccines, from August 31 to November 6, 2022.

The primary analysis did not find elevated stroke risk following COVID-19 bivalent vaccines. In the age subgroup analyses, only the ≥85 year age group had a risk of NHS (Incident Rate Ratio (IRR)=1.36, 95% CI 1.09 – 1.69 [1-21 days]) and NHS/TIA (IRR=1.28, 95% CI 1.08 – 1.52 [1-21 days]) with BNT162b2 Bivalent WT/OMI BA.4/BA.5. Among beneficiaries receiving a concomitant COVID-19 bivalent vaccine and a high-dose/adjuvanted influenza vaccine, an increased risk was observed for NHS (IRR=1.20, 95% CI 1.01 – 1.42 [22-42 days]) with BNT162b2 Bivalent WT/OMI BA.4/BA.5 and for TIA (IRR=1.35, 95% CI 1.06 – 1.74 [1-21 days]) with mRNA-1273.222.

Results of the secondary analyses showed a small increased risk of NHS following high-dose or adjuvanted influenza vaccines (IRR=1.09, 95% CI 1.02 – 1.17 [22-42 days]).

## INTRODUCTION

On August 31, 2022, the United States (U.S.) Food and Drug Administration (FDA) authorized both the Pfizer-BioNTech (BNT162b2; WT/OMI BA.4/BA.5) and Moderna (mRNA-1273.222) coronavirus disease 2019 (COVID-19) bivalent vaccines for use as boosters through emergency use authorization (EUA).^1^ The COVID-19 bivalent vaccines include an mRNA component of the original COVID-19 strain as well as an mRNA component from the BA.4 and BA.5 lineages of the omicron variant.^1^ Both COVID-19 bivalent mRNA vaccines are recommended for individuals aged 6 months and older.^2^

On January 13, 2023, the U.S. FDA and the Centers for Disease Control and Prevention (CDC) issued a public communication regarding the identification of a preliminary safety signal for ischemic stroke among persons aged ≥65 years in the 1-21 days after receipt of the BNT162b2; WT/OMI BA.4/BA.5, in the Vaccine Safety Datalink (VSD) near real-time surveillance system.^3^ The VSD study also noted the risk of stroke in individuals who received both a BNT162b2; WT/OMI BA.4/BA.5 and a high-dose or adjuvanted influenza vaccine on the same day was higher than the risk of stroke following receipt of BNT162b2; WT/OMI BA.4/BA.5 alone.^3^

This manuscript summarizes the results of a self-controlled case series (SCCS) analysis conducted in the Medicare fee-for-service (FFS) population aged ≥65 years. Our objective was to estimate the risk of incident stroke following COVID-19 bivalent vaccines among Medicare beneficiaries. We also sought to investigate stroke risk among age groups, following influenza vaccines, and following concomitant influenza and COVID-19 bivalent vaccine administration.

## METHODS

### Data Sources

Medicare is a federally funded health insurance program that covers individuals aged ≥65 and those under 65 years with a disability or end-stage renal disease in the U.S. We utilized Medicare administrative files, which provide comprehensive data on enrollment, inpatient claims (Part A), and hospital outpatient and physician office visit claims (Part B). Beneficiaries’ demographic and enrollment information was obtained from the Centers for Medicare and Medicaid Services (CMS) Enrollment Database and the Common Medicare Environment. Information on nursing home residency status was ascertained using Medicare Skilled Nursing Facility claims and the Minimum Data Set (MDS 3.0).^4^

### Study Population, Study Period, and Study Design

The primary and secondary study populations consisted of beneficiaries aged ≥65 years who received a COVID-19 bivalent or a high-dose/adjuvanted influenza vaccine, respectively, had a stroke outcome during follow-up, and met the specified inclusion/exclusion criteria (eTable 1). Continuous FFS enrollment was required from 365 days before the relevant vaccination date. The study start date, August 31, 2022, was the EUA date for the COVID-19 bivalent vaccines. The study end date for each outcome was determined independently to ensure at least 90 percent data completeness.^3^ The specific end dates for each outcome can be found in eTable 2.

We conducted an SCCS analysis to compare the incidence of stroke outcomes following COVID-19 bivalent and high-dose/adjuvanted influenza vaccine administration within hypothesized risk intervals (1-21 days and 22-42 days) to a control interval (43-90 days). The determination of the risk intervals was based on biological plausibility, prior studies, and input from subject matter experts.^5,6^ SCCS studies leverage exposed (risk) and unexposed (control) periods within the same individual, inherently adjusting for sources of time-invariant confounding in between-individual comparisons.^7,8^ The study plan is outlined in eTable 1.

### Exposures, Outcomes and Follow-Up Time

The primary and secondary exposures of interest were the receipt of any COVID-19 bivalent and high-dose/adjuvanted influenza vaccines, respectively. Exposures were identified through Current Procedural Terminology (CPT®)/Healthcare Common Procedure Coding System (HCPCS) codes in various care settings (eTable 3, eTable 4).

The stroke outcomes were non-hemorrhagic stroke (NHS), transient ischemic attack (TIA), a combined outcome of non-hemorrhagic stroke and/or TIA (NHS/TIA), and hemorrhagic stroke (HS). Persons who had both NHS and TIA contributed only their first event to the combined NHS/TIA outcome. Incident stroke outcomes were defined as the first recorded stroke for an individual during the observation period following the exposure, with no previous outcome identified during a predefined 365-day clean window (eTable 4). Additionally, outcome-specific exclusion criteria, such as trauma codes (eTable 5) were applied to eliminate stroke cases determined to have causes other than COVID-19 vaccination.^3^ For the primary analysis of the COVID-19 bivalent vaccine, patients diagnosed with COVID-19 within 30 days prior to the outcome were excluded.^9–11^ Stroke outcomes were identified *using International Classification of Diseases, Tenth Revision, Clinical Modification* (ICD-10-CM) codes (eTable 4). All outcomes were captured in inpatient (IP) care settings, and TIA cases were additionally captured in the outpatient emergency department setting (OP-ED).

To ensure adequate follow-up, beneficiaries were required to accumulate time during both the risk and control intervals, except in cases where death occurred before the control window (eTable 6). For both the primary and secondary analysis, beneficiaries with the relevant vaccine exposure (COVID-19 bivalent or high-dose/adjuvanted influenza vaccine, respectively) were followed until the earliest occurrence of the follow-up period end, study period end, disenrollment, administration of a subsequent exposure vaccine, or death.

### Medical Record Review

To validate the claims-based outcome definitions, we retrieved and adjudicated a random sample of NHS medical records from the inpatient care settings. Using predetermined clinical definitions, cases were identified as true cases, non-cases, or potentially indeterminate.^12^ We calculated the positive predictive value (PPV) along with corresponding 95% confidence intervals (CIs) and conducted a PPV adjusted analysis to evaluate the presence and magnitude of bias in the risk estimates resulting from potential misclassification of NHS.^13^ eTable 7 summarizes results of medical record review to verify NHS outcomes from the COVID-19 bivalent study. We collected a sample of 87 NHS cases. The PPV was 80.46% (95% CI: 70.92 – 87.43%), indicating accurate identification of NHS cases.

### Statistical Analysis

Descriptive statistics were obtained for beneficiaries’ demographics, socio-economic status, medical history, prior COVID-19 or influenza diagnosis and concomitant vaccination. Both primary and secondary analyses used an SCCS study design with a post-vaccination control interval. We used conditional Poisson regression to estimate the incidence rate ratio (IRR) comparing rates in the risk and control intervals for each outcome.^8^ The Attributable Risk (AR) representing the excess risk of the outcome attributable to the exposure was calculated by dividing the excess number of cases by the number of eligible vaccinations or person-time.^14^

Given the high case fatality rate (CFR) associated with stroke events and to address potential bias arising from event occurrence or observation time being dependent on outcome occurrence, we used the Farrington adjustment, which introduced an additional term to account for the curtailed censoring process in all SCCS analyses (eTable 8).^15^ We conducted sensitivity analyses including: (i) incorporating seasonal patterns and changes in stroke incidence rates to account for potential time-varying confounding, and (ii) utilizing the PPV derived from MRR to minimize the NHS outcome misclassification.

We conducted two subgroup analyses for both the primary and secondary studies to evaluate the risk of each stroke outcome (a) by age group (65-74, 75-84, ≥85) and (b) for concomitant and non-concomitant COVID-19 bivalent and high-dose/adjuvanted influenza vaccination.

We additionally conducted a temporal scan to identify clusters of increased stroke risk within the 1-90 day period following COVID-19 bivalent and influenza vaccination. The maximum cluster size was set to 50% of the observation period (45 days), and the minimum window length to three days. The temporal scan was conducted using SaTScan.^16^

All analyses were conducted using R 4.0.3 (R Foundation for Statistical Computing, Vienna, Austria) and SAS v. 9.4 (SAS Institute Inc., Cary, NC, United States).

## RESULTS

### Primary Bivalent Vaccine Analyses

#### Descriptive Results

The primary analysis evaluated risk of the stroke outcomes following a COVID-19 bivalent vaccine. The study population included 5,397,278 beneficiaries who received either the BNT162b2; WT/OMI BA.4/BA.5 or mRNA-1273.222 vaccine. There were 2,886 cases of NHS, 2,641 cases of TIA, 4,788 cases of NHS/TIA, and 808 cases of HS across both brands. Table 1 provides a summary of the baseline characteristics of the eligible vaccinated Medicare FFS beneficiaries who experienced an outcome. Population characteristics were largely consistent between the COVID-19 bivalent vaccine brands. HS and NHS cases exhibited a high CFR (17-34%), with a high proportion of cases dying before accruing control time (5-16%) (eTable 6, eTable 8). All outcomes exhibited distinct historical seasonality patterns (eFigure 1). Approximately 10-15% of the population had a COVID-19 diagnosis claim in the 31 to 365 days prior to stroke outcomes, and 34-45% of the population had a concomitant high-dose or adjuvanted influenza vaccination (Table 1). Patient characteristics stratified by age subgroup and concomitant influenza vaccine status are shown in eTables 9-12.

**Table 1.**
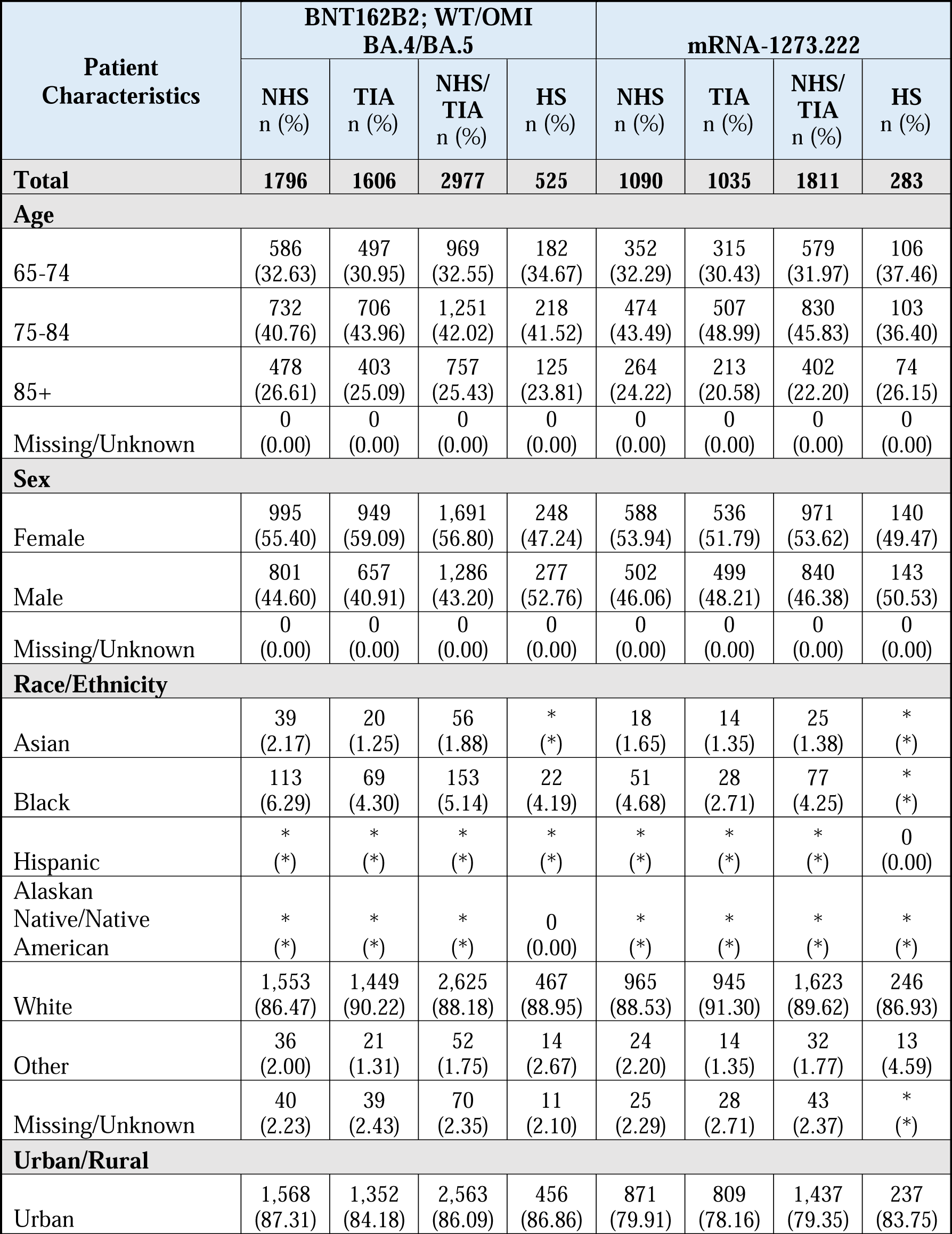
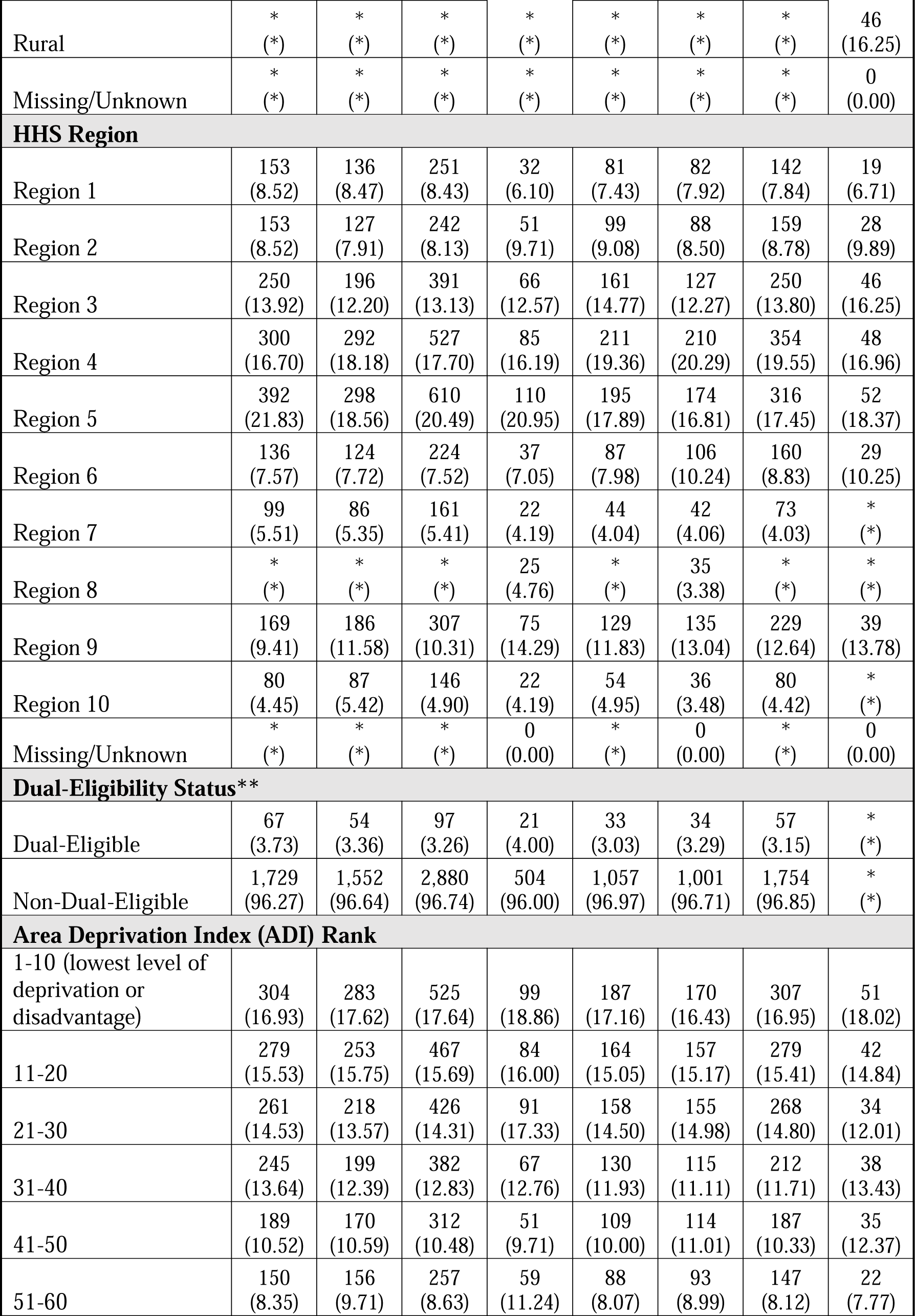
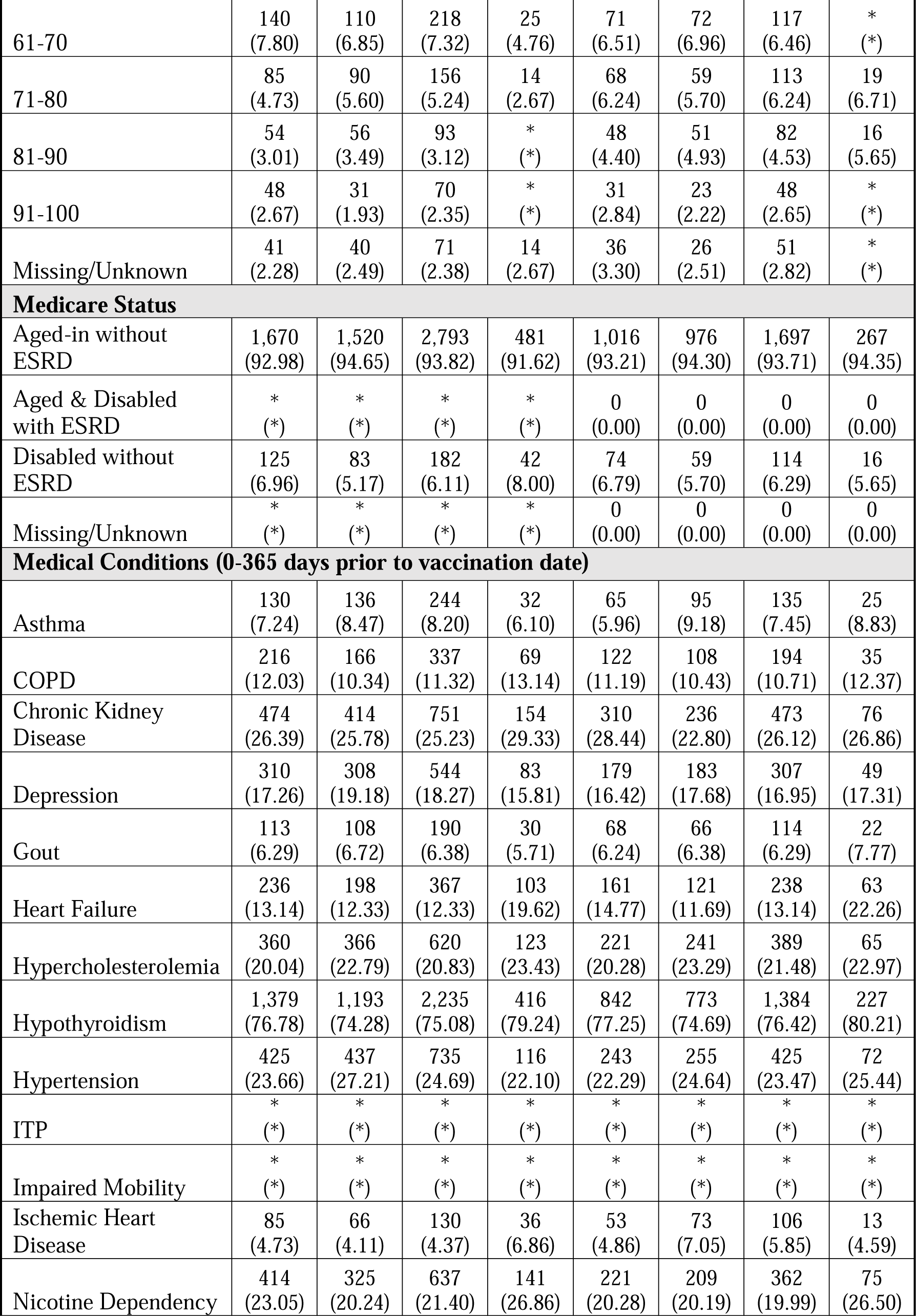
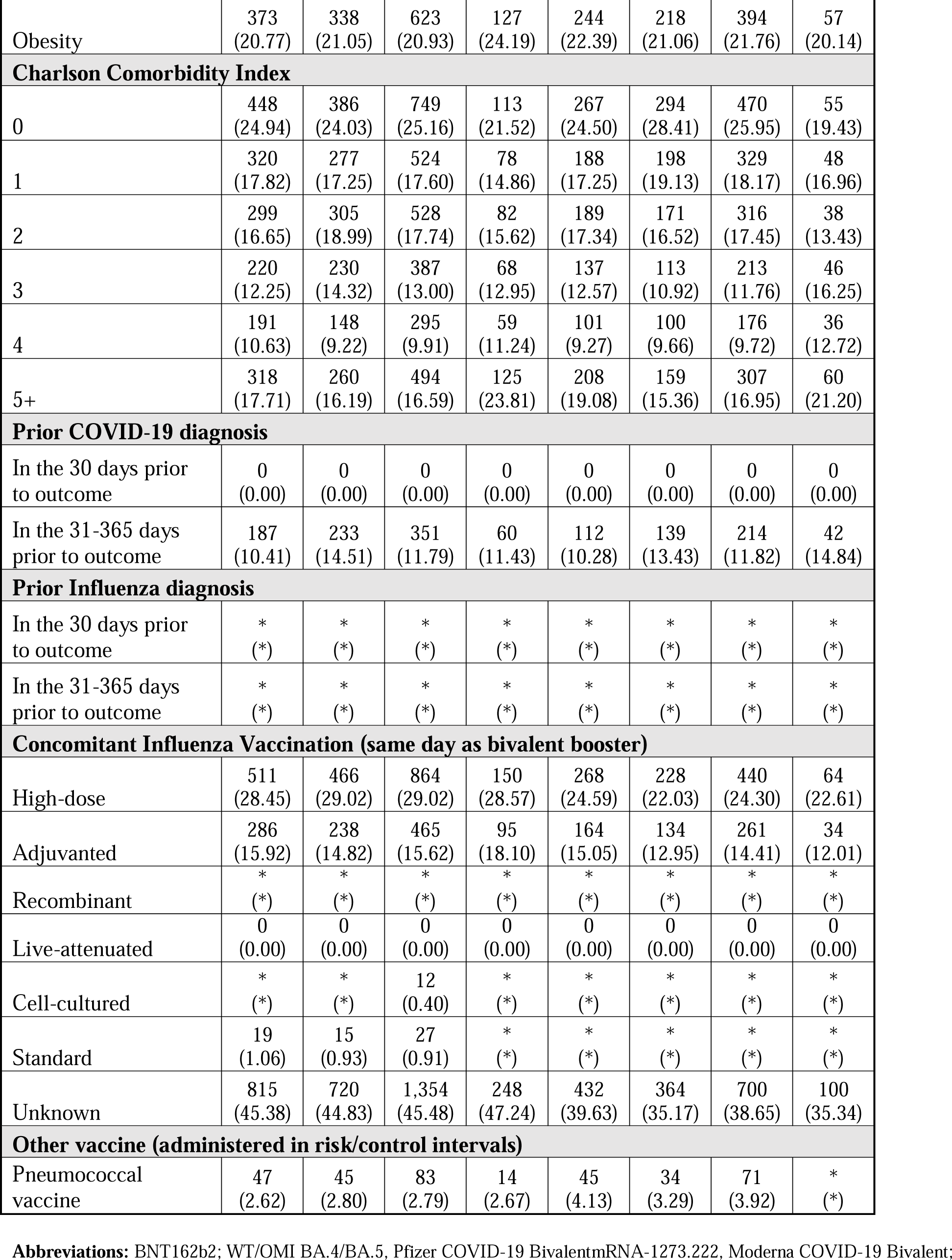
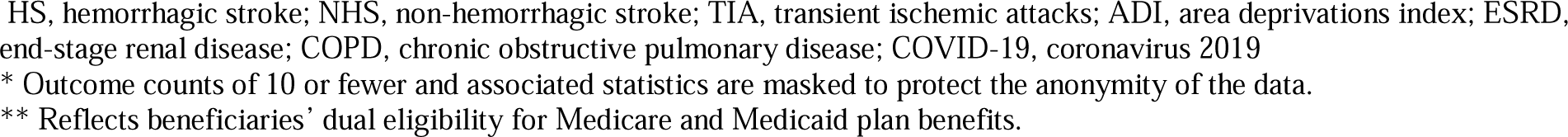
Summary of Demographics, Socio-Economic Status, Residence, Health Status, and Healthcare Utilization of COVID-19 Bivalent Vaccinated Medicare Beneficiaries with Stroke Outcomes in the Primary Analysis, by Vaccine Brand.

#### Inferential Results

There were no statistically significant associations of an increased risk of stroke following administration of the COVID-19 bivalent vaccines in the primary, seasonality adjusted, or PPV adjusted analyses (Figures 1, Figure 2, and eTable 13-15). The temporal scan did not identify any significant clusters for stroke outcomes after either COVID-19 bivalent vaccine (eFigure 2).

**Figure 1.**
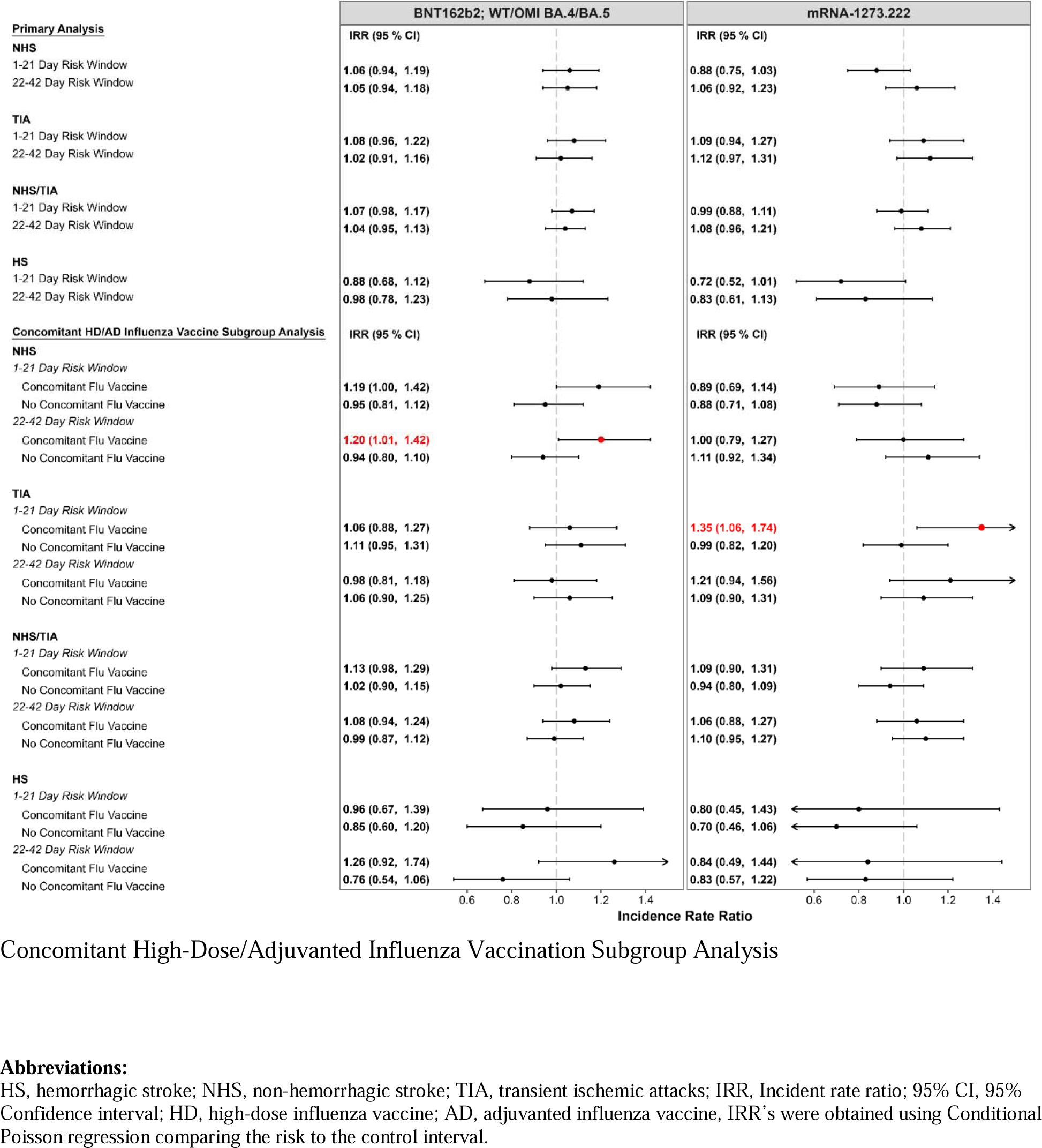
Risk of Stroke Following COVID-19 Bivalent Vaccines: Summary of Primary Analysis and

**Figure 2.**
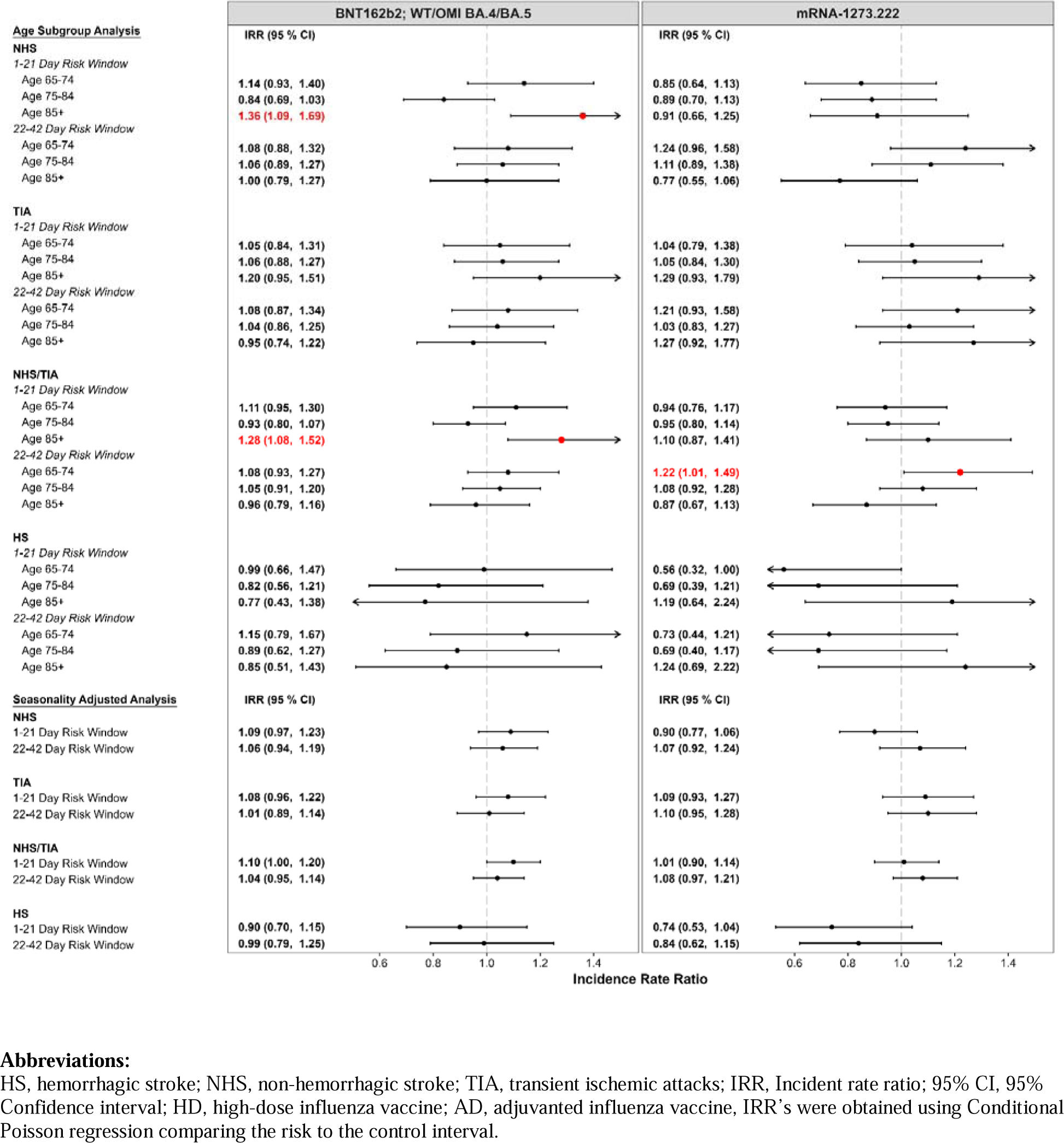
Risk of Stroke Following COVID-19 Bivalent Vaccines: Summary of Age Subgroup and Seasonality Adjusted Analyses

##### Age Subgroup Analyses

Among those who received the BNT162b2; WT/OMI BA.4/BA.5 vaccine, an increased risk was observed in the ≥85 population for NHS (IRR=1.36, 95% CI:1.09 – 1.69; AR/100,000 doses = 11.92, 95% CI:3.02 – 20.83 [1-21 days]) and for NHS/TIA (IRR=1.28, 95% CI:1.08 – 1.52; AR/100,000 doses= 15.34, 95% CI:4.34 – 26.34 [1-21 days]). Among those who received mRNA-1273.222, an increased risk was observed in the 65-74 age group for NHS/TIA (IRR=1.23, 95% CI:1.01 – 1.49; AR/100,000 doses = 3.26, 0.05 – 6.68 [22-42 days]). No elevated risk was observed for HS or TIA (Figure 2, eTable 16-17).

##### Concomitant Bivalent and High-Dose/Adjuvanted Influenza Vaccine Subgroup Analyses

Among those with concomitant high-dose/adjuvanted influenza and BNT162b2; WT/OMI BA.4/BA.5 vaccines, we observed an increased risk of NHS (IRR=1.20, 95% CI:1.01 – 1.42, AR/100,000 doses =3.13, 95% CI:0.05 – 6.21 [22-42 days]). Among those with concomitant high-dose/adjuvanted influenza and mRNA-1273.222 vaccines, an increased risk was observed for TIA (IRR=1.35, 95% CI: 1.06 – 1.74; AR/100,000 doses = 3.33, 95% CI 0.46 – 6.20 [1-21 days]). No risk was observed among those without a concomitant high-dose/adjuvanted influenza vaccine (Figure 1, eTable 18-19).

### Secondary Influenza Analyses

#### Descriptive Results

Given the elevated risk of stroke observed in the concomitant bivalent and influenza vaccine primary subgroup analysis, we conducted a secondary analysis to assess the risk of stroke following the administration of high-dose/adjuvanted influenza vaccines for the same study period. The study population included 6,961,413 eligible Medicare beneficiaries who received a high-dose/adjuvanted influenza vaccine. There were 5,497 cases of NHS, 4,871 cases of TIA, 9,065 cases of NHS/TIA, and 1,498 cases of HS. The baseline characteristics of the influenza vaccinated population are contained in eTable 20 and were similar to the COVID-19 bivalent vaccinated population.

#### Inferential Results

The secondary analysis identified a small, but statistically significant increased risk of NHS following high-dose/adjuvanted influenza vaccines (IRR= 1.09, 95% CI: 1.02 – 1.17; AR/100,000 doses = 1.65, 95% CI: 0.43 – 2.87 [22-42 days]) (Figure 3, eTable 21). The seasonality adjusted results additionally identified an increased risk of NHS/TIA in both risk intervals (IRR=1.06, 95% CI: 1.00 – 1.12; AR/100,000 doses = 3.91, 95% CI: 0.11 – 3.20 [1-21 days] and IRR=1.05, 95% CI: 1.00 – 1.11; AR/100,000 doses = 1.60, 95% CI: 0.02 – 3.18 [22-42 days]) (Figure 4, eTable 22). Results from the NHS PPV adjusted analysis were consistent with the original analysis (eTable 23).

**Figure 3.**
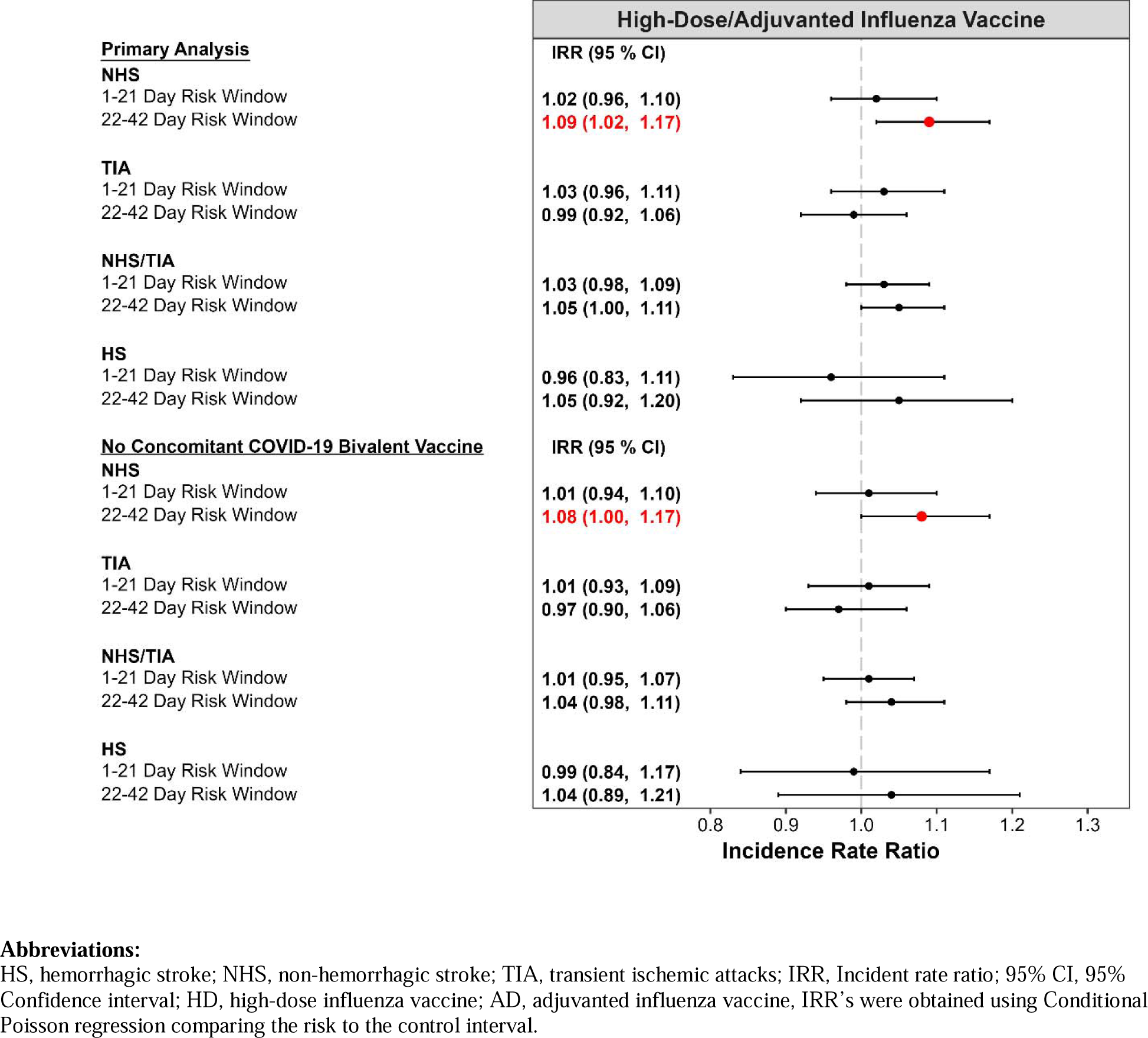
Risk of Stroke Following High-Dose/Adjuvanted Influenza Vaccines: Summary of Primary Analysis and no Concomitant COVID-19 Bivalent Vaccine Subgroup Analysis

**Figure 4.**
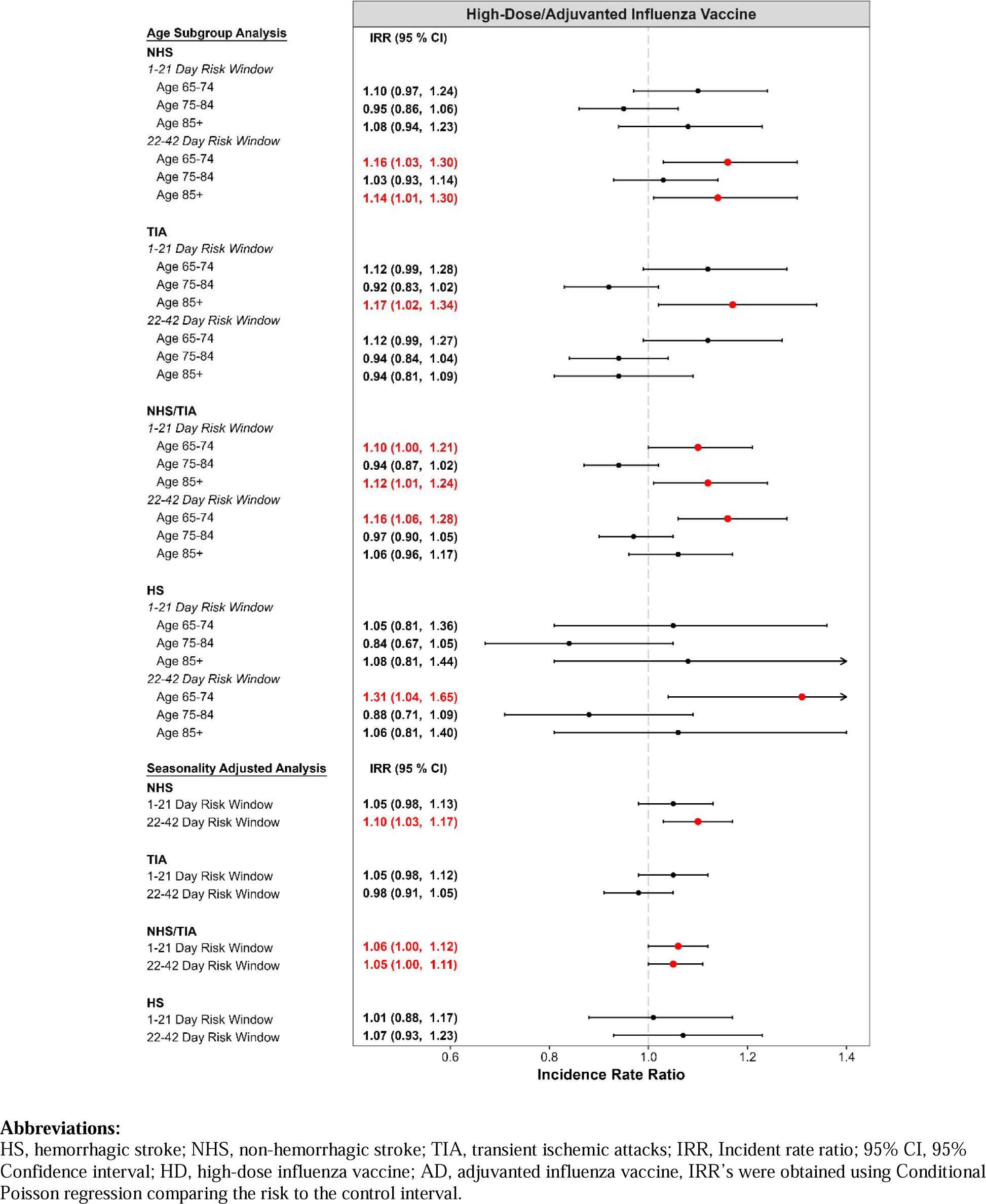
Risk of Stroke Following High-Dose/Adjuvanted Influenza Vaccines: Summary of Age Subgroup and Seasonality Adjusted Analyses

##### Age Subgroup High-Dose or Adjuvanted Influenza Vaccine Analyses

Elevated risk for all four stroke outcomes was observed in the age subgroup analyses.

For individuals in the 65-74 age group, an elevated risk was observed for HS (IRR=1.31, 95% CI: 1.04 – 1.65; AR=1.06, 0.13 – 1.98 [22-42 days]), NHS (IRR=1.16, 95% CI: 1.03 – 1.65; AR=1.68, 0.29 – 3.07 [22-42 days]), and NHS/TIA (IRR=1.10, 95% CI: 1.00 – 1.21; AR=1.79, 0.02 – 3.56 [1-21 days]. In the ≥85 age group, an elevated risk was observed for NHS (IRR=1.14, 95% CI: 1.01 – 1.30; AR=4.95, 0.21 – 9.69 [22-42 days]), TIA (IRR=1.01, 95% CI: 1.02 – 1.34; AR=3.77, 0.40 – 7.15 [1-21 days]) and NHS/TIA (IRR=1.12, 95% CI: 1.01 – 1.24; AR=6.57, 0.49 – 12.66 [1-21 days]). No increased risk was observed in the 75-84 age group (Figure 4, eTable 24).

##### Concomitant High-Dose/Adjuvanted Influenza and COVID-19 Bivalent Vaccine Subgroup Analyses

Among individuals who received a high-dose/adjuvanted influenza vaccine with no concomitant COVID-19 bivalent vaccine, we observed a slight, statistically significant elevated risk of NHS (IRR=1.08, 95% CI: 1.00 – 1.17; AR/100,000 doses = 1.47, 95% CI: 0.06 – 2.88 [22-42 days]) (Figure 3, eTable 25). This difference in risk was consistent with the risk observed among the concomitant BNT162b2; WT/OMI BA.4/BA.5 and high-dose/adjuvanted influenza vaccinated population.

## DISCUSSION

An earlier study on the CDC VSD data identified a preliminary signal of ischemic stroke following vaccination of persons ≥65 years with the COVID-19 bivalent vaccine.^3^ In this large U.S. population-based study of adults aged ≥65 years, we did not identify a statistically significant risk of stroke following vaccination with either the BNT162b2; WT/OMI BA.4/BA.5 or the mRNA-1273.222 vaccines in the primary analysis.

Our age subgroup analysis identified an elevated rate of NHS and NHS/TIA 1-21 days following BNT162b2; WT/OMI BA.4/BA.5 among the oldest age group (≥85 years). These findings are similar to the VSD study, where an increased risk of ischemic stroke was identified in the ≥85 age group.^3^ Our study did identify an elevated risk of stroke when the COVID-19 bivalent vaccines were administered with a concomitant high-dose/adjuvanted influenza vaccine. However, the observed effects were not consistent; elevated risk of NHS was detected 22-42 days after concomitant high-dose/adjuvanted influenza vaccines and BNT162b2; WT/OMI BA.4/BA.5 while the elevated risk of TIA was detected 1-21 days after concomitant use with mRNA-1273.222. Although a recent multicenter randomized controlled trial did not report elevated risk with concomitant use, the VSD study did identify a small increased risk of stroke following concomitant administration of the COVID-19 bivalent and high-dose/adjuvanted influenza vaccines.^3,17^

We additionally found a slightly elevated risk of stroke following influenza vaccines administered without concomitant COVID-19 bivalent vaccines. This finding suggests that the observed risk of stroke in the concomitant subgroup was likely driven by influenza vaccination alone rather than concomitant administration. These findings contradict previous SCCS studies by Asghar et. al and Sen et. al which do not report a risk of stroke associated with influenza vaccines.^18,19^ Several cohort studies in the literature report reduced risk of stroke following influenza vaccination when compared to the unvaccinated population.^20,21^ Because influenza illness is a known trigger of stroke, the apparent reduction of stroke risk following influenza vaccination in these studies can be partially attributed to reduced rates of influenza illness after vaccination.^20–23^ Moreover, a case control study by Vollmer et. al reported higher stroke odds ratio following influenza infection compared to vaccination.^24^ The clinical significance of the risk of stroke following vaccination must be carefully considered together with the significant benefits of receiving an influenza vaccination. Because the current framework of our SCCS study does not compare the vaccinated to the unvaccinated populations, it does not account for the reduced rate of severe influenza following vaccination. More studies are needed to better understand the association between influenza vaccination and stroke.

The results of our COVID-19 bivalent vaccine analysis contribute to a growing body of evidence that the COVID-19 mRNA bivalent vaccines are safe in the elderly population.^25,26^ Our findings are consistent with a study on the French National Health Data System, where no elevation in the risk of ischemic or hemorrhagic stroke was observed within 1-21 days following BNT162b2; WT/OMI BA.4/BA.5 in adults over 50 years.^27^ Consistent with our primary results, Gorenflo et al. found no risk of ischemic stroke in the BNT162b2; WT/OMI BA.4/BA.5 or mRNA-1273.222 COVID-19 bivalent vaccines among patients aged ≥65 years.^28^ Additionally, Yamin et al, investigated the safety profile of BNT162b2; WT/OMI BA.4/BA.5 in adults aged 60 and above and found no associated increase in the risk of ischemic stroke.^29^ Our study has several strengths. We used the CMS Medicare FFS database, a large, representative database with minimal beneficiary attrition, ensuring highly generalizable findings to the elderly U.S. population. Our study utilized self-controlled study designs, which inherently adjusted for time invariant confounders such as health conditions, socio-economic status, and health seeking behavior that may introduce bias in other study designs that draw from between-individual comparisons. Our study sampled vaccinated cases only and was not subject to bias due to underreporting of vaccination status in claims data. Finally, our study minimized outcome misclassification bias by performing medical record review of the claims-based NHS definition.

There are some limitations to our study. First, stroke is a known complication following COVID-19 diagnosis. Although we excluded cases with COVID-19 in the 30 days prior to outcome date, home antigen test-positive cases are not documented in claims records. This under capture of COVID-19 diagnoses may have biased our results and overestimated the risk of stroke.^30,31^ Second, the SCCS assumption that stroke events do not increase the probability of death was violated in our study. However, we minimized this bias by including these patients in the analysis using the Farrington adjustment.^15^ Third, we may have missed some beneficiaries with stroke outcomes or misclassified some beneficiaries who died before the end of follow-up because of delayed reporting. However, we accounted for claims delay in the calculation of the study end date for each outcome. Fourth, we conducted multiple tests which may increase the likelihood of detecting statistically significant results when there was no elevated risk. Finally, while we accounted for seasonality in the stroke outcomes, there may be additional bias from unmeasured time-varying covariates, such as circulating COVID-19 and influenza virus variants that may have altered the risk of stroke.

In our research on U.S. Medicare beneficiaries aged ≥65, the primary analysis did not find any statistically significant association of stroke following administration of COVID-19 mRNA bivalent vaccines. Our study found a small increase in the risk of NHS following high-dose/adjuvanted influenza vaccines. Overall our study supports the safety profile of the COVID-19 mRNA bivalent vaccines, and suggests the need for additional investigations into the safety of high-dose/adjuvanted influenza vaccines. The clinical significance of the slightly elevated NHS risk following influenza vaccination should be considered in the context of the risk of NHS following influenza infection in this population. However, our study results, in combination with the known benefits of COVID-19 and influenza vaccination, do not change the conclusion that the benefits of these vaccines outweigh their risks in persons ≥65 years.

## ACKNOWLEDGEMENTS

To Rowan McEvoy, Arnstein Lindaas, and Mao Hu, Acumen LLC, for contributions to methodology development and results review.

To Layo Laniyan, Acumen LLC, for contribution to manuscript writing and review.

## Author’s contributions

Yun Lu, Henry Zhang, Jeffrey Kelman, Richard Forshee, and Steven A. Anderson contributed to the study conception and design, data interpretation, manuscript writing and review. Kathryn Matuska, Gita Nadimpalli, Yuxin Ma, Nathan Duma, Ellen Chiang, and Hai Lyu contributed to study design, data collection and analysis, manuscript writing and review. All authors have seen and approved the manuscript.

## DATA AVAILABILITY

All data produced in the present work are contained in the manuscript.

