## Supplemental Tables and Figures for "Evaluation of Stroke Risk Following COVID-19 mRNA Bivalent Vaccines Among U.S. Adults Aged ≥65 Years"

**eTable 1. Summary of analytical plan and study specifications**

| **Analysis Specifications** | **Summary** |
| --- | --- |
| **Objective** | The analysis aims to assess the association of BNT162b2; WT/OMI BA.4/BA.5-BioNTech COVID-19 Vaccine, Bivalent and mRNA-1273.222 COVID-19 Vaccine, Bivalent booster doses (bivalent vaccines) administration with the four stroke outcomes in the Medicare population, accounting for potential sources of confounding using self-controlled study designs.  A secondary analysis aims to assess the association of high-dose or adjuvanted influenza vaccine administration using the same study design.  The adverse events were non-hemorrhagic stroke (NHS), transient ischemic attacks (TIA), NHS or TIA, and hemorrhagic stroke (HS). |
| **Data Sources** | We used Medicare claims and enrollment files from the Centers for Medicare and Medicaid Services (CMS) Medicare Shared Systems Data (SSD).  Demographics and information on death is derived from medicare enrollment databases. Information on vaccinations, health covariates, preventive services, and outcomes are derived from Medicare Part A (inpatient) and Part B (outpatient and community settings) and Part D (prescription) claims.  Beneficiaries' nursing home residence information was assessed using assessment data from the Minimum Data Set (MDS). |
| **Study Period** | August 31, 2022 - January 18, 2023 (NHS), February 4, 2023 (TIA), January 18, 2023 (NHS/TIA), and January 12, 2023 (HS) |
| **Exposures** | *Primary Analysis: COVID-19 Bivalent Vaccines*  BNT162b2; WT/OMI BA.4/BA.5 and mRNA-1273.222 vaccine receipt was identified using Current Procedural Terminology (CPT®)/Healthcare Common Procedure Coding System (HCPCS) codes or National Drug Codes (NDCs) in any care setting.  *Secondary Analysis: Influenza Vaccines*  High-dose or adjuvanted influenza vaccine receipt was identified using Current Procedural Terminology (CPT®)/Healthcare Common Procedure Coding System (HCPCS) codes or National Drug Codes (NDCs) in any care setting. |
| **Observation Period** | *Primary Analysis: COVID-19 Bivalent Vaccines*  For each individual, the observation period will start from the day following bivalent booster vaccination date, i.e., day 1 after vaccination and until the earlier of 90 days post-vaccination, disenrollment, administration of a subsequent mRNA COVID-19 vaccine, death or study period end.  *Secondary Analysis: Influenza Vaccines*  For each individual, the observation period will start from the day following high-dose or adjuvanted influenza vaccination date, i.e., day 1 after vaccination and until the earlier of 90 days post-vaccination, disenrollment, administration of a subsequent influenza vaccine, death or study period end. |
| **Risk Interval and Control Interval** | The risk interval is defined as the time during which risk of a stroke outcome is hypothesized to be related to vaccination based on biological plausibility and clinical input. The selected risk intervals for all stroke outcomes are days 1-21 and days 22-42.  The post-vaccination control interval is defined as all follow-up time during the observation period following a vaccination that is outside of the risk interval(s) in the observation period. |
| **Study Population** | *Primary Analysis: COVID-19 Bivalent Vaccines*  Eligible study population included those who received COVID-19 bivalent vaccine, experienced an incident adverse event in the observation period, and met the following inclusion/exclusion criteria:   (i) Medicare beneficiaries enrolled in Part A (hospital insurance) and Part B (medical insurance) during the study observation period (ii) Continuous enrollment from 365 days prior to vaccination date until the earlier of 90 days post-vaccination, disenrollment, administration of a subsequent mRNA COVID-19 vaccine, death or study period end. (iii) Received a bivalent booster in the observation period but not before the bivalent booster EUA, August 31, 2022. (iv) Had a record of a stroke outcome diagnosis during the study observation period. (v) At least 65 years of age at the time of vaccination (vi) Was not vaccinated with multiple brands on the same day or multiple vaccinations (same/different brand) within 3 days of each other (vii) Did not have a dialysis claim (identified as a type of bill code 72x) in the 90 days prior to event. (viii) Not on hospice care on vaccination date (ix) Do not reside in a nursing home at any point between August 31, 2022 and vaccination date. (x) Do not contribute follow-up time to both risk and control intervals. If individuals disenroll or reach the end of the study period during the risk window prior to accumulating any time for post-vaccination control intervals, they will be excluded. (xi) Do not have a diagnosis of a stroke outcome during the outcome-specific clean window. (xii) Do not have a COVID-19 diagnosis in the 30 days prior to the stroke outcome. (xiii) Do not have an exclusion code as listed in eTable 5.   *Secondary Analysis: Influenza Vaccines*  Eligible study population included those who received high-dose or adjuvanted influenza vaccines, experienced an incident adverse event in the observation period, and met the following inclusion/exclusion criteria:   (i) Medicare beneficiaries enrolled in Part A (hospital insurance) and Part B (medical insurance) during the study observation period (ii) Continuous enrollment from 365 days prior to vaccination date until the earlier of 90 days post-vaccination, disenrollment, administration of any subsequent influenza vaccine, death or study period end. (iii) Received a high-dose or adjuvanted influenza in the observation period without an influenza vaccine in the 60 days prior to the study start date, August 31, 2022. (iv) Had a record of a stroke outcome diagnosis during the study observation period. (v) At least 65 years of age at the time of vaccination (vi) Did not have multiple influenza vaccine codes on the same day, or multiple or duplicate vaccination codes observed within 3 days of each other. (vii) Did not have a dialysis claim (identified as a type of bill code 72x) in the 90 days prior to event. (viii) Not on hospice care on vaccination date (ix) Do not reside in a nursing home at any point between August 31, 2022 and vaccination date. (x) Do not contribute follow-up time to both risk and control intervals. If individuals disenroll or reach the end of the study period during the risk window prior to accumulating any time for post-vaccination control intervals, they will be excluded. (xi) Do not have a diagnosis of a stroke outcome during the outcome-specific clean window. (xii) Do not have an exclusion code as listed in eTable 5. |
| **Outcomes** | Incident occurrences of NHS, TIA, NHS/TIA, and HS were identified with International Classification of Diseases, Tenth Revision, Clinical Modification (ICD-10-CM) codes. The first occurrence of the event was defined as an incident case if no event was recorded during the preceding pre-defined interval, i.e., the clean window (eTable 4).  Case fatality was determined by identifying deaths that occurred following observation of a case during the observation period, based on CMS enrollment databases. |
| **Covariates** | The following covariates were assessed descriptively:   (i) Demographics were assessed at the time of COVID-19 vaccination receipt, including age, sex, race/ethnicity, region, urban/rural, dual eligibility status, area deprivation index (ADI), nursing home residency status (ii) Influenza vaccine receipt was assessed at the same time as the COVID-19 vaccine. (iii) History of a medically attended COVID-19 infection, defined by the ICD-10-CM codes U07.1 (COVID-19) were assessed using all available data (iv) Risk factors for the adverse event were assessed in the 365 days prior to index date |
| **Additional Confounding Control** | We controlled for seasonal variation in adverse event occurrence using outcome incidence estimated from 2021. |
| **Medical Record Review** | To validate our claims-based adverse event case definition, we retrieved a sample of NHS medical records, which underwent an adjudication process by board-certified clinicians. Cases were identified as true cases, non-cases, and potentially indeterminate using pre-determined clinical definitions |
| **Statistical Analysis** | Incident rate ratios (IRR), the ratio of the incidence in the exposure risk periods relative to the incidence in control periods, were estimated.  We also calculated absolute risk (AR), estimated as the number excess adverse events per 100,000 doses or person-years following vaccination, compared to if no vaccine was administered. |

**eTable 2. Outcome-specific claims delays and vaccine cutoff date**

| **Adverse Event** | **Claims Delay (days)** | **Study End Date** | **Vaccine Cutoff Date** |
| --- | --- | --- | --- |
| Hemorrhagic stroke (HS) | 78 | 1/12/2023 | 10/14/2022 |
| Non-hemorrhagic stroke (NHS) | 72 | 1/18/2023 | 10/20/202 |
| Transient ischemic attack (TIA) | 55 | 2/4/2023 | 11/6/2022 |
| Non-hemorrhagic stroke/ TIA (NHS/TIA) | 72 | 1/18/2023 | 10/20/2022 |

End dates were independently determined based on outcome specifc claims delay distributions

**eTable 3. HCPCS/CPT Codes for COVID-19 Vaccines**

| **HCPCS/CPT Code** | **Manufacturer** | **Name** | **Age Group** |
| --- | --- | --- | --- |
| 91312 | Pfizer-BioNTech | BNT162b2 Bivalent (WT/OMI BA.4/BA.5) | 12+ years |
| 91313 | Moderna | Moderna COVID-19 Vaccine, Bivalent | 18+ years |
| 90662 | Sanofi Pasteur | Fluzone High-Dose Quadrivalent (2022/2023) | 65+ years |
| 90694 | Seqirus | Fluad Quadrivalent (Adjuvanted) (2022/2023) | 65+ years |

**eTable 4. Stroke outcomes and their respective care settings, clean and risk windows**

| **Stroke Outcomes** | **ICD-10 Codes** | **Care Setting** | **Clean Window** | **Risk Windows** |
| --- | --- | --- | --- | --- |
|  |  | **(Primary and Secondary Analysis)** |  |  |
| Hemorrhagic stroke (HS) | I61*, I62* | IP | 365 days | 1-21 days; 22-42 days |
| Non-hemorrhagic stroke (NHS) | I63* | IP | 365 days | 1-21 days; 22-42 days |
| Transient ischemic attack (TIA) | G45.8, G45.9 | IP; OP-ED | 365 days | 1-21 days; 22-42 days |
| Non-hemorrhagic stroke or transient ischemic attack (NHS/TIA) | I63*, G45.8, G45.9 | IP (NHS & TIA); OP-ED (TIA) | 365 days | 1-21 days; 22-42 days |

Definitions: Clean Window is defined as an interval relative to stroke outcomes date used to define incident stroke outcomes where an individual enters the study cohort only if the stroke outcomes of interest did not occur during that interval. Risk Window is defined as an interval relative to exposure date during which occurrence of the stroke outcomes of interest will be included in the analyses

**eTable 5. ICD-10 Codes Which Indicate Onset Date Adjustment or Exclusion for Outcomes of Interest**

| **Outcome** | **Adjust Onset Date If Observed in the 1 Day Prior to Outcome** | **Exclusions for Prevalence** | **Exclusions – Other Known Causes** |
| --- | --- | --- | --- |
|  | **(in all settings)** | **(in all settings)** | **(in all settings)** |
| Hemorrhagic Stroke (HS) | I63.9, R51*, R47*, R29.810, R53.1, R42*, R41.82, R40.4, H53.13*, H53.9, G81.9* | If occurs in the 365 window prior to outcome | If in last 30 days prior to outcome |
|  |  | I69*, Z86.73 | U07.1 |
|  |  |  | If in last 1 day prior to outcome |
|  |  |  | S06* |
|  |  |  | If same day as outcome |
|  |  |  | S06*, Physical Trauma Code |
| Non-Hemorrhagic Stroke (NHS) | Z92.82, R51*, R47*, R29.810, R53.1, R42, R41.82, R40.4, G81.9*, H53.9, H53.13* | If occurs in the 365 window prior to outcome | If in last 30 days prior to outcome |
|  |  | I69*, Z86.73, I48*, D57*, D68.5* | U07.1 |
|  |  |  | If in last 28 days prior to outcome |
|  |  |  | I21* |
|  |  |  | If in last 1 day prior to outcome |
|  |  |  | S15*, 174* |
|  |  |  | In same day as outcome |
|  |  |  | S15*, I74*, Physical Trauma Code |
| Non-hemorrhagic stroke or Transient Ischemic Attacks (NHS / TIA) | Z92.82, R51*, R47*, R29.810, R53.1, R42, R41.82, R40.4, G81.9*, H53.9, H53.13* | If occurs in the 365 window prior to outcome | If in last 30 days prior to outcome |
|  |  | I69*, Z86.73, | U07.1 |
|  |  | I48*, D57*, D68.5* | If in last 28 days prior to outcome |
|  |  |  | I21* |
|  |  |  | If in last 1 day prior to outcome |
|  |  |  | S15*, 174* |
|  |  |  | In same day as outcome |
|  |  |  | S15*, I74*, Physical Trauma Code |
| Transient Ischemic Attacks (TIA) | R51*, R47*, R29.810, R53.1, R42, R41.82, R40.4, G81.9*, H53.9, H53.13* | If occurs in the 365 window prior to outcome | If in last 30 days prior to outcome |
|  |  | I69*, Z86.73, I48*, D57*, D68.5* | U07.1 |
|  |  |  | If in last 28 days prior to outcome |
|  |  |  | I21* |
|  |  |  | If in last 1 day prior to outcome |
|  |  |  | S15* |
|  |  |  | In same day as outcome |
|  |  |  | S15*, Physical Trauma Code |

**eTable 6. Summary of death cases following AESI, by risk/control interval, exposure, and AESI**

| **AESI** | **Exposure** | **Interval** | **# of Cases** | **Cases That Died** | | | | | | | | **Total Control Time Accrued** |
| --- | --- | --- | --- | --- | --- | --- | --- | --- | --- | --- | --- | --- |
|  |  |  |  | **Overall** | | **Risk 1** | | **Risk 2** | | **Control** | |  |
|  |  |  |  | **#** | **%** | **#** | **%** | **#** | **%** | **#** | **%** | **# days** |
| NHS | Moderna Bivalent | Overall | 1,090 | 108 | 9.91% | * | * | * | * | 80 | 7.34% | 49,149 |
|  |  | Risk 1 | 215 | 19 | 8.84% | * | * | * | * | * | * | 9,450 |
|  |  | Risk 2 | 268 | 31 | 11.57% |  |  | 13 | 4.85% | 18 | 6.72% | 11,683 |
|  |  | Control | 607 | 58 | 9.56% |  |  |  |  | 58 | 9.56% | 28,016 |
|  | Pfizer Bivalent | Overall | 1,796 | 200 | 11.14% | 23 | 1.28% | 43 | 2.39% | 134 | 7.46% | 79,659 |
|  |  | Risk 1 | 425 | 60 | 14.12% | 23 | 5.41% | 21 | 4.94% | 16 | 3.76% | 17,736 |
|  |  | Risk 2 | 418 | 56 | 13.40% |  |  | 22 | 5.26% | 34 | 8.13% | 17,817 |
|  |  | Control | 953 | 84 | 8.81% |  |  |  |  | 84 | 8.81% | 44,106 |
|  | High Dose or Adjuvanted Influenza | Overall | 3,288 | 406 | 12.35% | 45 | 1.37% | 90 | 2.74% | 271 | 8.24% | 141,210 |
|  |  | Risk 1 | 830 | 133 | 16.02% | 45 | 5.42% | 47 | 5.66% | 41 | 4.94% | 32,778 |
|  |  | Risk 2 | 800 | 117 | 14.62% |  |  | 43 | 5.38% | 74 | 9.25% | 32,489 |
|  |  | Control | 1,658 | 156 | 9.41% |  |  |  |  | 156 | 9.41% | 75,943 |
| TIA | Moderna Bivalent | Overall | 1,035 | 12 | 1.16% | * | * | * | * | * | * | 48,958 |
|  |  | Risk 1 | 245 | * | * | * | * | * | * | * | * | 11,498 |
|  |  | Risk 2 | 254 | * | * |  |  | * | * | * | * | 11,949 |
|  |  | Control | 536 | * | * |  |  |  |  | * | * | 25,511 |
|  | Pfizer Bivalent | Overall | 1,606 | 15 | 0.93% | * | * | * | * | * | * | 76,135 |
|  |  | Risk 1 | 389 | * | * | * | * | * | * | * | * | 18,223 |
|  |  | Risk 2 | 368 | * | * |  |  | * | * | * | * | 17,389 |
|  |  | Control | 849 | * | * |  |  |  |  | * | * | 40,523 |
|  | High Dose or Adjuvanted Influenza | Overall | 2,867 | 33 | 1.15% | * | * | * | * | 24 | 0.84% | 131,724 |
|  |  | Risk 1 | 693 | 12 | 1.73% | * | * | * | * | * | * | 31,016 |
|  |  | Risk 2 | 660 | * | * |  |  | * | * | * | * | 29,520 |
|  |  | Control | 1,514 | 11 | 0.73% |  |  |  |  | 11 | 0.73% | 71,188 |
| NHS/TIA | Moderna Bivalent | Overall | 1,811 | 112 | 6.18% | * | * | * | * | 82 | 4.53% | 83,476 |
|  |  | Risk 1 | 394 | 22 | 5.58% | * | * | 11 | 2.79% | * | * | 17,912 |
|  |  | Risk 2 | 440 | 31 | 7.05% |  |  | 13 | 2.95% | 18 | 4.09% | 19,893 |
|  |  | Control | 977 | 59 | 6.04% |  |  |  |  | 59 | 6.04% | 45,671 |
|  | Pfizer Bivalent | Overall | 2,977 | 209 | 7.02% | 26 | 0.87% | 43 | 1.44% | 140 | 4.70% | 135,904 |
|  |  | Risk 1 | 712 | 67 | 9.41% | 26 | 3.65% | 22 | 3.09% | 19 | 2.67% | 31,207 |
|  |  | Risk 2 | 686 | 58 | 8.45% |  |  | 21 | 3.06% | 37 | 5.39% | 30,614 |
|  |  | Control | 1,579 | 84 | 5.32% |  |  |  |  | 84 | 5.32% | 74,083 |
|  | High Dose or Adjuvanted Influenza | Overall | 5,328 | 428 | 8.03% | 46 | 0.86% | 95 | 1.78% | 287 | 5.39% | 234,544 |
|  |  | Risk 1 | 1,316 | 143 | 10.87% | 46 | 3.50% | 50 | 3.80% | 47 | 3.57% | 54,388 |
|  |  | Risk 2 | 1,275 | 121 | 9.49% |  |  | 45 | 3.53% | 76 | 5.96% | 53,590 |
|  |  | Control | 2,737 | 164 | 5.99% |  |  |  |  | 164 | 5.99% | 126,566 |
| HS | Moderna Bivalent | Overall | 283 | 64 | 22.61% | * | * | 15 | 5.30% | 45 | 15.90% | 11,740 |
|  |  | Risk 1 | 50 | 13 | 26.00% | * | * | * | * | * | * | 1,832 |
|  |  | Risk 2 | 61 | 19 | 31.15% |  |  | * | * | 11 | 18.03% | 2,204 |
|  |  | Control | 172 | 32 | 18.60% |  |  |  |  | 32 | 18.60% | 7,704 |
|  | Pfizer Bivalent | Overall | 525 | 158 | 30.10% | 18 | 3.43% | 39 | 7.43% | 101 | 19.24% | 20,103 |
|  |  | Risk 1 | 107 | 37 | 34.58% | 18 | 16.82% | * | * | * | * | 3,519 |
|  |  | Risk 2 | 123 | 45 | 36.59% |  |  | 29 | 23.58% | 16 | 13.01% | 3,920 |
|  |  | Control | 295 | 76 | 25.76% |  |  |  |  | 76 | 25.76% | 12,664 |
|  | High Dose or Adjuvanted Influenza | Overall | 870 | 303 | 34.83% | 62 | 7.13% | 72 | 8.28% | 169 | 19.43% | 30,261 |
|  |  | Risk 1 | 243 | 110 | 45.27% | 62 | 25.51% | 29 | 11.93% | 19 | 7.82% | 6,237 |
|  |  | Risk 2 | 204 | 77 | 37.75% |  |  | 43 | 21.08% | 34 | 16.67% | 6,302 |
|  |  | Control | 423 | 116 | 27.42% |  |  |  |  | 116 | 27.42% | 17,722 |

**Abbreviations**: HS, hemorrhagic stroke; NHS, non-hemorrhagic stroke; TIA, transient ischemic attacks

* Outcome counts of 10 or fewer and associated statistics are masked to protect the anonymity of the data.

**eTable 7. Summary of NHS Medical Record Review Case Adjudication Results and Associated PPVs**

| **Category** | **NHS Cases Received** |
| --- | --- |
| **Final Case Classifications** | **87** |
| Confirmed | 58 |
| Probable | 12 |
| Possible | 12 |
| Insufficient Evidence | 2 |
| Not a Case | 3 |
| PPV (Confirmed + Probable) | 80.46% (70.92, 87.43) |

**eTable 8. 30 Day Case Fatality Rate by Outcome; October 31, 2021 - October 31, 2022**

| **Stroke outcomes** | Non-hemorrhagic Stroke (**NHS)** | Transient Ischemic Attack (**TIA)** | Hemorrhagic Stroke (**HS)** |
| --- | --- | --- | --- |
| Case Fatality Rate | 17.0% | 2.1% | 34.3% |
| The combined NHS/TIA outcome is expected to have a similar CFR to the NHS only outcome. | | | |

**eTable 9. Summary of Demographics, Socio-Economic Status, Residence, Health Status, and Healthcare Utilization of BNT162b2; WT/OMI BA.4/BA.5 Bivalent Vaccinated Medicare Beneficiaries with Stroke Outcomes in the Primary Analysis, by Age**

| **Patient Characteristics** | **BNT162b2; WT/OMI BA.4/BA.5** | | | | | | | | | | | |
| --- | --- | --- | --- | --- | --- | --- | --- | --- | --- | --- | --- | --- |
|  | **65-74** | | | | **75-84** | | | | **85+** | | | |
|  | **NHS**  n (%) | **TIA**  n (%) | **NHS/ TIA**  n (%) | **HS**  n (%) | **NHS**  n (%) | **TIA**  n (%) | **NHS/ TIA**  n (%) | **HS**  n (%) | **NHS**  n (%) | **TIA**  n (%) | **NHS/ TIA**  n (%) | **HS**  n (%) |
| **Total** | **586** | **497** | **969** | **182** | **732** | **706** | **1251** | **218** | **478** | **403** | **757** | **125** |
| **Sex** | | | | | | | | | | | | |
| Female | 281 (47.95) | 279 (56.14) | 499 (51.50) | 83 (45.60) | 391 (53.42) | 390 (55.24) | 676 (54.04) | 90 (41.28) | 323 (67.57) | 280 (69.48) | 516 (68.16) | 75 (60.00) |
| Male | 305 (52.05) | 218 (43.86) | 470 (48.50) | 99 (54.40) | 341 (46.58) | 316 (44.76) | 575 (45.96) | 128 (58.72) | 155 (32.43) | 123 (30.52) | 241 (31.84) | 50 (40.00) |
| Missing/Unknown | 0 (0.00) | 0 (0.00) | 0 (0.00) | 0 (0.00) | 0 (0.00) | 0 (0.00) | 0 (0.00) | 0 (0.00) | 0 (0.00) | 0 (0.00) | 0 (0.00) | 0 (0.00) |
| **Race/Ethnicity** | | | | | | | | | | | | |
| Asian | *  (*) | *  (*) | 18 (1.86) | *  (*) | *  (*) | *  (*) | 18 (1.44) | *  (*) | 16 (3.35) | *  (*) | 20 (2.64) | *  (*) |
| Black | 44 (7.51) | 27 (5.43) | 60 (6.19) | *  (*) | 42 (5.74) | 22 (3.12) | 55 (4.40) | *  (*) | 27 (5.65) | 20 (4.96) | 38 (5.02) | *  (*) |
| Hispanic | *  (*) | *  (*) | *  (*) | *  (*) | *  (*) | *  (*) | *  (*) | *  (*) | *  (*) | *  (*) | *  (*) | 0 (0.00) |
| Alaskan Native/Native American | *  (*) | *  (*) | *  (*) | 0 (0.00) | *  (*) | 0 (0.00) | *  (*) | 0 (0.00) | *  (*) | *  (*) | *  (*) | 0 (0.00) |
| White | 491 (83.79) | 423 (85.11) | 821 (84.73) | 160 (87.91) | 646 (88.25) | 653 (92.49) | 1,129 (90.25) | 192 (88.07) | 416 (87.03) | 373 (92.56) | 675 (89.17) | 115 (92.00) |
| Other | *  (*) | *  (*) | *  (*) | *  (*) | 19 (2.60) | 12 (1.70) | 28 (2.24) | *  (*) | 13 (2.72) | *  (*) | 15 (1.98) | *  (*) |
| Missing/Unknown | 31 (5.29) | 30 (6.04) | 55 (5.68) | *  (*) | *  (*) | *  (*) | 15 (1.20) | *  (*) | 0 (0.00) | 0 (0.00) | 0 (0.00) | 0 (0.00) |
| **Urban/Rural** | | | | | | | | | | | | |
| Urban | 508 (86.69) | 414 (83.30) | 819 (84.52) | 157 (86.26) | 639 (87.30) | 602 (85.27) | 1,086 (86.81) | 186 (85.32) | 421 (88.08) | 336 (83.37) | 658 (86.92) | 113 (90.40) |
| Rural | *  (*) | *  (*) | *  (*) | *  (*) | *  (*) | *  (*) | *  (*) | *  (*) | 57 (11.92) | 67 (16.63) | 99 (13.08) | *  (*) |
| Missing/Unknown | *  (*) | *  (*) | *  (*) | *  (*) | *  (*) | *  (*) | *  (*) | *  (*) | 0 (0.00) | 0 (0.00) | 0 (0.00) | *  (*) |
| **HHS Region** | | | | | | | | | | | | |
| Region 1 | 48 (8.19) | 35 (7.04) | 77 (7.95) | *  (*) | 64 (8.74) | 65 (9.21) | 111 (8.87) | 14 (6.42) | 41 (8.58) | 36 (8.93) | 63 (8.32) | *  (*) |
| Region 2 | 39 (6.66) | 37 (7.44) | 65 (6.71) | *  (*) | 62 (8.47) | 48 (6.80) | 95 (7.59) | 22 (10.09) | 52 (10.88) | 42 (10.42) | 82 (10.83) | 19 (15.20) |
| Region 3 | 80 (13.65) | 63 (12.68) | 127 (13.11) | 26 (14.29) | 94 (12.84) | 88 (12.46) | 155 (12.39) | 29 (13.30) | 76 (15.90) | 45 (11.17) | 109 (14.40) | *  (*) |
| Region 4 | 111 (18.94) | 88 (17.71) | 185 (19.09) | 37 (20.33) | 131 (17.90) | 132 (18.70) | 231 (18.47) | 22 (10.09) | 58 (12.13) | 72 (17.87) | 111 (14.66) | 26 (20.80) |
| Region 5 | 130 (22.18) | 96 (19.32) | 205 (21.16) | 33 (18.13) | 157 (21.45) | 132 (18.70) | 252 (20.14) | 52 (23.85) | 105 (21.97) | 70 (17.37) | 153 (20.21) | 25 (20.00) |
| Region 6 | 41 (7.00) | 46 (9.26) | 73 (7.53) | *  (*) | 62 (8.47) | 52 (7.37) | 100 (7.99) | 19 (8.72) | 33 (6.90) | 26 (6.45) | 51 (6.74) | *  (*) |
| Region 7 | 34 (5.80) | 30 (6.04) | 57 (5.88) | *  (*) | 37 (5.05) | 31 (4.39) | 59 (4.72) | 12 (5.50) | 28 (5.86) | 25 (6.20) | 45 (5.94) | *  (*) |
| Region 8 | 25 (4.27) | 26 (5.23) | *  (*) | 11 (6.04) | 23 (3.14) | *  (*) | *  (*) | *  (*) | 15 (3.14) | 20 (4.96) | 29 (3.83) | *  (*) |
| Region 9 | 54 (9.22) | 46 (9.26) | 90 (9.29) | 35 (19.23) | 69 (9.43) | 89 (12.61) | 137 (10.95) | 30 (13.76) | 46 (9.62) | 51 (12.66) | 80 (10.57) | *  (*) |
| Region 10 | *  (*) | 30 (6.04) | 46 (4.75) | *  (*) | 33 (4.51) | 41 (5.81) | 66 (5.28) | *  (*) | 24 (5.02) | 16 (3.97) | 34 (4.49) | *  (*) |
| Missing/Unknown | *  (*) | 0 (0.00) | *  (*) | 0 (0.00) | 0 (0.00) | *  (*) | *  (*) | 0 (0.00) | 0 (0.00) | 0 (0.00) | 0 (0.00) | 0 (0.00) |
| **Dual-Eligibility Status**** | | | | | | | | | | | | |
| Dual-Eligible | 21 (3.58) | 21 (4.23) | 32 (3.30) | *  (*) | 21 (2.87) | 20 (2.83) | 31 (2.48) | *  (*) | 25 (5.23) | 13 (3.23) | 34 (4.49) | *  (*) |
| Non-Dual-Eligible | 565 (96.42) | 476 (95.77) | 937 (96.70) | 171 (93.96) | 711 (97.13) | 686 (97.17) | 1,220 (97.52) | 212 (97.25) | 453 (94.77) | 390 (96.77) | 723 (95.51) | 121 (96.80) |
| **Area Deprivation Index (ADI) Rank** | | | | | | | | | | | | |
| 1-10 (lowest level of deprivation or disadvantage) | 80 (13.65) | 76 (15.29) | 140 (14.45) | 30 (16.48) | 143 (19.54) | 131 (18.56) | 249 (19.90) | 46 (21.10) | 81 (16.95) | 76 (18.86) | 136 (17.97) | 23 (18.40) |
| 11-20 | 95 (16.21) | 76 (15.29) | 158 (16.31) | 28 (15.38) | 117 (15.98) | 128 (18.13) | 209 (16.71) | 36 (16.51) | 67 (14.02) | 49 (12.16) | 100 (13.21) | 20 (16.00) |
| 21-30 | 73 (12.46) | 69 (13.88) | 124 (12.80) | 32 (17.58) | 114 (15.57) | 85 (12.04) | 182 (14.55) | 39 (17.89) | 74 (15.48) | 64 (15.88) | 120 (15.85) | 20 (16.00) |
| 31-40 | 78 (13.31) | 71 (14.29) | 133 (13.73) | 26 (14.29) | 92 (12.57) | 83 (11.76) | 149 (11.91) | 23 (10.55) | 75 (15.69) | 45 (11.17) | 100 (13.21) | 18 (14.40) |
| 41-50 | 63 (10.75) | 49 (9.86) | 102 (10.53) | 19 (10.44) | 76 (10.38) | 80 (11.33) | 132 (10.55) | 22 (10.09) | 50 (10.46) | 41 (10.17) | 78 (10.30) | *  (*) |
| 51-60 | 58 (9.90) | 55 (11.07) | 96 (9.91) | 23 (12.64) | 59 (8.06) | 66 (9.35) | 104 (8.31) | 22 (10.09) | 33 (6.90) | 35 (8.68) | 57 (7.53) | 14 (11.20) |
| 61-70 | 55 (9.39) | 34 (6.84) | 83 (8.57) | *  (*) | 44 (6.01) | 41 (5.81) | 71 (5.68) | *  (*) | 41 (8.58) | 35 (8.68) | 64 (8.45) | *  (*) |
| 71-80 | 28 (4.78) | 24 (4.83) | 48 (4.95) | *  (*) | 30 (4.10) | 45 (6.37) | 63 (5.04) | *  (*) | 27 (5.65) | 21 (5.21) | 45 (5.94) | *  (*) |
| 81-90 | 24 (4.10) | 23 (4.63) | 41 (4.23) | *  (*) | 22 (3.01) | 17 (2.41) | 34 (2.72) | *  (*) | *  (*) | 16 (3.97) | 18 (2.38) | *  (*) |
| 91-100 | 14 (2.39) | *  (*) | 20 (2.06) | *  (*) | 20 (2.73) | 12 (1.70) | 28 (2.24) | *  (*) | 14 (2.93) | *  (*) | 22 (2.91) | *  (*) |
| Missing/Unknown | 18 (3.07) | *  (*) | 24 (2.48) | *  (*) | 15 (2.05) | 18 (2.55) | 30 (2.40) | *  (*) | *  (*) | *  (*) | 17 (2.25) | *  (*) |
| **Medicare Status** | | | | | | | | | | | | |
| Aged-in without ESRD | 514 (87.71) | 444 (89.34) | 862 (88.96) | 151 (82.97) | 691 (94.40) | 679 (96.18) | 1,192 (95.28) | 206 (94.50) | 465 (97.28) | 397 (98.51) | 739 (97.62) | 124 (99.20) |
| Aged & Disabled with ESRD | *  (*) | *  (*) | *  (*) | *  (*) | 0 (0.00) | *  (*) | 0 (0.00) | 0 (0.00) | 0 (0.00) | 0 (0.00) | 0 (0.00) | 0 (0.00) |
| Disabled without ESRD | 71 (12.12) | 51 (10.26) | 105 (10.84) | 29 (15.93) | 41 (5.60) | 26 (3.68) | 59 (4.72) | 12 (5.50) | 13 (2.72) | *  (*) | 18 (2.38) | *  (*) |
| Missing/Unknown | *  (*) | *  (*) | *  (*) | *  (*) | 0 (0.00) | *  (*) | 0 (0.00) | 0 (0.00) | 0 (0.00) | *  (*) | 0 (0.00) | *  (*) |
| **Medical Conditions (0-365 days prior to vaccination date)** | | | | | | | | | | | | |
| Asthma | 40 (6.83) | 50 (10.06) | 80 (8.26) | *  (*) | 67 (9.15) | 58 (8.22) | 115 (9.19) | 18 (8.26) | 23 (4.81) | 28 (6.95) | 49 (6.47) | *  (*) |
| COPD | 61 (10.41) | 47 (9.46) | 96 (9.91) | 27 (14.84) | 103 (14.07) | 81 (11.47) | 162 (12.95) | 30 (13.76) | 52 (10.88) | 38 (9.43) | 79 (10.44) | 12 (9.60) |
| Chronic Kidney Disease | 114 (19.45) | 97 (19.52) | 178 (18.37) | 38 (20.88) | 198 (27.05) | 176 (24.93) | 314 (25.10) | 61 (27.98) | 162 (33.89) | 141 (34.99) | 259 (34.21) | 55 (44.00) |
| Depression | 94 (16.04) | 105 (21.13) | 176 (18.16) | 36 (19.78) | 135 (18.44) | 133 (18.84) | 232 (18.55) | 30 (13.76) | 81 (16.95) | 70 (17.37) | 136 (17.97) | 17 (13.60) |
| Gout | 33 (5.63) | 28 (5.63) | 51 (5.26) | *  (*) | 52 (7.10) | 53 (7.51) | 91 (7.27) | 13 (5.96) | 28 (5.86) | 27 (6.70) | 48 (6.34) | *  (*) |
| Heart Failure | 61 (10.41) | 36 (7.24) | 81 (8.36) | 28 (15.38) | 100 (13.66) | 92 (13.03) | 167 (13.35) | 46 (21.10) | 75 (15.69) | 70 (17.37) | 119 (15.72) | 29 (23.20) |
| Hypercholesterolemia | 125 (21.33) | 99 (19.92) | 201 (20.74) | 42 (23.08) | 141 (19.26) | 172 (24.36) | 262 (20.94) | 54 (24.77) | 94 (19.67) | 95 (23.57) | 157 (20.74) | 27 (21.60) |
| Hypothyroidism | 405 (69.11) | 326 (65.59) | 650 (67.08) | 142 (78.02) | 575 (78.55) | 541 (76.63) | 964 (77.06) | 165 (75.69) | 399 (83.47) | 326 (80.89) | 621 (82.03) | 109 (87.20) |
| Hypertension | 111 (18.94) | 119 (23.94) | 199 (20.54) | 45 (24.73) | 173 (23.63) | 194 (27.48) | 307 (24.54) | 46 (21.10) | 141 (29.50) | 124 (30.77) | 229 (30.25) | 25 (20.00) |
| ITP | *  (*) | *  (*) | *  (*) | *  (*) | *  (*) | *  (*) | *  (*) | *  (*) | *  (*) | *  (*) | *  (*) | 0 (0.00) |
| Impaired Mobility | *  (*) | *  (*) | *  (*) | *  (*) | *  (*) | *  (*) | *  (*) | *  (*) | *  (*) | *  (*) | *  (*) | *  (*) |
| Ischemic Heart Disease | 31 (5.29) | 16 (3.22) | 44 (4.54) | *  (*) | 34 (4.64) | 30 (4.25) | 53 (4.24) | 16 (7.34) | 20 (4.18) | 20 (4.96) | 33 (4.36) | *  (*) |
| Nicotine Dependency | 156 (26.62) | 101 (20.32) | 221 (22.81) | 50 (27.47) | 181 (24.73) | 141 (19.97) | 286 (22.86) | 66 (30.28) | 77 (16.11) | 83 (20.60) | 130 (17.17) | 25 (20.00) |
| Obesity | 179 (30.55) | 146 (29.38) | 286 (29.51) | 55 (30.22) | 139 (18.99) | 149 (21.10) | 252 (20.14) | 59 (27.06) | 55 (11.51) | 43 (10.67) | 85 (11.23) | 13 (10.40) |
| **Charlson Comorbidity Index** | | | | | | | | | | | | |
| 0 | 193 (32.94) | 151 (30.38) | 314 (32.40) | 48 (26.37) | 162 (22.13) | 167 (23.65) | 292 (23.34) | 48 (22.02) | 93 (19.46) | 68 (16.87) | 143 (18.89) | 17 (13.60) |
| 1 | 109 (18.60) | 83 (16.70) | 171 (17.65) | 37 (20.33) | 125 (17.08) | 117 (16.57) | 217 (17.35) | 26 (11.93) | 86 (17.99) | 77 (19.11) | 136 (17.97) | 15 (12.00) |
| 2 | 93 (15.87) | 95 (19.11) | 171 (17.65) | 28 (15.38) | 122 (16.67) | 132 (18.70) | 221 (17.67) | 30 (13.76) | 84 (17.57) | 78 (19.35) | 136 (17.97) | 24 (19.20) |
| 3 | 60 (10.24) | 58 (11.67) | 108 (11.15) | 17 (9.34) | 89 (12.16) | 111 (15.72) | 165 (13.19) | 28 (12.84) | 71 (14.85) | 61 (15.14) | 114 (15.06) | 23 (18.40) |
| 4 | 45 (7.68) | 39 (7.85) | 71 (7.33) | 14 (7.69) | 90 (12.30) | 65 (9.21) | 138 (11.03) | 27 (12.39) | 56 (11.72) | 44 (10.92) | 86 (11.36) | 18 (14.40) |
| 5+ | 86 (14.68) | 71 (14.29) | 134 (13.83) | 38 (20.88) | 144 (19.67) | 114 (16.15) | 218 (17.43) | 59 (27.06) | 88 (18.41) | 75 (18.61) | 142 (18.76) | 28 (22.40) |
| **Prior COVID-19 diagnosis** | | | | | | | | | | | | |
| In the 30 days prior to outcome | 0 (0.00) | 0 (0.00) | 0 (0.00) | 0 (0.00) | 0 (0.00) | 0 (0.00) | 0 (0.00) | 0 (0.00) | 0 (0.00) | 0 (0.00) | 0 (0.00) | 0 (0.00) |
| In the 31-365 days prior to outcome | 44 (7.51) | 93 (18.71) | 114 (11.76) | 19 (10.44) | 88 (12.02) | 101 (14.31) | 158 (12.63) | 31 (14.22) | 55 (11.51) | 39 (9.68) | 79 (10.44) | *  (*) |
| **Concomitant Influenza Vaccination (same day as bivalent booster)** | | | | | | | | | | | | |
| High-dose | 156 (26.62) | 160 (32.19) | 279 (28.79) | 49 (26.92) | 216 (29.51) | 197 (27.90) | 365 (29.18) | 75 (34.40) | 139 (29.08) | 109 (27.05) | 220 (29.06) | 26 (20.80) |
| Adjuvanted | 104 (17.75) | 91 (18.31) | 170 (17.54) | 32 (17.58) | 108 (14.75) | 85 (12.04) | 171 (13.67) | 33 (15.14) | 74 (15.48) | 62 (15.38) | 124 (16.38) | 30 (24.00) |
| Recombinant | *  (*) | *  (*) | *  (*) | *  (*) | *  (*) | 0 (0.00) | *  (*) | 0 (0.00) | 0 (0.00) | *  (*) | *  (*) | *  (*) |
| Live-attenuated | 0 (0.00) | 0 (0.00) | 0 (0.00) | 0 (0.00) | 0 (0.00) | 0 (0.00) | 0 (0.00) | 0 (0.00) | 0 (0.00) | 0 (0.00) | 0 (0.00) | 0 (0.00) |
| Cell-cultured | *  (*) | *  (*) | *  (*) | *  (*) | *  (*) | *  (*) | *  (*) | *  (*) | *  (*) | *  (*) | *  (*) | 0 (0.00) |
| Standard | *  (*) | *  (*) | *  (*) | *  (*) | *  (*) | *  (*) | *  (*) | *  (*) | *  (*) | *  (*) | *  (*) | *  (*) |
| **Other vaccine (administered in risk/control intervals)** | | | | | | | | | | | | |
| Pneumococcal vaccine | 19 (3.24) | 17 (3.42) | 34 (3.51) | *  (*) | 18 (2.46) | 22 (3.12) | 34 (2.72) | *  (*) | *  (*) | *  (*) | 15 (1.98) | *  (*) |

**Abbreviations:** BNT162b2; WT/OMI BA.4/BA.5, Pfizer COVID-19 BivalentmRNA-1273.222, Moderna COVID-19 Bivalent;

HS, hemorrhagic stroke; NHS, non-hemorrhagic stroke; TIA, transient ischemic attacks; ADI, area deprivations index; ESRD, end-stage renal disease; COPD, chronic obstructive pulmonary disease; COVID-19, coronavirus 2019

* Outcome counts of 10 or fewer and associated statistics are masked to protect the anonymity of the data.

** Reflects beneficiaries’ dual eligibility for Medicare and Medicaid plan benefits.

**eTable 10. Summary of Demographics, Socio-Economic Status, Residence, Health Status, and Healthcare Utilization of MRNA-1273.222 Bivalent Vaccinated Medicare Beneficiaries with Stroke Outcomes in the Primary Analysis, by Age**

| **Patient Characteristics** | **MRNA-1273.222** | | | | | | | | | | | |
| --- | --- | --- | --- | --- | --- | --- | --- | --- | --- | --- | --- | --- |
|  | **65-74** | | | | **75-84** | | | | **85+** | | | |
|  | **NHS**  n (%) | **TIA**  n (%) | **NHS/ TIA**  n (%) | **HS**  n (%) | **NHS**  n (%) | **TIA**  n (%) | **NHS/ TIA**  n (%) | **HS**  n (%) | **NHS**  n (%) | **TIA**  n (%) | **NHS/ TIA**  n (%) | **HS**  n (%) |
| **Total** | **352** | **315** | **579** | **106** | **474** | **507** | **830** | **103** | **264** | **213** | **402** | **74** |
| **Sex** | | | | | | | | | | | | |
| Female | 173 (49.15) | 161 (51.11) | 299 (51.64) | 46 (43.40) | 247 (52.11) | 256 (50.49) | 432 (52.05) | 44 (42.72) | 168 (63.64) | 119 (55.87) | 240 (59.70) | 50 (67.57) |
| Male | 179 (50.85) | 154 (48.89) | 280 (48.36) | 60 (56.60) | 227 (47.89) | 251 (49.51) | 398 (47.95) | 59 (57.28) | 96 (36.36) | 94 (44.13) | 162 (40.30) | 24 (32.43) |
| Missing/Unknown | 0 (0.00) | 0 (0.00) | 0 (0.00) | 0 (0.00) | 0 (0.00) | 0 (0.00) | 0 (0.00) | 0 (0.00) | 0 (0.00) | 0 (0.00) | 0 (0.00) | 0 (0.00) |
| **Race/Ethnicity** | | | | | | | | | | | | |
| Asian | *  (*) | *  (*) | *  (*) | 0 (0.00) | *  (*) | *  (*) | *  (*) | *  (*) | *  (*) | *  (*) | *  (*) | *  (*) |
| Black | 17 (4.83) | *  (*) | 23 (3.97) | *  (*) | 24 (5.06) | 18 (3.55) | 40 (4.82) | *  (*) | *  (*) | *  (*) | 14 (3.48) | *  (*) |
| Hispanic | 0 (0.00) | *  (*) | *  (*) | 0 (0.00) | *  (*) | *  (*) | *  (*) | 0 (0.00) | *  (*) | *  (*) | *  (*) | 0 (0.00) |
| Alaskan Native/Native American | *  (*) | 0 (0.00) | *  (*) | *  (*) | 0 (0.00) | 0 (0.00) | 0 (0.00) | *  (*) | *  (*) | 0 (0.00) | *  (*) | 0 (0.00) |
| White | 302 (85.80) | 276 (87.62) | 501 (86.53) | 91 (85.85) | 423 (89.24) | 466 (91.91) | 753 (90.72) | 91 (88.35) | 240 (90.91) | 203 (95.31) | 369 (91.79) | 64 (86.49) |
| Other | *  (*) | *  (*) | *  (*) | *  (*) | 12 (2.53) | *  (*) | 16 (1.93) | *  (*) | *  (*) | *  (*) | *  (*) | *  (*) |
| Missing/Unknown | 19 (5.40) | 20 (6.35) | 34 (5.87) | *  (*) | *  (*) | *  (*) | *  (*) | *  (*) | 0 (0.00) | 0 (0.00) | 0 (0.00) | 0 (0.00) |
| **Urban/Rural** | | | | | | | | | | | | |
| Urban | 276 (78.41) | 238 (75.56) | 446 (77.03) | 87 (82.08) | 384 (81.01) | 408 (80.47) | 669 (80.60) | 87 (84.47) | 211 (79.92) | 163 (76.53) | 322 (80.10) | 63 (85.14) |
| Rural | *  (*) | *  (*) | *  (*) | 19 (17.92) | *  (*) | 99 (19.53) | *  (*) | 16 (15.53) | *  (*) | 50 (23.47) | *  (*) | *  (*) |
| Missing/Unknown | *  (*) | *  (*) | *  (*) | 0 (0.00) | *  (*) | 0 (0.00) | *  (*) | 0 (0.00) | *  (*) | 0 (0.00) | *  (*) | *  (*) |
| **HHS Region** | | | | | | | | | | | | |
| Region 1 | 23 (6.53) | 35 (11.11) | 50 (8.64) | *  (*) | 41 (8.65) | 27 (5.33) | 62 (7.47) | *  (*) | 17 (6.44) | 20 (9.39) | 30 (7.46) | *  (*) |
| Region 2 | 33 (9.38) | 20 (6.35) | 48 (8.29) | 13 (12.26) | 41 (8.65) | 48 (9.47) | 74 (8.92) | *  (*) | 25 (9.47) | 20 (9.39) | 37 (9.20) | *  (*) |
| Region 3 | 52 (14.77) | 37 (11.75) | 79 (13.64) | 22 (20.75) | 72 (15.19) | 58 (11.44) | 110 (13.25) | 13 (12.62) | 37 (14.02) | 32 (15.02) | 61 (15.17) | 11 (14.86) |
| Region 4 | 58 (16.48) | 71 (22.54) | 106 (18.31) | 19 (17.92) | 102 (21.52) | 104 (20.51) | 172 (20.72) | 15 (14.56) | 51 (19.32) | 35 (16.43) | 76 (18.91) | 14 (18.92) |
| Region 5 | 78 (22.16) | 54 (17.14) | 119 (20.55) | 16 (15.09) | 67 (14.14) | 86 (16.96) | 129 (15.54) | 22 (21.36) | 50 (18.94) | 34 (15.96) | 68 (16.92) | 14 (18.92) |
| Region 6 | 25 (7.10) | 33 (10.48) | 50 (8.64) | *  (*) | 43 (9.07) | 48 (9.47) | 77 (9.28) | 17 (16.50) | 19 (7.20) | 25 (11.74) | 33 (8.21) | *  (*) |
| Region 7 | *  (*) | *  (*) | 22 (3.80) | *  (*) | 19 (4.01) | 25 (4.93) | 35 (4.22) | *  (*) | 12 (4.55) | *  (*) | 16 (3.98) | *  (*) |
| Region 8 | *  (*) | *  (*) | *  (*) | *  (*) | 15 (3.16) | 19 (3.75) | 28 (3.37) | *  (*) | *  (*) | *  (*) | *  (*) | *  (*) |
| Region 9 | 45 (12.78) | 35 (11.11) | 69 (11.92) | 16 (15.09) | 53 (11.18) | 75 (14.79) | 111 (13.37) | 13 (12.62) | 31 (11.74) | 25 (11.74) | 49 (12.19) | *  (*) |
| Region 10 | 18 (5.11) | *  (*) | 25 (4.32) | *  (*) | 21 (4.43) | 17 (3.35) | 32 (3.86) | *  (*) | 15 (5.68) | *  (*) | 23 (5.72) | *  (*) |
| Missing/Unknown | 0 (0.00) | 0 (0.00) | *  (*) | 0 (0.00) | 0 (0.00) | 0 (0.00) | 0 (0.00) | 0 (0.00) | *  (*) | 0 (0.00) | *  (*) | 0 (0.00) |
| **Dual-Eligibility Status**** | | | | | | | | | | | | |
| Dual-Eligible | 14 (3.98) | 11 (3.49) | 23 (3.97) | *  (*) | *  (*) | 14 (2.76) | 18 (2.17) | *  (*) | 11 (4.17) | *  (*) | 16 (3.98) | *  (*) |
| Non-Dual-Eligible | 338 (96.02) | 304 (96.51) | 556 (96.03) | *  (*) | *  (*) | 493 (97.24) | 812 (97.83) | *  (*) | 253 (95.83) | *  (*) | 386 (96.02) | *  (*) |
| **Area Deprivation Index (ADI) Rank** | | | | | | | | | | | | |
| 1-10 (lowest level of deprivation or disadvantage) | 56 (15.91) | 50 (15.87) | 89 (15.37) | 17 (16.04) | 82 (17.30) | 88 (17.36) | 146 (17.59) | 17 (16.50) | 49 (18.56) | 32 (15.02) | 72 (17.91) | 17 (22.97) |
| 11-20 | 53 (15.06) | 52 (16.51) | 93 (16.06) | 17 (16.04) | 76 (16.03) | 80 (15.78) | 131 (15.78) | 18 (17.48) | 35 (13.26) | 25 (11.74) | 55 (13.68) | *  (*) |
| 21-30 | 50 (14.20) | 43 (13.65) | 81 (13.99) | 12 (11.32) | 75 (15.82) | 80 (15.78) | 130 (15.66) | 14 (13.59) | 33 (12.50) | 32 (15.02) | 57 (14.18) | *  (*) |
| 31-40 | 46 (13.07) | 37 (11.75) | 75 (12.95) | 16 (15.09) | 53 (11.18) | 57 (11.24) | 92 (11.08) | *  (*) | 31 (11.74) | 21 (9.86) | 45 (11.19) | 13 (17.57) |
| 41-50 | 32 (9.09) | 35 (11.11) | 57 (9.84) | 12 (11.32) | 48 (10.13) | 51 (10.06) | 86 (10.36) | 13 (12.62) | 29 (10.98) | 28 (13.15) | 44 (10.95) | *  (*) |
| 51-60 | 33 (9.38) | 31 (9.84) | 56 (9.67) | *  (*) | 37 (7.81) | 38 (7.50) | 63 (7.59) | 13 (12.62) | 18 (6.82) | 24 (11.27) | 28 (6.97) | *  (*) |
| 61-70 | 23 (6.53) | 16 (5.08) | 35 (6.04) | *  (*) | 28 (5.91) | 35 (6.90) | 50 (6.02) | *  (*) | 20 (7.58) | 21 (9.86) | 32 (7.96) | *  (*) |
| 71-80 | 25 (7.10) | 18 (5.71) | 39 (6.74) | *  (*) | 29 (6.12) | 32 (6.31) | 54 (6.51) | *  (*) | 14 (5.30) | *  (*) | 20 (4.98) | *  (*) |
| 81-90 | 17 (4.83) | 14 (4.44) | 24 (4.15) | *  (*) | 16 (3.38) | 22 (4.34) | 33 (3.98) | *  (*) | 15 (5.68) | 15 (7.04) | 25 (6.22) | *  (*) |
| 91-100 | *  (*) | *  (*) | 14 (2.42) | *  (*) | 14 (2.95) | 12 (2.37) | 22 (2.65) | *  (*) | *  (*) | *  (*) | 12 (2.99) | *  (*) |
| Missing/Unknown | *  (*) | *  (*) | 16 (2.76) | *  (*) | 16 (3.38) | 12 (2.37) | 23 (2.77) | *  (*) | *  (*) | *  (*) | 12 (2.99) | 0 (0.00) |
| **Medicare Status** | | | | | | | | | | | | |
| Aged-in without ESRD | 305 (86.65) | 287 (91.11) | 515 (88.95) | 96 (90.57) | 451 (95.15) | 481 (94.87) | 787 (94.82) | 98 (95.15) | 260 (98.48) | 208 (97.65) | 395 (98.26) | 73 (98.65) |
| Aged & Disabled with ESRD | 0 (0.00) | 0 (0.00) | 0 (0.00) | 0 (0.00) | 0 (0.00) | 0 (0.00) | 0 (0.00) | 0 (0.00) | 0 (0.00) | 0 (0.00) | 0 (0.00) | 0 (0.00) |
| Disabled without ESRD | 47 (13.35) | 28 (8.89) | 64 (11.05) | *  (*) | 23 (4.85) | 26 (5.13) | 43 (5.18) | *  (*) | *  (*) | *  (*) | *  (*) | *  (*) |
| Missing/Unknown | 0 (0.00) | 0 (0.00) | 0 (0.00) | *  (*) | 0 (0.00) | 0 (0.00) | 0 (0.00) | *  (*) | *  (*) | *  (*) | *  (*) | *  (*) |
| **Medical Conditions (0-365 days prior to vaccination date)** | | | | | | | | | | | | |
| Asthma | 23 (6.53) | 36 (11.43) | 53 (9.15) | *  (*) | 32 (6.75) | 42 (8.28) | 58 (6.99) | 12 (11.65) | *  (*) | 17 (7.98) | 24 (5.97) | *  (*) |
| COPD | 39 (11.08) | 30 (9.52) | 61 (10.54) | 17 (16.04) | 55 (11.60) | 61 (12.03) | 95 (11.45) | 13 (12.62) | 28 (10.61) | 17 (7.98) | 38 (9.45) | *  (*) |
| Chronic Kidney Disease | 81 (23.01) | 46 (14.60) | 114 (19.69) | 22 (20.75) | 142 (29.96) | 122 (24.06) | 227 (27.35) | 28 (27.18) | 87 (32.95) | 68 (31.92) | 132 (32.84) | 26 (35.14) |
| Depression | 66 (18.75) | 56 (17.78) | 106 (18.31) | 21 (19.81) | 79 (16.67) | 102 (20.12) | 151 (18.19) | 19 (18.45) | 34 (12.88) | 25 (11.74) | 50 (12.44) | *  (*) |
| Gout | 19 (5.40) | 15 (4.76) | 31 (5.35) | *  (*) | 37 (7.81) | 40 (7.89) | 65 (7.83) | 14 (13.59) | 12 (4.55) | 11 (5.16) | 18 (4.48) | *  (*) |
| Heart Failure | 34 (9.66) | 23 (7.30) | 51 (8.81) | 15 (14.15) | 72 (15.19) | 60 (11.83) | 108 (13.01) | 20 (19.42) | 55 (20.83) | 38 (17.84) | 79 (19.65) | 28 (37.84) |
| Hypercholesterolemia | 64 (18.18) | 75 (23.81) | 123 (21.24) | 26 (24.53) | 109 (23.00) | 114 (22.49) | 184 (22.17) | 23 (22.33) | 48 (18.18) | 52 (24.41) | 82 (20.40) | 16 (21.62) |
| Hypothyroidism | 257 (73.01) | 211 (66.98) | 411 (70.98) | 76 (71.70) | 377 (79.54) | 387 (76.33) | 652 (78.55) | 85 (82.52) | 208 (78.79) | 175 (82.16) | 321 (79.85) | 66 (89.19) |
| Hypertension | 68 (19.32) | 66 (20.95) | 122 (21.07) | 22 (20.75) | 100 (21.10) | 137 (27.02) | 196 (23.61) | 27 (26.21) | 75 (28.41) | 52 (24.41) | 107 (26.62) | 23 (31.08) |
| ITP | *  (*) | *  (*) | *  (*) | *  (*) | *  (*) | *  (*) | *  (*) | *  (*) | *  (*) | *  (*) | *  (*) | *  (*) |
| Impaired Mobility | *  (*) | *  (*) | *  (*) | *  (*) | *  (*) | *  (*) | *  (*) | *  (*) | *  (*) | *  (*) | *  (*) | *  (*) |
| Ischemic Heart Disease | 16 (4.55) | 19 (6.03) | 33 (5.70) | *  (*) | 24 (5.06) | 36 (7.10) | 47 (5.66) | *  (*) | 13 (4.92) | 18 (8.45) | 26 (6.47) | *  (*) |
| Nicotine Dependency | 83 (23.58) | 57 (18.10) | 127 (21.93) | 30 (28.30) | 95 (20.04) | 118 (23.27) | 174 (20.96) | 32 (31.07) | 43 (16.29) | 34 (15.96) | 61 (15.17) | 13 (17.57) |
| Obesity | 101 (28.69) | 77 (24.44) | 154 (26.60) | 30 (28.30) | 113 (23.84) | 112 (22.09) | 188 (22.65) | 25 (24.27) | 30 (11.36) | 29 (13.62) | 52 (12.94) | *  (*) |
| **Charlson Comorbidity Index** | | | | | | | | | | | | |
| 0 | 104 (29.55) | 119 (37.78) | 189 (32.64) | 22 (20.75) | 108 (22.78) | 128 (25.25) | 199 (23.98) | 22 (21.36) | 55 (20.83) | 47 (22.07) | 82 (20.40) | 11 (14.86) |
| 1 | 54 (15.34) | 64 (20.32) | 101 (17.44) | 16 (15.09) | 84 (17.72) | 96 (18.93) | 152 (18.31) | 18 (17.48) | 50 (18.94) | 38 (17.84) | 76 (18.91) | 14 (18.92) |
| 2 | 64 (18.18) | 49 (15.56) | 104 (17.96) | 20 (18.87) | 83 (17.51) | 88 (17.36) | 148 (17.83) | *  (*) | 42 (15.91) | 34 (15.96) | 64 (15.92) | *  (*) |
| 3 | 36 (10.23) | 24 (7.62) | 49 (8.46) | *  (*) | 61 (12.87) | 59 (11.64) | 103 (12.41) | *  (*) | 40 (15.15) | 30 (14.08) | 61 (15.17) | 19 (25.68) |
| 4 | 34 (9.66) | 22 (6.98) | 51 (8.81) | *  (*) | 37 (7.81) | 55 (10.85) | 79 (9.52) | 18 (17.48) | 30 (11.36) | 23 (10.80) | 46 (11.44) | *  (*) |
| 5+ | 60 (17.05) | 37 (11.75) | 85 (14.68) | 27 (25.47) | 101 (21.31) | 81 (15.98) | 149 (17.95) | 20 (19.42) | 47 (17.80) | 41 (19.25) | 73 (18.16) | 13 (17.57) |
| **Prior COVID-19 diagnosis** | | | | | | | | | | | | |
| In the 30 days prior to outcome | 0 (0.00) | 0 (0.00) | 0 (0.00) | 0 (0.00) | 0 (0.00) | 0 (0.00) | 0 (0.00) | 0 (0.00) | 0 (0.00) | 0 (0.00) | 0 (0.00) | *  (*) |
| In the 31-365 days prior to outcome | 41 (11.65) | 51 (16.19) | 77 (13.30) | 14 (13.21) | 51 (10.76) | 73 (14.40) | 106 (12.77) | 18 (17.48) | 20 (7.58) | 15 (7.04) | 31 (7.71) | *  (*) |
| **Concomitant Influenza Vaccination (same day as bivalent booster)** | | | | | | | | | | | | |
| High-dose | 97 (27.56) | 79 (25.08) | 161 (27.81) | 24 (22.64) | 106 (22.36) | 108 (21.30) | 189 (22.77) | 23 (22.33) | 65 (24.62) | 41 (19.25) | 90 (22.39) | 17 (22.97) |
| Adjuvanted | 50 (14.20) | 44 (13.97) | 84 (14.51) | 14 (13.21) | 69 (14.56) | 64 (12.62) | 116 (13.98) | *  (*) | 45 (17.05) | 26 (12.21) | 61 (15.17) | 12 (16.22) |
| Recombinant | *  (*) | *  (*) | *  (*) | 0 (0.00) | *  (*) | 0 (0.00) | *  (*) | *  (*) | 0 (0.00) | 0 (0.00) | 0 (0.00) | *  (*) |
| Live-attenuated | 0 (0.00) | 0 (0.00) | 0 (0.00) | 0 (0.00) | 0 (0.00) | 0 (0.00) | 0 (0.00) | 0 (0.00) | 0 (0.00) | 0 (0.00) | 0 (0.00) | 0 (0.00) |
| Cell-cultured | 0 (0.00) | 0 (0.00) | 0 (0.00) | *  (*) | *  (*) | *  (*) | *  (*) | *  (*) | *  (*) | 0 (0.00) | *  (*) | *  (*) |
| Standard | *  (*) | *  (*) | *  (*) | *  (*) | *  (*) | *  (*) | *  (*) | 0 (0.00) | *  (*) | *  (*) | *  (*) | 0 (0.00) |
| **Other vaccine (administered in risk/control intervals)** | | | | | | | | | | | | |
| Pneumococcal vaccine | 18 (5.11) | 15 (4.76) | 29 (5.01) | *  (*) | 16 (3.38) | 16 (3.16) | 28 (3.37) | 0 (0.00) | 11 (4.17) | *  (*) | 14 (3.48) | *  (*) |

**Abbreviations:** BNT162b2; WT/OMI BA.4/BA.5, Pfizer COVID-19 BivalentmRNA-1273.222, Moderna COVID-19 Bivalent;

HS, hemorrhagic stroke; NHS, non-hemorrhagic stroke; TIA, transient ischemic attacks; ADI, area deprivations index; ESRD, end-stage renal disease; COPD, chronic obstructive pulmonary disease; COVID-19, coronavirus 2019

* Outcome counts of 10 or fewer and associated statistics are masked to protect the anonymity of the data.

** Reflects beneficiaries’ dual eligibility for Medicare and Medicaid plan benefits.

**eTable 11. Summary of Demographics, Socio-Economic Status, Residence, Health Status, and Healthcare Utilization of BNT162b2; WT/OMI BA.4/BA.5 Bivalent Vaccinated Medicare Beneficiaries with Stroke Outcomes in the Primary Analysis, by Concomitant Status**

| **Patient Characteristics** | **BNT162b2; WT/OMI BA.4/BA.5** | | | | | | | |
| --- | --- | --- | --- | --- | --- | --- | --- | --- |
|  | **Concomitant Influenza** | | | | **No Concomitant** | | | |
|  | **NHS**  n (%) | **TIA**  n (%) | **NHS/ TIA**  n (%) | **HS**  n (%) | **NHS**  n (%) | **TIA**  n (%) | **NHS/ TIA**  n (%) | **HS**  n (%) |
| **Total** | **797** | **704** | **1,329** | **245** | **967** | **876** | **1,601** | **271** |
| **Age** | | | | | | | | |
| 65-74 | 260 (32.62) | 251 (35.65) | 449 (33.78) | 81 (33.06) | 314 (32.47) | 235 (26.83) | 502 (31.36) | 97 (35.79) |
| 75-84 | 324 (40.65) | 282 (40.06) | 536 (40.33) | 108 (44.08) | 394 (40.74) | 418 (47.72) | 696 (43.47) | 108 (39.85) |
| 85+ | 213 (26.73) | 171 (24.29) | 344 (25.88) | 56 (22.86) | 259 (26.78) | 223 (25.46) | 403 (25.17) | 66 (24.35) |
| **Sex** | | | | | | | | |
| Female | 421 (52.82) | 401 (56.96) | 727 (54.70) | 109 (44.49) | 554 (57.29) | 529 (60.39) | 934 (58.34) | 136 (50.18) |
| Male | 376 (47.18) | 303 (43.04) | 602 (45.30) | 136 (55.51) | 413 (42.71) | 347 (39.61) | 667 (41.66) | 135 (49.82) |
| Missing/Unknown | 0 (0.00) | 0 (0.00) | 0 (0.00) | 0 (0.00) | 0 (0.00) | 0 (0.00) | 0 (0.00) | 0 (0.00) |
| **Race/Ethnicity** | | | | | | | | |
| Asian | 13 (1.63) | *  (*) | 18 (1.35) | *  (*) | 25 (2.59) | 13 (1.48) | 36 (2.25) | *  (*) |
| Black | 48 (6.02) | 28 (3.98) | 63 (4.74) | 11 (4.49) | 61 (6.31) | 40 (4.57) | 85 (5.31) | *  (*) |
| Hispanic | *  (*) | *  (*) | *  (*) | *  (*) | *  (*) | *  (*) | *  (*) | 0 (0.00) |
| Alaskan Native/Native American | *  (*) | *  (*) | *  (*) | 0 (0.00) | *  (*) | *  (*) | *  (*) | 0 (0.00) |
| White | 691 (86.70) | 644 (91.48) | 1,183 (89.01) | 219 (89.39) | 835 (86.35) | 782 (89.27) | 1,403 (87.63) | 240 (88.56) |
| Other | 14 (1.76) | *  (*) | 20 (1.50) | *  (*) | 22 (2.28) | 14 (1.60) | 32 (2.00) | *  (*) |
| Missing/Unknown | 21 (2.63) | 17 (2.41) | 33 (2.48) | *  (*) | 19 (1.96) | 21 (2.40) | 36 (2.25) | *  (*) |
| **Urban/Rural** | | | | | | | | |
| Urban | 677 (84.94) | 586 (83.24) | 1,119 (84.20) | 217 (88.57) | 862 (89.14) | 745 (85.05) | 1,402 (87.57) | 232 (85.61) |
| Rural | *  (*) | *  (*) | *  (*) | 28 (11.43) | *  (*) | *  (*) | *  (*) | *  (*) |
| Missing/Unknown | *  (*) | *  (*) | *  (*) | 0 (0.00) | *  (*) | *  (*) | *  (*) | *  (*) |
| **HHS Region** | | | | | | | | |
| Region 1 | 58 (7.28) | 64 (9.09) | 104 (7.83) | *  (*) | 91 (9.41) | 70 (7.99) | 142 (8.87) | 22 (8.12) |
| Region 2 | 62 (7.78) | 34 (4.83) | 84 (6.32) | 20 (8.16) | 89 (9.20) | 92 (10.50) | 156 (9.74) | 30 (11.07) |
| Region 3 | 113 (14.18) | 77 (10.94) | 172 (12.94) | 32 (13.06) | 135 (13.96) | 116 (13.24) | 214 (13.37) | 34 (12.55) |
| Region 4 | 118 (14.81) | 129 (18.32) | 222 (16.70) | 36 (14.69) | 177 (18.30) | 161 (18.38) | 298 (18.61) | 48 (17.71) |
| Region 5 | 173 (21.71) | 150 (21.31) | 287 (21.60) | 54 (22.04) | 206 (21.30) | 144 (16.44) | 309 (19.30) | 56 (20.66) |
| Region 6 | 70 (8.78) | 52 (7.39) | 105 (7.90) | 19 (7.76) | 62 (6.41) | 66 (7.53) | 110 (6.87) | 17 (6.27) |
| Region 7 | 55 (6.90) | 49 (6.96) | 89 (6.70) | 12 (4.90) | 44 (4.55) | 36 (4.11) | 72 (4.50) | *  (*) |
| Region 8 | 30 (3.76) | *  (*) | *  (*) | 14 (5.71) | 33 (3.41) | 35 (4.00) | *  (*) | *  (*) |
| Region 9 | 72 (9.03) | 67 (9.52) | 128 (9.63) | 35 (14.29) | 96 (9.93) | 116 (13.24) | 176 (10.99) | 38 (14.02) |
| Region 10 | 46 (5.77) | 45 (6.39) | 81 (6.09) | 13 (5.31) | *  (*) | 40 (4.57) | 64 (4.00) | *  (*) |
| Missing/Unknown | 0 (0.00) | *  (*) | *  (*) | *  (*) | *  (*) | 0 (0.00) | *  (*) | 0 (0.00) |
| **Dual-Eligibility Status**** | | | | | | | | |
| Dual-Eligible | 29 (3.64) | 29 (4.12) | 44 (3.31) | *  (*) | 32 (3.31) | 22 (2.51) | 45 (2.81) | 12 (4.43) |
| Non-Dual-Eligible | 768 (96.36) | 675 (95.88) | 1,285 (96.69) | *  (*) | 935 (96.69) | 854 (97.49) | 1,556 (97.19) | 259 (95.57) |
| **Area Deprivation Index (ADI) Rank** | | | | | | | | |
| 1-10 (lowest level of deprivation or disadvantage) | 120 (15.06) | 109 (15.48) | 205 (15.43) | 45 (18.37) | 182 (18.82) | 171 (19.52) | 315 (19.68) | 52 (19.19) |
| 11-20 | 119 (14.93) | 101 (14.35) | 194 (14.60) | 46 (18.78) | 155 (16.03) | 151 (17.24) | 267 (16.68) | 38 (14.02) |
| 21-30 | 120 (15.06) | 101 (14.35) | 202 (15.20) | 38 (15.51) | 139 (14.37) | 114 (13.01) | 221 (13.80) | 50 (18.45) |
| 31-40 | 111 (13.93) | 94 (13.35) | 179 (13.47) | 25 (10.20) | 129 (13.34) | 103 (11.76) | 197 (12.30) | 42 (15.50) |
| 41-50 | 94 (11.79) | 83 (11.79) | 155 (11.66) | 26 (10.61) | 92 (9.51) | 84 (9.59) | 153 (9.56) | 24 (8.86) |
| 51-60 | 78 (9.79) | 79 (11.22) | 137 (10.31) | 32 (13.06) | 71 (7.34) | 74 (8.45) | 119 (7.43) | 26 (9.59) |
| 61-70 | 61 (7.65) | 47 (6.68) | 93 (7.00) | 12 (4.90) | 72 (7.45) | 58 (6.62) | 115 (7.18) | 13 (4.80) |
| 71-80 | 38 (4.77) | 38 (5.40) | 71 (5.34) | *  (*) | 47 (4.86) | 50 (5.71) | 84 (5.25) | *  (*) |
| 81-90 | 18 (2.26) | 28 (3.98) | 39 (2.93) | *  (*) | 33 (3.41) | 28 (3.20) | 51 (3.19) | *  (*) |
| 91-100 | 19 (2.38) | *  (*) | 24 (1.81) | *  (*) | 25 (2.59) | 20 (2.28) | 41 (2.56) | *  (*) |
| Missing/Unknown | 19 (2.38) | *  (*) | 30 (2.26) | *  (*) | 22 (2.28) | 23 (2.63) | 38 (2.37) | *  (*) |
| **Medicare Status** | | | | | | | | |
| Aged-in without ESRD | 732 (91.84) | 662 (94.03) | 1,235 (92.93) | 226 (92.24) | 911 (94.21) | 835 (95.32) | 1,518 (94.82) | 246 (90.77) |
| Aged & Disabled with ESRD | 0 (0.00) | *  (*) | *  (*) | *  (*) | *  (*) | 0 (0.00) | *  (*) | *  (*) |
| Disabled without ESRD | 65 (8.16) | 39 (5.54) | 93 (7.00) | 18 (7.35) | 55 (5.69) | 41 (4.68) | 82 (5.12) | 24 (8.86) |
| Missing/Unknown | 0 (0.00) | *  (*) | *  (*) | *  (*) | *  (*) | 0 (0.00) | *  (*) | *  (*) |
| **Medical Conditions (0-365 days prior to vaccination date)** | | | | | | | | |
| Asthma | 49 (6.15) | 60 (8.52) | 100 (7.52) | *  (*) | 76 (7.86) | 73 (8.33) | 136 (8.49) | 24 (8.86) |
| COPD | 92 (11.54) | 71 (10.09) | 146 (10.99) | 32 (13.06) | 117 (12.10) | 93 (10.62) | 182 (11.37) | 37 (13.65) |
| Chronic Kidney Disease | 226 (28.36) | 175 (24.86) | 348 (26.19) | 72 (29.39) | 240 (24.82) | 233 (26.60) | 391 (24.42) | 79 (29.15) |
| Depression | 139 (17.44) | 136 (19.32) | 245 (18.43) | 40 (16.33) | 164 (16.96) | 165 (18.84) | 288 (17.99) | 43 (15.87) |
| Gout | 49 (6.15) | 46 (6.53) | 85 (6.40) | 13 (5.31) | 61 (6.31) | 58 (6.62) | 99 (6.18) | 16 (5.90) |
| Heart Failure | 106 (13.30) | 90 (12.78) | 168 (12.64) | 50 (20.41) | 125 (12.93) | 103 (11.76) | 192 (11.99) | 53 (19.56) |
| Hypercholesterolemia | 146 (18.32) | 141 (20.03) | 250 (18.81) | 62 (25.31) | 210 (21.72) | 216 (24.66) | 360 (22.49) | 59 (21.77) |
| Hypothyroidism | 618 (77.54) | 532 (75.57) | 1,014 (76.30) | 200 (81.63) | 735 (76.01) | 642 (73.29) | 1,184 (73.95) | 209 (77.12) |
| Hypertension | 176 (22.08) | 178 (25.28) | 310 (23.33) | 46 (18.78) | 242 (25.03) | 251 (28.65) | 414 (25.86) | 68 (25.09) |
| ITP | *  (*) | *  (*) | *  (*) | *  (*) | *  (*) | *  (*) | *  (*) | *  (*) |
| Impaired Mobility | *  (*) | *  (*) | *  (*) | *  (*) | *  (*) | *  (*) | *  (*) | *  (*) |
| Ischemic Heart Disease | 33 (4.14) | 30 (4.26) | 55 (4.14) | 18 (7.35) | 49 (5.07) | 35 (4.00) | 71 (4.43) | 18 (6.64) |
| Nicotine Dependency | 190 (23.84) | 152 (21.59) | 290 (21.82) | 66 (26.94) | 214 (22.13) | 169 (19.29) | 334 (20.86) | 74 (27.31) |
| Obesity | 174 (21.83) | 143 (20.31) | 282 (21.22) | 65 (26.53) | 191 (19.75) | 188 (21.46) | 329 (20.55) | 61 (22.51) |
| **Charlson Comorbidity Index** | | | | | | | | |
| 0 | 192 (24.09) | 179 (25.43) | 336 (25.28) | 54 (22.04) | 248 (25.65) | 198 (22.60) | 400 (24.98) | 54 (19.93) |
| 1 | 143 (17.94) | 121 (17.19) | 229 (17.23) | 40 (16.33) | 172 (17.79) | 151 (17.24) | 287 (17.93) | 38 (14.02) |
| 2 | 146 (18.32) | 139 (19.74) | 257 (19.34) | 40 (16.33) | 148 (15.31) | 163 (18.61) | 265 (16.55) | 40 (14.76) |
| 3 | 91 (11.42) | 98 (13.92) | 162 (12.19) | 25 (10.20) | 125 (12.93) | 128 (14.61) | 218 (13.62) | 43 (15.87) |
| 4 | 91 (11.42) | 53 (7.53) | 130 (9.78) | 30 (12.24) | 99 (10.24) | 94 (10.73) | 163 (10.18) | 27 (9.96) |
| 5+ | 134 (16.81) | 114 (16.19) | 215 (16.18) | 56 (22.86) | 175 (18.10) | 142 (16.21) | 268 (16.74) | 69 (25.46) |
| **Prior COVID-19 diagnosis** | | | | | | | | |
| In the 30 days prior to outcome | 0 (0.00) | 0 (0.00) | 0 (0.00) | 0 (0.00) | 0 (0.00) | 0 (0.00) | 0 (0.00) | 0 (0.00) |
| In the 31-365 days prior to outcome | 77 (9.66) | 98 (13.92) | 148 (11.14) | 27 (11.02) | 110 (11.38) | 134 (15.30) | 202 (12.62) | 33 (12.18) |
| **Other vaccine (administered in risk/control intervals)** | | | | | | | | |
| Pneumococcal vaccine | 20 (2.51) | 22 (3.12) | 39 (2.93) | *  (*) | 26 (2.69) | 23 (2.63) | 43 (2.69) | *  (*) |

**Abbreviations:** BNT162b2; WT/OMI BA.4/BA.5, Pfizer COVID-19 BivalentmRNA-1273.222, Moderna COVID-19 Bivalent;

HS, hemorrhagic stroke; NHS, non-hemorrhagic stroke; TIA, transient ischemic attacks; ADI, area deprivations index; ESRD, end-stage renal disease; COPD, chronic obstructive pulmonary disease; COVID-19, coronavirus 2019

* Outcome counts of 10 or fewer and associated statistics are masked to protect the anonymity of the data.

** Reflects beneficiaries’ dual eligibility for Medicare and Medicaid plan benefits.

**eTable 12. Summary of Demographics, Socio-Economic Status, Residence, Health Status, and Healthcare Utilization of MRNA-1273.222 Bivalent Vaccinated Medicare Beneficaries with Stroke Outcomes in the Primary Analysis, by Concomitant Status**

| **Patient Characteristics** | **MRNA-1273.222** | | | | | | | |
| --- | --- | --- | --- | --- | --- | --- | --- | --- |
|  | **Concomitant Influenza** | | | | **No Concomitant** | | | |
|  | **NHS**  n (%) | **TIA**  n (%) | **NHS/ TIA**  n (%) | **HS**  n (%) | **NHS**  n (%) | **TIA**  n (%) | **NHS/ TIA**  n (%) | **HS**  n (%) |
| **Total** | **432** | **362** | **701** | **98** | **643** | **664** | **1090** | **181** |
| **Age** | | | | | | | | |
| 65-74 | 147 (34.03) | 123 (33.98) | 245 (34.95) | 38 (38.78) | 201 (31.26) | 189 (28.46) | 327 (30.00) | 67 (37.02) |
| 75-84 | 175 (40.51) | 172 (47.51) | 305 (43.51) | 31 (31.63) | 290 (45.10) | 330 (49.70) | 515 (47.25) | 70 (38.67) |
| 85+ | 110 (25.46) | 67 (18.51) | 151 (21.54) | 29 (29.59) | 152 (23.64) | 145 (21.84) | 248 (22.75) | 44 (24.31) |
| **Sex** | | | | | | | | |
| Female | 247 (57.18) | 191 (52.76) | 395 (56.35) | 41 (41.84) | 335 (52.10) | 339 (51.05) | 567 (52.02) | 97 (53.59) |
| Male | 185 (42.82) | 171 (47.24) | 306 (43.65) | 57 (58.16) | 308 (47.90) | 325 (48.95) | 523 (47.98) | 84 (46.41) |
| Missing/Unknown | 0 (0.00) | 0 (0.00) | 0 (0.00) | 0 (0.00) | 0 (0.00) | 0 (0.00) | 0 (0.00) | 0 (0.00) |
| **Race/Ethnicity** | | | | | | | | |
| Asian | *  (*) | *  (*) | 12 (1.71) | 0 (0.00) | *  (*) | *  (*) | 12 (1.10) | *  (*) |
| Black | 17 (3.94) | *  (*) | 25 (3.57) | 0 (0.00) | 34 (5.29) | 19 (2.86) | 52 (4.77) | *  (*) |
| Hispanic | *  (*) | *  (*) | *  (*) | 0 (0.00) | *  (*) | *  (*) | *  (*) | 0 (0.00) |
| Alaskan Native/Native American | 0 (0.00) | 0 (0.00) | 0 (0.00) | *  (*) | *  (*) | 0 (0.00) | *  (*) | *  (*) |
| White | 389 (90.05) | 333 (91.99) | 635 (90.58) | 88 (89.80) | 562 (87.40) | 604 (90.96) | 970 (88.99) | 154 (85.08) |
| Other | *  (*) | *  (*) | *  (*) | *  (*) | 19 (2.95) | *  (*) | 25 (2.29) | *  (*) |
| Missing/Unknown | *  (*) | *  (*) | 17 (2.43) | *  (*) | 16 (2.49) | 18 (2.71) | 26 (2.39) | *  (*) |
| **Urban/Rural** | | | | | | | | |
| Urban | 340 (78.70) | 273 (75.41) | 547 (78.03) | 81 (82.65) | 521 (81.03) | 529 (79.67) | 875 (80.28) | 152 (83.98) |
| Rural | *  (*) | *  (*) | *  (*) | 17 (17.35) | *  (*) | *  (*) | *  (*) | 29 (16.02) |
| Missing/Unknown | *  (*) | *  (*) | *  (*) | 0 (0.00) | *  (*) | *  (*) | *  (*) | 0 (0.00) |
| **HHS Region** | | | | | | | | |
| Region 1 | 40 (9.26) | 33 (9.12) | 66 (9.42) | *  (*) | 41 (6.38) | 48 (7.23) | 75 (6.88) | 12 (6.63) |
| Region 2 | 29 (6.71) | 25 (6.91) | 48 (6.85) | *  (*) | 69 (10.73) | 63 (9.49) | 110 (10.09) | 19 (10.50) |
| Region 3 | 56 (12.96) | 39 (10.77) | 84 (11.98) | 19 (19.39) | 104 (16.17) | 87 (13.10) | 165 (15.14) | 26 (14.36) |
| Region 4 | 70 (16.20) | 68 (18.78) | 117 (16.69) | *  (*) | 136 (21.15) | 140 (21.08) | 232 (21.28) | 36 (19.89) |
| Region 5 | 88 (20.37) | 65 (17.96) | 135 (19.26) | 22 (22.45) | 105 (16.33) | 108 (16.27) | 179 (16.42) | 28 (15.47) |
| Region 6 | 40 (9.26) | 45 (12.43) | 72 (10.27) | *  (*) | 45 (7.00) | 60 (9.04) | 85 (7.80) | 22 (12.15) |
| Region 7 | 15 (3.47) | 18 (4.97) | 29 (4.14) | *  (*) | 29 (4.51) | 24 (3.61) | 44 (4.04) | *  (*) |
| Region 8 | 15 (3.47) | *  (*) | 19 (2.71) | *  (*) | *  (*) | 26 (3.92) | *  (*) | *  (*) |
| Region 9 | 50 (11.57) | 43 (11.88) | 90 (12.84) | 14 (14.29) | 76 (11.82) | 89 (13.40) | 133 (12.20) | 25 (13.81) |
| Region 10 | 29 (6.71) | *  (*) | 41 (5.85) | *  (*) | 24 (3.73) | 19 (2.86) | 38 (3.49) | *  (*) |
| Missing/Unknown | 0 (0.00) | 0 (0.00) | 0 (0.00) | 0 (0.00) | *  (*) | 0 (0.00) | *  (*) | 0 (0.00) |
| **Dual-Eligibility Status**** | | | | | | | | |
| Dual-Eligible | 14 (3.24) | 11 (3.04) | 22 (3.14) | *  (*) | 18 (2.80) | 23 (3.46) | 34 (3.12) | *  (*) |
| Non-Dual-Eligible | 418 (96.76) | 351 (96.96) | 679 (96.86) | *  (*) | 625 (97.20) | 641 (96.54) | 1,056 (96.88) | *  (*) |
| **Area Deprivation Index (ADI) Rank** | | | | | | | | |
| 1-10 (lowest level of deprivation or disadvantage) | 60 (13.89) | 47 (12.98) | 100 (14.27) | 23 (23.47) | 126 (19.60) | 121 (18.22) | 204 (18.72) | 28 (15.47) |
| 11-20 | 70 (16.20) | 56 (15.47) | 115 (16.41) | 19 (19.39) | 94 (14.62) | 100 (15.06) | 163 (14.95) | 23 (12.71) |
| 21-30 | 65 (15.05) | 59 (16.30) | 105 (14.98) | 11 (11.22) | 92 (14.31) | 95 (14.31) | 161 (14.77) | 23 (12.71) |
| 31-40 | 59 (13.66) | 34 (9.39) | 86 (12.27) | 12 (12.24) | 68 (10.58) | 80 (12.05) | 123 (11.28) | 26 (14.36) |
| 41-50 | 43 (9.95) | 39 (10.77) | 72 (10.27) | 12 (12.24) | 63 (9.80) | 75 (11.30) | 112 (10.28) | 20 (11.05) |
| 51-60 | 41 (9.49) | 35 (9.67) | 66 (9.42) | *  (*) | 45 (7.00) | 58 (8.73) | 79 (7.25) | 13 (7.18) |
| 61-70 | 27 (6.25) | 29 (8.01) | 48 (6.85) | *  (*) | 44 (6.84) | 42 (6.33) | 69 (6.33) | *  (*) |
| 71-80 | 27 (6.25) | 24 (6.63) | 46 (6.56) | *  (*) | 40 (6.22) | 35 (5.27) | 66 (6.06) | 16 (8.84) |
| 81-90 | *  (*) | 21 (5.80) | 29 (4.14) | *  (*) | 31 (4.82) | 29 (4.37) | 51 (4.68) | 12 (6.63) |
| 91-100 | *  (*) | *  (*) | 14 (2.00) | *  (*) | 22 (3.42) | 12 (1.81) | 32 (2.94) | *  (*) |
| Missing/Unknown | 17 (3.94) | *  (*) | 20 (2.85) | 0 (0.00) | 18 (2.80) | 17 (2.56) | 30 (2.75) | *  (*) |
| **Medicare Status** | | | | | | | | |
| Aged-in without ESRD | 410 (94.91) | 338 (93.37) | 662 (94.44) | 92 (93.88) | 593 (92.22) | 629 (94.73) | 1,017 (93.30) | 172 (95.03) |
| Aged & Disabled with ESRD | 0 (0.00) | 0 (0.00) | 0 (0.00) | 0 (0.00) | 0 (0.00) | 0 (0.00) | 0 (0.00) | 0 (0.00) |
| Disabled without ESRD | 22 (5.09) | 24 (6.63) | 39 (5.56) | *  (*) | 50 (7.78) | 35 (5.27) | 73 (6.70) | *  (*) |
| Missing/Unknown | 0 (0.00) | 0 (0.00) | 0 (0.00) | *  (*) | 0 (0.00) | 0 (0.00) | 0 (0.00) | *  (*) |
| **Medical Conditions (0-365 days prior to vaccination date)** | | | | | | | | |
| Asthma | 25 (5.79) | 30 (8.29) | 50 (7.13) | *  (*) | 39 (6.07) | 64 (9.64) | 83 (7.61) | 18 (9.94) |
| COPD | 52 (12.04) | 36 (9.94) | 74 (10.56) | *  (*) | 68 (10.58) | 71 (10.69) | 117 (10.73) | 23 (12.71) |
| Chronic Kidney Disease | 108 (25.00) | 76 (20.99) | 165 (23.54) | 22 (22.45) | 195 (30.33) | 160 (24.10) | 301 (27.61) | 53 (29.28) |
| Depression | 79 (18.29) | 70 (19.34) | 133 (18.97) | 22 (22.45) | 97 (15.09) | 113 (17.02) | 171 (15.69) | 26 (14.36) |
| Gout | 28 (6.48) | 20 (5.52) | 43 (6.13) | *  (*) | 39 (6.07) | 45 (6.78) | 69 (6.33) | 15 (8.29) |
| Heart Failure | 59 (13.66) | 43 (11.88) | 87 (12.41) | 26 (26.53) | 99 (15.40) | 77 (11.60) | 147 (13.49) | 37 (20.44) |
| Hypercholesterolemia | 95 (21.99) | 78 (21.55) | 152 (21.68) | 23 (23.47) | 126 (19.60) | 159 (23.95) | 235 (21.56) | 40 (22.10) |
| Hypothyroidism | 330 (76.39) | 278 (76.80) | 537 (76.60) | 78 (79.59) | 499 (77.60) | 489 (73.64) | 830 (76.15) | 145 (80.11) |
| Hypertension | 106 (24.54) | 95 (26.24) | 179 (25.53) | 15 (15.31) | 135 (21.00) | 159 (23.95) | 243 (22.29) | 56 (30.94) |
| ITP | *  (*) | 0 (0.00) | *  (*) | 0 (0.00) | *  (*) | *  (*) | *  (*) | *  (*) |
| Impaired Mobility | *  (*) | 0 (0.00) | *  (*) | *  (*) | *  (*) | *  (*) | *  (*) | *  (*) |
| Ischemic Heart Disease | 22 (5.09) | 30 (8.29) | 46 (6.56) | *  (*) | 31 (4.82) | 41 (6.17) | 58 (5.32) | *  (*) |
| Nicotine Dependency | 104 (24.07) | 86 (23.76) | 164 (23.40) | 28 (28.57) | 113 (17.57) | 121 (18.22) | 193 (17.71) | 45 (24.86) |
| Obesity | 104 (24.07) | 81 (22.38) | 160 (22.82) | 17 (17.35) | 135 (21.00) | 136 (20.48) | 229 (21.01) | 39 (21.55) |
| **Charlson Comorbidity Index** | | | | | | | | |
| 0 | 115 (26.62) | 101 (27.90) | 190 (27.10) | 23 (23.47) | 150 (23.33) | 192 (28.92) | 278 (25.50) | 32 (17.68) |
| 1 | 82 (18.98) | 70 (19.34) | 135 (19.26) | 16 (16.33) | 104 (16.17) | 125 (18.83) | 190 (17.43) | 32 (17.68) |
| 2 | 77 (17.82) | 56 (15.47) | 121 (17.26) | *  (*) | 110 (17.11) | 112 (16.87) | 192 (17.61) | 27 (14.92) |
| 3 | 53 (12.27) | 42 (11.60) | 82 (11.70) | 15 (15.31) | 82 (12.75) | 70 (10.54) | 128 (11.74) | 29 (16.02) |
| 4 | 35 (8.10) | 43 (11.88) | 71 (10.13) | *  (*) | 64 (9.95) | 57 (8.58) | 103 (9.45) | 23 (12.71) |
| 5+ | 70 (16.20) | 50 (13.81) | 102 (14.55) | 21 (21.43) | 133 (20.68) | 108 (16.27) | 199 (18.26) | 38 (20.99) |
| **Prior COVID-19 diagnosis** | | | | | | | | |
| In the 30 days prior to outcome | 0 (0.00) | 0 (0.00) | 0 (0.00) | 0 (0.00) | 0 (0.00) | 0 (0.00) | 0 (0.00) | 0 (0.00) |
| In the 31-365 days prior to outcome | 43 (9.95) | 51 (14.09) | 83 (11.84) | 13 (13.27) | 66 (10.26) | 88 (13.25) | 128 (11.74) | 29 (16.02) |
| **Other vaccine (administered in risk/control intervals)** | | | | | | | | |
| Pneumococcal vaccine | 13 (3.01) | *  (*) | 19 (2.71) | 0 (0.00) | 32 (4.98) | 25 (3.77) | 52 (4.77) | *  (*) |

**Abbreviations:** BNT162b2; WT/OMI BA.4/BA.5, Pfizer COVID-19 BivalentmRNA-1273.222, Moderna COVID-19 Bivalent;

HS, hemorrhagic stroke; NHS, non-hemorrhagic stroke; TIA, transient ischemic attacks; ADI, area deprivations index; ESRD, end-stage renal disease; COPD, chronic obstructive pulmonary disease; COVID-19, coronavirus 2019

* Outcome counts of 10 or fewer and associated statistics are masked to protect the anonymity of the data.

** Reflects beneficiaries’ dual eligibility for Medicare and Medicaid plan benefits.

**eTable 13. Summary of Relative and Attributable Risk of AESI Following COVID-19 Vaccination in the Primary SCCS Analysis, Adjusting for Event Dependent Observation Time**

|  | **BNT162b2; WT/OMI BA.4/BA.5** | | | | | **MRNA-1273.222** | | | | |
| --- | --- | --- | --- | --- | --- | --- | --- | --- | --- | --- |
| **NHS** | | | | | | | | | | |
| **1-21 Day Risk Window** | | | | | | | | | | |
| ***Conditional Poisson Regression*** | | | | | | | | | | |
| *Cases in Risk Window* | 425 | | | | | 215 | | | | |
| *Cases in Control Window* | 953 | | | | | 607 | | | | |
| *Risk Days* | 37,525 | | | | | 22,859 | | | | |
| *Control Days* | 79,659 | | | | | 49,149 | | | | |
| *Model Results* | RR | 95% CI | | P-Value | SE | RR | 95% CI | | P-Value | SE |
|  | 1.06 | 0.94 | 1.19 | 0.33 | 0.06 | 0.88 | 0.75 | 1.03 | 0.12 | 0.08 |
| ***Attributable Risk*** | | | | | | | | | | |
| *# of Eligible Vaccinated* | 2,447,285 | | | | | 1,641,072 | | | | |
| *# of Vaccines* | 5,273,984 | | | | | 3,766,192 | | | | |
| *Person Years* | 102,591,870 | | | | | 68,806,702 | | | | |
| *Attributable Risk per 100,000 Doses* | AR | 95% CI | | P-Value | SE | AR | 95% CI | | P-Value | SE |
|  | 0.99 | -1.01 | 2.99 | 0.33 | 1.02 | -1.79 | -3.93 | 0.36 | 0.10 | 1.10 |
| *Attributable Risk per 100,000 Person Years* | AR | 95% CI | | P-Value | SE | AR | 95% CI | | P-Value | SE |
|  | 8.62 | -8.77 | 26.00 | 0.33 | 8.87 | -15.55 | -34.24 | 3.13 | 0.10 | 9.53 |
| **22-42 Day Risk Window** | | | | | | | | | | |
| ***Conditional Poisson Regression*** | | | | | | | | | | |
| *Cases in Risk Window* | 418 | | | | | 268 | | | | |
| *Cases in Control Window* | 953 | | | | | 607 | | | | |
| *Risk Days* | 36,830 | | | | | 22,595 | | | | |
| *Control Days* | 79,659 | | | | | 49,149 | | | | |
| *Model Results* | RR | 95% CI | | P-Value | SE | RR | 95% CI | | P-Value | SE |
|  | 1.05 | 0.94 | 1.18 | 0.40 | 0.06 | 1.06 | 0.92 | 1.23 | 0.43 | 0.07 |
| ***Attributable Risk*** | | | | | | | | | | |
| *# of Eligible Vaccinated* | 2,447,285 | | | | | 1,641,072 | | | | |
| *# of Vaccines* | 5,273,984 | | | | | 3,766,192 | | | | |
| *Person Years* | 102,591,870 | | | | | 68,806,702 | | | | |
| *Attributable Risk per 100,000 Doses* | AR | 95% CI | | P-Value | SE | AR | 95% CI | | P-Value | SE |
|  | 0.83 | -1.10 | 2.76 | 0.40 | 0.99 | 0.94 | -1.40 | 3.28 | 0.43 | 1.19 |
| *Attributable Risk per 100,000 Person Years* | AR | 95% CI | | P-Value | SE | AR | 95% CI | | P-Value | SE |
|  | 7.23 | -9.62 | 24.07 | 0.40 | 8.59 | 8.18 | -12.18 | 28.55 | 0.43 | 10.39 |
| **TIA** | | | | | | | | | | |
| **1-21 Day Risk Window** | | | | | | | | | | |
| ***Conditional Poisson Regression*** | | | | | | | | | | |
| *Cases in Risk Window* | 389 | | | | | 245 | | | | |
| *Cases in Control Window* | 849 | | | | | 536 | | | | |
| *Risk Days* | 33,703 | | | | | 21,729 | | | | |
| *Control Days* | 76,135 | | | | | 48,958 | | | | |
| *Model Results* | RR | 95% CI | | P-Value | SE | RR | 95% CI | | P-Value | SE |
|  | 1.08 | 0.96 | 1.22 | 0.22 | 0.06 | 1.09 | 0.94 | 1.27 | 0.27 | 0.08 |
| ***Attributable Risk*** | | | | | | | | | | |
| *# of Eligible Vaccinated* | 3,173,426 | | | | | 2,223,852 | | | | |
| *# of Vaccines* | 5,273,984 | | | | | 3,766,192 | | | | |
| *Person Years* | 133,031,634 | | | | | 93,241,357 | | | | |
| *Attributable Risk per 100,000 Doses* | AR | 95% CI | | P-Value | SE | AR | 95% CI | | P-Value | SE |
|  | 0.90 | -0.53 | 2.34 | 0.22 | 0.73 | 0.91 | -0.73 | 2.54 | 0.28 | 0.83 |
| *Attributable Risk per 100,000 Person Years* | AR | 95% CI | | P-Value | SE | AR | 95% CI | | P-Value | SE |
|  | 7.84 | -4.65 | 20.33 | 0.22 | 6.37 | 7.88 | -6.36 | 22.12 | 0.28 | 7.27 |
| **22-42 Day Risk Window** | | | | | | | | | | |
| ***Conditional Poisson Regression*** | | | | | | | | | | |
| *Cases in Risk Window* | 368 | | | | | 254 | | | | |
| *Cases in Control Window* | 849 | | | | | 536 | | | | |
| *Risk Days* | 33,643 | | | | | 21,673 | | | | |
| *Control Days* | 76,135 | | | | | 48,958 | | | | |
| *Model Results* | RR | 95% CI | | P-Value | SE | RR | 95% CI | | P-Value | SE |
|  | 1.02 | 0.91 | 1.16 | 0.71 | 0.06 | 1.12 | 0.97 | 1.31 | 0.13 | 0.08 |
| ***Attributable Risk*** | | | | | | | | | | |
| *# of Eligible Vaccinated* | 3,173,426 | | | | | 2,223,852 | | | | |
| *# of Vaccines* | 5,273,984 | | | | | 3,766,192 | | | | |
| *Person Years* | 133,031,634 | | | | | 93,241,357 | | | | |
| *Attributable Risk per 100,000 Doses* | AR | 95% CI | | P-Value | SE | AR | 95% CI | | P-Value | SE |
|  | 0.27 | -1.14 | 1.67 | 0.71 | 0.72 | 1.26 | -0.38 | 2.90 | 0.13 | 0.84 |
| *Attributable Risk per 100,000 Person Years* | AR | 95% CI | | P-Value | SE | AR | 95% CI | | P-Value | SE |
|  | 2.31 | -9.90 | 14.53 | 0.71 | 6.23 | 10.95 | -3.33 | 25.24 | 0.13 | 7.29 |
| **NHS/TIA** | | | | | | | | | | |
| **1-21 Day Risk Window** | | | | | | | | | | |
| ***Conditional Poisson Regression*** | | | | | | | | | | |
| *Cases in Risk Window* | 712 | | | | | 394 | | | | |
| *Cases in Control Window* | 1,579 | | | | | 977 | | | | |
| *Risk Days* | 62,303 | | | | | 37,995 | | | | |
| *Control Days* | 135,904 | | | | | 83,476 | | | | |
| *Model Results* | RR | 95% CI | | P-Value | SE | RR | 95% CI | | P-Value | SE |
|  | 1.07 | 0.98 | 1.17 | 0.15 | 0.05 | 0.99 | 0.88 | 1.11 | 0.82 | 0.06 |
| ***Attributable Risk*** | | | | | | | | | | |
| *# of Eligible Vaccinated* | 2,447,285 | | | | | 1,641,072 | | | | |
| *# of Vaccines* | 5,273,984 | | | | | 3,766,192 | | | | |
| *Person Years* | 102,591,870 | | | | | 68,806,702 | | | | |
| *Attributable Risk per 100,000 Doses* | AR | 95% CI | | P-Value | SE | AR | 95% CI | | P-Value | SE |
|  | 1.89 | -0.67 | 4.45 | 0.15 | 1.31 | -0.33 | -3.14 | 2.47 | 0.82 | 1.43 |
| *Attributable Risk per 100,000 Person Years* | AR | 95% CI | | P-Value | SE | AR | 95% CI | | P-Value | SE |
|  | 16.47 | -5.84 | 38.77 | 0.15 | 11.38 | -2.89 | -27.31 | 21.52 | 0.82 | 12.46 |
| **22-42 Day Risk Window** | | | | | | | | | | |
| ***Conditional Poisson Regression*** | | | | | | | | | | |
| *Cases in Risk Window* | 686 | | | | | 440 | | | | |
| *Cases in Control Window* | 1,579 | | | | | 977 | | | | |
| *Risk Days* | 61,566 | | | | | 37,700 | | | | |
| *Control Days* | 135,904 | | | | | 83,476 | | | | |
| *Model Results* | RR | 95% CI | | P-Value | SE | RR | 95% CI | | P-Value | SE |
|  | 1.04 | 0.95 | 1.13 | 0.43 | 0.05 | 1.08 | 0.96 | 1.21 | 0.18 | 0.06 |
| ***Attributable Risk*** | | | | | | | | | | |
| *# of Eligible Vaccinated* | 2,447,285 | | | | | 1,641,072 | | | | |
| *# of Vaccines* | 5,273,984 | | | | | 3,766,192 | | | | |
| *Person Years* | 102,591,870 | | | | | 68,806,702 | | | | |
| *Attributable Risk per 100,000 Doses* | AR | 95% CI | | P-Value | SE | AR | 95% CI | | P-Value | SE |
|  | 0.99 | -1.50 | 3.49 | 0.44 | 1.27 | 2.00 | -0.95 | 4.95 | 0.18 | 1.51 |
| *Attributable Risk per 100,000 Person Years* | AR | 95% CI | | P-Value | SE | AR | 95% CI | | P-Value | SE |
|  | 8.65 | -13.08 | 30.39 | 0.44 | 11.09 | 17.40 | -8.30 | 43.10 | 0.18 | 13.11 |
| **HS** | | | | | | | | | | |
| **1-21 Day Risk Window** | | | | | | | | | | |
| ***Conditional Poisson Regression*** | | | | | | | | | | |
| *Cases in Risk Window* | 107 | | | | | 50 | | | | |
| *Cases in Control Window* | 295 | | | | | 172 | | | | |
| *Risk Days* | 10,886 | | | | | 5,891 | | | | |
| *Control Days* | 20,103 | | | | | 11,740 | | | | |
| *Model Results* | RR | 95% CI | | P-Value | SE | RR | 95% CI | | P-Value | SE |
|  | 0.88 | 0.68 | 1.12 | 0.29 | 0.13 | 0.72 | 0.52 | 1.01 | 0.06 | 0.17 |
| ***Attributable Risk*** | | | | | | | | | | |
| *# of Eligible Vaccinated* | 2,117,754 | | | | | 1,399,291 | | | | |
| *# of Vaccines* | 5,273,984 | | | | | 3,766,192 | | | | |
| *Person Years* | 88,778,327 | | | | | 58,670,699 | | | | |
| *Attributable Risk per 100,000 Doses* | AR | 95% CI | | P-Value | SE | AR | 95% CI | | P-Value | SE |
|  | -0.72 | -2.07 | 0.63 | 0.30 | 0.69 | -1.37 | -2.72 | -0.02 | 0.05 | 0.69 |
| *Attributable Risk per 100,000 Person Years* | AR | 95% CI | | P-Value | SE | AR | 95% CI | | P-Value | SE |
|  | -6.28 | -18.05 | 5.50 | 0.30 | 6.01 | -11.92 | -23.67 | -0.16 | 0.05 | 6.00 |
| **22-42 Day Risk Window** | | | | | | | | | | |
| ***Conditional Poisson Regression*** | | | | | | | | | | |
| *Cases in Risk Window* | 123 | | | | | 61 | | | | |
| *Cases in Control Window* | 295 | | | | | 172 | | | | |
| *Risk Days* | 10,277 | | | | | 5,731 | | | | |
| *Control Days* | 20,103 | | | | | 11,740 | | | | |
| *Model Results* | RR | 95% CI | | P-Value | SE | RR | 95% CI | | P-Value | SE |
|  | 0.98 | 0.78 | 1.23 | 0.84 | 0.12 | 0.83 | 0.61 | 1.13 | 0.23 | 0.16 |
| ***Attributable Risk*** | | | | | | | | | | |
| *# of Eligible Vaccinated* | 2,117,754 | | | | | 1,399,291 | | | | |
| *# of Vaccines* | 5,273,984 | | | | | 3,766,192 | | | | |
| *Person Years* | 88,778,327 | | | | | 58,670,699 | | | | |
| *Attributable Risk per 100,000 Doses* | AR | 95% CI | | P-Value | SE | AR | 95% CI | | P-Value | SE |
|  | -0.14 | -1.47 | 1.19 | 0.84 | 0.68 | -0.91 | -2.36 | 0.55 | 0.22 | 0.74 |
| *Attributable Risk per 100,000 Person Years* | AR | 95% CI | | P-Value | SE | AR | 95% CI | | P-Value | SE |
|  | -1.20 | -12.79 | 10.40 | 0.84 | 5.92 | -7.92 | -20.59 | 4.75 | 0.22 | 6.46 |
| **Abbreviations:** BNT162b2; WT/OMI BA.4/BA.5, Pfizer COVID-19 BivalentmRNA-1273.222, Moderna COVID-19 Bivalent;  HS, hemorrhagic stroke; NHS, non-hemorrhagic stroke; TIA, transient ischemic attacks | | | | | | | | | | |

**eTable 14. Summary of Relative and Attributable Risk of AESI Following COVID-19 Vaccination in the Primary SCCS Analysis, Adjusting for Seasonality and Event Dependent Observation Time**

|  | **BNT162b2; WT/OMI BA.4/BA.5** | | | | | **MRNA-1273.222** | | | | |
| --- | --- | --- | --- | --- | --- | --- | --- | --- | --- | --- |
| **NHS** | | | | | | | | | | |
| **1-21 Day Risk Window** | | | | | | | | | | |
| ***Conditional Poisson Regression*** | | | | | | | | | | |
| *Cases in Risk Window* | 425 | | | | | 215 | | | | |
| *Cases in Control Window* | 953 | | | | | 607 | | | | |
| *Risk Days* | 37,525 | | | | | 22,859 | | | | |
| *Control Days* | 79,659 | | | | | 49,149 | | | | |
| *Model Results* | RR | 95% CI | | P-Value | SE | RR | 95% CI | | P-Value | SE |
|  | 1.09 | 0.97 | 1.23 | 0.16 | 0.06 | 0.90 | 0.77 | 1.06 | 0.21 | 0.08 |
| ***Attributable Risk*** | | | | | | | | | | |
| *# of Eligible Vaccinated* | 2,447,285 | | | | | 1,641,072 | | | | |
| *# of Vaccines* | 5,273,984 | | | | | 3,766,192 | | | | |
| *Person Years* | 102,591,870 | | | | | 68,806,702 | | | | |
| *Attributable Risk per 100,000 Doses* | AR | 95% CI | | P-Value | SE | AR | 95% CI | | P-Value | SE |
|  | 1.42 | -0.56 | 3.40 | 0.16 | 1.01 | -1.40 | -3.55 | 0.75 | 0.20 | 1.10 |
| *Attributable Risk per 100,000 Person Years* | AR | 95% CI | | P-Value | SE | AR | 95% CI | | P-Value | SE |
|  | 12.36 | -4.88 | 29.60 | 0.16 | 8.79 | -12.21 | -30.93 | 6.50 | 0.20 | 9.55 |
| **22-42 Day Risk Window** | | | | | | | | | | |
| ***Conditional Poisson Regression*** | | | | | | | | | | |
| *Cases in Risk Window* | 418 | | | | | 268 | | | | |
| *Cases in Control Window* | 953 | | | | | 607 | | | | |
| *Risk Days* | 36,830 | | | | | 22,595 | | | | |
| *Control Days* | 79,659 | | | | | 49,149 | | | | |
| *Model Results* | RR | 95% CI | | P-Value | SE | RR | 95% CI | | P-Value | SE |
|  | 1.06 | 0.94 | 1.19 | 0.32 | 0.06 | 1.07 | 0.92 | 1.24 | 0.37 | 0.07 |
| ***Attributable Risk*** | | | | | | | | | | |
| *# of Eligible Vaccinated* | 2,447,285 | | | | | 1,641,072 | | | | |
| *# of Vaccines* | 5,273,984 | | | | | 3,766,192 | | | | |
| *Person Years* | 102,591,870 | | | | | 68,806,702 | | | | |
| *Attributable Risk per 100,000 Doses* | AR | 95% CI | | P-Value | SE | AR | 95% CI | | P-Value | SE |
|  | 0.98 | -0.97 | 2.93 | 0.33 | 0.99 | 1.06 | -1.28 | 3.41 | 0.37 | 1.19 |
| *Attributable Risk per 100,000 Person Years* | AR | 95% CI | | P-Value | SE | AR | 95% CI | | P-Value | SE |
|  | 8.51 | -8.45 | 25.47 | 0.33 | 8.65 | 9.26 | -11.13 | 29.64 | 0.37 | 10.40 |
| **TIA** | | | | | | | | | | |
| **1-21 Day Risk Window** | | | | | | | | | | |
| ***Conditional Poisson Regression*** | | | | | | | | | | |
| *Cases in Risk Window* | 389 | | | | | 245 | | | | |
| *Cases in Control Window* | 849 | | | | | 536 | | | | |
| *Risk Days* | 33,703 | | | | | 21,729 | | | | |
| *Control Days* | 76,135 | | | | | 48,958 | | | | |
| *Model Results* | RR | 95% CI | | P-Value | SE | RR | 95% CI | | P-Value | SE |
|  | 1.08 | 0.96 | 1.22 | 0.21 | 0.06 | 1.09 | 0.93 | 1.27 | 0.28 | 0.08 |
| ***Attributable Risk*** | | | | | | | | | | |
| *# of Eligible Vaccinated* | 3,173,426 | | | | | 2,223,852 | | | | |
| *# of Vaccines* | 5,273,984 | | | | | 3,766,192 | | | | |
| *Person Years* | 133,031,634 | | | | | 93,241,357 | | | | |
| *Attributable Risk per 100,000 Doses* | AR | 95% CI | | P-Value | SE | AR | 95% CI | | P-Value | SE |
|  | 0.91 | -0.53 | 2.35 | 0.22 | 0.73 | 0.89 | -0.74 | 2.51 | 0.28 | 0.83 |
| *Attributable Risk per 100,000 Person Years* | AR | 95% CI | | P-Value | SE | AR | 95% CI | | P-Value | SE |
|  | 7.92 | -4.60 | 20.44 | 0.22 | 6.39 | 7.73 | -6.42 | 21.89 | 0.28 | 7.22 |
| **22-42 Day Risk Window** | | | | | | | | | | |
| ***Conditional Poisson Regression*** | | | | | | | | | | |
| *Cases in Risk Window* | 368 | | | | | 254 | | | | |
| *Cases in Control Window* | 849 | | | | | 536 | | | | |
| *Risk Days* | 33,643 | | | | | 21,673 | | | | |
| *Control Days* | 76,135 | | | | | 48,958 | | | | |
| *Model Results* | RR | 95% CI | | P-Value | SE | RR | 95% CI | | P-Value | SE |
|  | 1.01 | 0.89 | 1.14 | 0.92 | 0.06 | 1.10 | 0.95 | 1.28 | 0.20 | 0.08 |
| ***Attributable Risk*** | | | | | | | | | | |
| *# of Eligible Vaccinated* | 3,173,426 | | | | | 2,223,852 | | | | |
| *# of Vaccines* | 5,273,984 | | | | | 3,766,192 | | | | |
| *Person Years* | 133,031,634 | | | | | 93,241,357 | | | | |
| *Attributable Risk per 100,000 Doses* | AR | 95% CI | | P-Value | SE | AR | 95% CI | | P-Value | SE |
|  | 0.07 | -1.34 | 1.47 | 0.92 | 0.72 | 1.07 | -0.58 | 2.73 | 0.20 | 0.85 |
| *Attributable Risk per 100,000 Person Years* | AR | 95% CI | | P-Value | SE | AR | 95% CI | | P-Value | SE |
|  | 0.59 | -11.64 | 12.82 | 0.92 | 6.24 | 9.34 | -5.08 | 23.77 | 0.20 | 7.36 |
| **NHS/TIA** | | | | | | | | | | |
| **1-21 Day Risk Window** | | | | | | | | | | |
| ***Conditional Poisson Regression*** | | | | | | | | | | |
| *Cases in Risk Window* | 712 | | | | | 394 | | | | |
| *Cases in Control Window* | 1,579 | | | | | 977 | | | | |
| *Risk Days* | 62,303 | | | | | 37,995 | | | | |
| *Control Days* | 135,904 | | | | | 83,476 | | | | |
| *Model Results* | RR | 95% CI | | P-Value | SE | RR | 95% CI | | P-Value | SE |
|  | 1.10 | 1.00 | 1.20 | 0.05 | 0.05 | 1.01 | 0.90 | 1.14 | 0.87 | 0.06 |
| ***Attributable Risk*** | | | | | | | | | | |
| *# of Eligible Vaccinated* | 2,447,285 | | | | | 1,641,072 | | | | |
| *# of Vaccines* | 5,273,984 | | | | | 3,766,192 | | | | |
| *Person Years* | 102,591,870 | | | | | 68,806,702 | | | | |
| *Attributable Risk per 100,000 Doses* | AR | 95% CI | | P-Value | SE | AR | 95% CI | | P-Value | SE |
|  | 2.53 | -0.01 | 5.08 | 0.05 | 1.30 | 0.23 | -2.58 | 3.04 | 0.87 | 1.43 |
| *Attributable Risk per 100,000 Person Years* | AR | 95% CI | | P-Value | SE | AR | 95% CI | | P-Value | SE |
|  | 22.04 | -0.11 | 44.20 | 0.05 | 11.30 | 2.00 | -22.43 | 26.43 | 0.87 | 12.47 |
| **22-42 Day Risk Window** | | | | | | | | | | |
| ***Conditional Poisson Regression*** | | | | | | | | | | |
| *Cases in Risk Window* | 686 | | | | | 440 | | | | |
| *Cases in Control Window* | 1,579 | | | | | 977 | | | | |
| *Risk Days* | 61,566 | | | | | 37,700 | | | | |
| *Control Days* | 135,904 | | | | | 83,476 | | | | |
| *Model Results* | RR | 95% CI | | P-Value | SE | RR | 95% CI | | P-Value | SE |
|  | 1.04 | 0.95 | 1.14 | 0.39 | 0.05 | 1.08 | 0.97 | 1.21 | 0.17 | 0.06 |
| ***Attributable Risk*** | | | | | | | | | | |
| *# of Eligible Vaccinated* | 2,447,285 | | | | | 1,641,072 | | | | |
| *# of Vaccines* | 5,273,984 | | | | | 3,766,192 | | | | |
| *Person Years* | 102,591,870 | | | | | 68,806,702 | | | | |
| *Attributable Risk per 100,000 Doses* | AR | 95% CI | | P-Value | SE | AR | 95% CI | | P-Value | SE |
|  | 1.09 | -1.41 | 3.59 | 0.39 | 1.28 | 2.07 | -0.90 | 5.04 | 0.17 | 1.52 |
| *Attributable Risk per 100,000 Person Years* | AR | 95% CI | | P-Value | SE | AR | 95% CI | | P-Value | SE |
|  | 9.49 | -12.31 | 31.29 | 0.39 | 11.12 | 17.99 | -7.87 | 43.84 | 0.17 | 13.19 |
| **HS** | | | | | | | | | | |
| **1-21 Day Risk Window** | | | | | | | | | | |
| ***Conditional Poisson Regression*** | | | | | | | | | | |
| *Cases in Risk Window* | 107 | | | | | 50 | | | | |
| *Cases in Control Window* | 295 | | | | | 172 | | | | |
| *Risk Days* | 10,886 | | | | | 5,891 | | | | |
| *Control Days* | 20,103 | | | | | 11,740 | | | | |
| *Model Results* | RR | 95% CI | | P-Value | SE | RR | 95% CI | | P-Value | SE |
|  | 0.90 | 0.70 | 1.15 | 0.40 | 0.13 | 0.74 | 0.53 | 1.04 | 0.08 | 0.17 |
| ***Attributable Risk*** | | | | | | | | | | |
| *# of Eligible Vaccinated* | 2,117,754 | | | | | 1,399,291 | | | | |
| *# of Vaccines* | 5,273,984 | | | | | 3,766,192 | | | | |
| *Person Years* | 88,778,327 | | | | | 58,670,699 | | | | |
| *Attributable Risk per 100,000 Doses* | AR | 95% CI | | P-Value | SE | AR | 95% CI | | P-Value | SE |
|  | -0.57 | -1.91 | 0.76 | 0.40 | 0.68 | -1.24 | -2.58 | 0.10 | 0.07 | 0.68 |
| *Attributable Risk per 100,000 Person Years* | AR | 95% CI | | P-Value | SE | AR | 95% CI | | P-Value | SE |
|  | -5.00 | -16.60 | 6.60 | 0.40 | 5.92 | -10.79 | -22.44 | 0.85 | 0.07 | 5.94 |
| **22-42 Day Risk Window** | | | | | | | | | | |
| ***Conditional Poisson Regression*** | | | | | | | | | | |
| *Cases in Risk Window* | 123 | | | | | 61 | | | | |
| *Cases in Control Window* | 295 | | | | | 172 | | | | |
| *Risk Days* | 10,277 | | | | | 5,731 | | | | |
| *Control Days* | 20,103 | | | | | 11,740 | | | | |
| *Model Results* | RR | 95% CI | | P-Value | SE | RR | 95% CI | | P-Value | SE |
|  | 0.99 | 0.79 | 1.25 | 0.95 | 0.12 | 0.84 | 0.62 | 1.15 | 0.27 | 0.16 |
| ***Attributable Risk*** | | | | | | | | | | |
| *# of Eligible Vaccinated* | 2,117,754 | | | | | 1,399,291 | | | | |
| *# of Vaccines* | 5,273,984 | | | | | 3,766,192 | | | | |
| *Person Years* | 88,778,327 | | | | | 58,670,699 | | | | |
| *Attributable Risk per 100,000 Doses* | AR | 95% CI | | P-Value | SE | AR | 95% CI | | P-Value | SE |
|  | -0.04 | -1.37 | 1.29 | 0.95 | 0.68 | -0.83 | -2.27 | 0.61 | 0.26 | 0.74 |
| *Attributable Risk per 100,000 Person Years* | AR | 95% CI | | P-Value | SE | AR | 95% CI | | P-Value | SE |
|  | -0.39 | -11.97 | 11.20 | 0.95 | 5.91 | -7.19 | -19.73 | 5.35 | 0.26 | 6.40 |
| **Abbreviations:** BNT162b2; WT/OMI BA.4/BA.5, Pfizer COVID-19 BivalentmRNA-1273.222, Moderna COVID-19 Bivalent;  HS, hemorrhagic stroke; NHS, non-hemorrhagic stroke; TIA, transient ischemic attacks | | | | | | | | | | |

**eTable 15. Summary of Relative and Attributable Risk of NHS Following COVID-19 Vaccination in the Primary SCCS Analysis, Adjusting for PPV and Event Dependent Observation Time**

|  | **BNT162b2; WT/OMI BA.4/BA.5** | | | | | **MRNA-1273.222** | | | | |
| --- | --- | --- | --- | --- | --- | --- | --- | --- | --- | --- |
| **NHS** | | | | | | | | |  |  |
| ***1-21 Day Risk Window*** |  |  |  |  |  |  |  |  |  |  |
| ***Conditional Poisson Regression*** |  |  |  |  |  |  |  |  |  |  |
| *Cases in Risk Window* | 342 | | | | | 173 | | | | |
| *Cases in Control Window* | 767 | | | | | 489 | | | | |
| *Risk Days* | 30,200 | | | | | 18,391 | | | | |
| *Control Days* | 64,101 | | | | | 39,543 | | | | |
| *Model Results* | IRR | 95% CI | | P-Value | SE | IRR | 95% CI | | P-Value | SE |
|  | 1.06 | 0.92 | 1.23 | 0.42 | 0.07 | 0.88 | 0.72 | 1.07 | 0.19 | 0.10 |
| ***Attributable Risk*** |  |  |  |  |  |  |  |  |  |  |
| *# of Eligible Vaccinated* | 2,447,285 | | | | | 1,641,072 | | | | |
| *# of Vaccines* | 5,273,984 | | | | | 3,766,192 | | | | |
| *Person Days* | 102,591,870 | | | | | 68,806,702 | | | | |
| *Attributable Risk per 100,000 Doses* | AR | 95% CI | | P-Value | SE | AR | 95% CI | | P-Value | SE |
|  | 0.81 | -1.25 | 2.59 | NA | NA | -1.46 | -4.05 | 0.66 | NA | NA |
| *Attributable Risk per 100,000 Person Years* | AR | 95% CI | | P-Value | SE | AR | 95% CI | | P-Value | SE |
|  | 7.04 | -10.84 | 22.51 | NA | NA | -12.75 | -35.27 | 5.77 | NA | NA |
| ***22-42 Day Risk Window*** |  |  |  |  |  |  |  |  |  |  |
| ***Conditional Poisson Regression*** |  |  |  |  |  |  |  |  |  |  |
| *Cases in Risk Window* | 336 | | | | | 216 | | | | |
| *Cases in Control Window* | 767 | | | | | 489 | | | | |
| *Risk Days* | 29,638 | | | | | 18,178 | | | | |
| *Control Days* | 64,101 | | | | | 39,543 | | | | |
| *Model Results* | IRR | 95% CI | | P-Value | SE | IRR | 95% CI | | P-Value | SE |
|  | 1.05 | 0.91 | 1.21 | 0.49 | 0.07 | 1.06 | 0.89 | 1.27 | 0.52 | 0.09 |
| ***Attributable Risk*** |  |  |  |  |  |  |  |  |  |  |
| *# of Eligible Vaccinated* | 2,447,285 | | | | | 1,641,072 | | | | |
| *# of Vaccines* | 5,273,984 | | | | | 3,766,192 | | | | |
| *Person Days* | 102,591,870 | | | | | 68,806,702 | | | | |
| *Attributable Risk per 100,000 Doses* | AR | 95% CI | | P-Value | SE | AR | 95% CI | | P-Value | SE |
|  | 0.67 | -1.32 | 2.41 | NA | NA | 0.75 | -1.67 | 2.77 | NA | NA |
| *Attributable Risk per 100,000 Person Years* | AR | 95% CI | | P-Value | SE | AR | 95% CI | | P-Value | SE |
|  | 5.86 | -11.53 | 20.95 | NA | NA | 6.50 | -14.50 | 24.07 | NA | NA |
| **Abbreviations:** BNT162b2; WT/OMI BA.4/BA.5, Pfizer COVID-19 BivalentmRNA-1273.222, Moderna COVID-19 Bivalent;  HS, hemorrhagic stroke; NHS, non-hemorrhagic stroke; TIA, transient ischemic attacks | | | | | | | | | | |

**eTable 16. Summary of Relative and Attributable Risk of AESI Following BNT162b2; WT/OMI BA.4/BA.5 COVID-19 Vaccination in the Primary SCCS Analysis Stratified by Age, Adjusting for Event Dependent Observation Time**

|  | **BNT162b2; WT/OMI BA.4/BA.5** | | | | | | | | | | | | | | |
| --- | --- | --- | --- | --- | --- | --- | --- | --- | --- | --- | --- | --- | --- | --- | --- |
|  | **65-74** | | | | | **75-84** | | | | | **85+** | | | | |
| **NHS** | | | | | | | | | | | | | | | |
| **1-21 Day Risk Window** | | | | | | | | | | | | | | | |
| ***Conditional Poisson Regression*** | | | | | | | | | | | | | | | |
| *Cases in Risk Window* | 144 | | | | | 145 | | | | | 136 | | | | |
| *Cases in Control Window* | 305 | | | | | 405 | | | | | 243 | | | | |
| *Risk Days* | 12,270 | | | | | 15,285 | | | | | 9,970 | | | | |
| *Control Days* | 26,735 | | | | | 32,747 | | | | | 20,177 | | | | |
| *Model Results* | RR | 95% CI | | P-Value | SE | RR | 95% CI | | P-Value | SE | RR | 95% CI | | P-Value | SE |
|  | 1.14 | 0.93 | 1.40 | 0.20 | 0.10 | 0.84 | 0.69 | 1.03 | 0.09 | 0.10 | 1.36 | 1.09 | 1.69 | 0.01 | 0.11 |
| ***Attributable Risk*** | | | | | | | | | | | | | | | |
| *# of Eligible Vaccinated* | 1,231,349 | | | | | 916,804 | | | | | 299,132 | | | | |
| *Person Years* | 51,637,519 | | | | | 38,437,348 | | | | | 12,517,003 | | | | |
| *Attributable Risk per 100,000 Doses* | AR | 95% CI | | P-Value | SE | AR | 95% CI | | P-Value | SE | AR | 95% CI | | P-Value | SE |
|  | 1.46 | -0.78 | 3.69 | 0.20 | 1.14 | -2.94 | -6.20 | 0.33 | 0.08 | 1.67 | 11.93 | 3.02 | 20.84 | 0.01 | 4.54 |
| *Attributable Risk per 100,000 Person Years* | AR | 95% CI | | P-Value | SE | AR | 95% CI | | P-Value | SE | AR | 95% CI | | P-Value | SE |
|  | 12.68 | -6.76 | 32.12 | 0.20 | 9.92 | -25.56 | -53.98 | 2.86 | 0.08 | 14.50 | 104.05 | 26.36 | 181.74 | 0.01 | 39.64 |
| **22-42 Day Risk Window** | | | | | | | | | | | | | | | |
| ***Conditional Poisson Regression*** | | | | | | | | | | | | | | | |
| *Cases in Risk Window* | 137 | | | | | 182 | | | | | 99 | | | | |
| *Cases in Control Window* | 305 | | | | | 405 | | | | | 243 | | | | |
| *Risk Days* | 12,143 | | | | | 15,078 | | | | | 9,609 | | | | |
| *Control Days* | 26,735 | | | | | 32,747 | | | | | 20,177 | | | | |
| *Model Results* | RR | 95% CI | | P-Value | SE | RR | 95% CI | | P-Value | SE | RR | 95% CI | | P-Value | SE |
|  | 1.08 | 0.88 | 1.32 | 0.46 | 0.10 | 1.06 | 0.89 | 1.27 | 0.52 | 0.09 | 1.00 | 0.79 | 1.27 | 0.98 | 0.12 |
| ***Attributable Risk*** | | | | | | | | | | | | | | | |
| *# of Eligible Vaccinated* | 1,231,349 | | | | | 916,804 | | | | | 299,132 | | | | |
| *Person Years* | 51,637,519 | | | | | 38,437,348 | | | | | 12,517,003 | | | | |
| *Attributable Risk per 100,000 Doses* | AR | 95% CI | | P-Value | SE | AR | 95% CI | | P-Value | SE | AR | 95% CI | | P-Value | SE |
|  | 0.82 | -1.38 | 3.01 | 0.47 | 1.12 | 1.12 | -2.36 | 4.60 | 0.53 | 1.78 | -0.09 | -7.82 | 7.64 | 0.98 | 3.94 |
| *Attributable Risk per 100,000 Person Years* | AR | 95% CI | | P-Value | SE | AR | 95% CI | | P-Value | SE | AR | 95% CI | | P-Value | SE |
|  | 7.10 | -12.00 | 26.19 | 0.47 | 9.74 | 9.74 | -20.58 | 40.05 | 0.53 | 15.47 | -0.83 | -68.25 | 66.60 | 0.98 | 34.40 |
| **TIA** | | | | | | | | | | | | | | | |
| **1-21 Day Risk Window** | | | | | | | | | | | | | | | |
| ***Conditional Poisson Regression*** | | | | | | | | | | | | | | | |
| *Cases in Risk Window* | 116 | | | | | 167 | | | | | 106 | | | | |
| *Cases in Control Window* | 262 | | | | | 375 | | | | | 212 | | | | |
| *Risk Days* | 10,425 | | | | | 14,818 | | | | | 8,460 | | | | |
| *Control Days* | 23,656 | | | | | 33,439 | | | | | 19,040 | | | | |
| *Model Results* | RR | 95% CI | | P-Value | SE | RR | 95% CI | | P-Value | SE | RR | 95% CI | | P-Value | SE |
|  | 1.05 | 0.84 | 1.31 | 0.66 | 0.11 | 1.06 | 0.88 | 1.27 | 0.53 | 0.09 | 1.20 | 0.95 | 1.51 | 0.13 | 0.12 |
| ***Attributable Risk*** | | | | | | | | | | | | | | | |
| *# of Eligible Vaccinated* | 1,593,152 | | | | | 1,182,051 | | | | | 398,223 | | | | |
| *Person Years* | 66,810,594 | | | | | 49,558,044 | | | | | 16,662,996 | | | | |
| *Attributable Risk per 100,000 Doses* | AR | 95% CI | | P-Value | SE | AR | 95% CI | | P-Value | SE | AR | 95% CI | | P-Value | SE |
|  | 0.35 | -1.23 | 1.93 | 0.66 | 0.81 | 0.80 | -1.71 | 3.32 | 0.53 | 1.28 | 4.37 | -1.45 | 10.18 | 0.14 | 2.97 |
| *Attributable Risk per 100,000 Person Years* | AR | 95% CI | | P-Value | SE | AR | 95% CI | | P-Value | SE | AR | 95% CI | | P-Value | SE |
|  | 3.06 | -10.68 | 16.80 | 0.66 | 7.01 | 6.99 | -14.92 | 28.90 | 0.53 | 11.18 | 38.08 | -12.68 | 88.83 | 0.14 | 25.90 |
| **22-42 Day Risk Window** | | | | | | | | | | | | | | | |
| ***Conditional Poisson Regression*** | | | | | | | | | | | | | | | |
| *Cases in Risk Window* | 119 | | | | | 164 | | | | | 85 | | | | |
| *Cases in Control Window* | 262 | | | | | 375 | | | | | 212 | | | | |
| *Risk Days* | 10,416 | | | | | 14,788 | | | | | 8,439 | | | | |
| *Control Days* | 23,656 | | | | | 33,439 | | | | | 19,040 | | | | |
| *Model Results* | RR | 95% CI | | P-Value | SE | RR | 95% CI | | P-Value | SE | RR | 95% CI | | P-Value | SE |
|  | 1.08 | 0.87 | 1.34 | 0.51 | 0.11 | 1.04 | 0.86 | 1.25 | 0.69 | 0.09 | 0.95 | 0.74 | 1.22 | 0.70 | 0.13 |
| ***Attributable Risk*** | | | | | | | | | | | | | | | |
| *# of Eligible Vaccinated* | 1,593,152 | | | | | 1,182,051 | | | | | 398,223 | | | | |
| *Person Years* | 66,810,594 | | | | | 49,558,044 | | | | | 16,662,996 | | | | |
| *Attributable Risk per 100,000 Doses* | AR | 95% CI | | P-Value | SE | AR | 95% CI | | P-Value | SE | AR | 95% CI | | P-Value | SE |
|  | 0.53 | -1.06 | 2.12 | 0.52 | 0.81 | 0.50 | -2.00 | 3.01 | 0.69 | 1.28 | -1.09 | -6.52 | 4.34 | 0.69 | 2.77 |
| *Attributable Risk per 100,000 Person Years* | AR | 95% CI | | P-Value | SE | AR | 95% CI | | P-Value | SE | AR | 95% CI | | P-Value | SE |
|  | 4.59 | -9.24 | 18.43 | 0.52 | 7.06 | 4.38 | -17.40 | 26.16 | 0.69 | 11.11 | -9.51 | -56.86 | 37.84 | 0.69 | 24.16 |
| **NHS/TIA** | | | | | | | | | | | | | | | |
| **1-21 Day Risk Window** | | | | | | | | | | | | | | | |
| ***Conditional Poisson Regression*** | | | | | | | | | | | | | | | |
| *Cases in Risk Window* | 235 | | | | | 267 | | | | | 210 | | | | |
| *Cases in Control Window* | 505 | | | | | 682 | | | | | 392 | | | | |
| *Risk Days* | 20,301 | | | | | 26,176 | | | | | 15,826 | | | | |
| *Control Days* | 44,972 | | | | | 57,434 | | | | | 33,498 | | | | |
| *Model Results* | RR | 95% CI | | P-Value | SE | RR | 95% CI | | P-Value | SE | RR | 95% CI | | P-Value | SE |
|  | 1.11 | 0.95 | 1.30 | 0.17 | 0.08 | 0.93 | 0.80 | 1.07 | 0.31 | 0.07 | 1.28 | 1.08 | 1.52 | 0.01 | 0.09 |
| ***Attributable Risk*** | | | | | | | | | | | | | | | |
| *# of Eligible Vaccinated* | 1,231,349 | | | | | 916,804 | | | | | 299,132 | | | | |
| *Person Years* | 51,637,519 | | | | | 38,437,348 | | | | | 12,517,003 | | | | |
| *Attributable Risk per 100,000 Doses* | AR | 95% CI | | P-Value | SE | AR | 95% CI | | P-Value | SE | AR | 95% CI | | P-Value | SE |
|  | 1.96 | -0.95 | 4.87 | 0.19 | 1.49 | -2.28 | -6.59 | 2.04 | 0.30 | 2.20 | 15.35 | 4.34 | 26.36 | 0.01 | 5.62 |
| *Attributable Risk per 100,000 Person Years* | AR | 95% CI | | P-Value | SE | AR | 95% CI | | P-Value | SE | AR | 95% CI | | P-Value | SE |
|  | 17.07 | -8.28 | 42.41 | 0.19 | 12.93 | -19.81 | -57.41 | 17.79 | 0.30 | 19.18 | 133.93 | 37.89 | 229.97 | 0.01 | 49.00 |
| **22-42 Day Risk Window** | | | | | | | | | | | | | | | |
| ***Conditional Poisson Regression*** | | | | | | | | | | | | | | | |
| *Cases in Risk Window* | 229 | | | | | 302 | | | | | 155 | | | | |
| *Cases in Control Window* | 505 | | | | | 682 | | | | | 392 | | | | |
| *Risk Days* | 20,165 | | | | | 25,939 | | | | | 15,462 | | | | |
| *Control Days* | 44,972 | | | | | 57,434 | | | | | 33,498 | | | | |
| *Model Results* | RR | 95% CI | | P-Value | SE | RR | 95% CI | | P-Value | SE | RR | 95% CI | | P-Value | SE |
|  | 1.08 | 0.93 | 1.27 | 0.31 | 0.08 | 1.05 | 0.91 | 1.20 | 0.50 | 0.07 | 0.96 | 0.79 | 1.16 | 0.66 | 0.10 |
| ***Attributable Risk*** | | | | | | | | | | | | | | | |
| *# of Eligible Vaccinated* | 1,231,349 | | | | | 916,804 | | | | | 299,132 | | | | |
| *Person Years* | 51,637,519 | | | | | 38,437,348 | | | | | 12,517,003 | | | | |
| *Attributable Risk per 100,000 Doses* | AR | 95% CI | | P-Value | SE | AR | 95% CI | | P-Value | SE | AR | 95% CI | | P-Value | SE |
|  | 1.44 | -1.40 | 4.28 | 0.32 | 1.45 | 1.51 | -2.93 | 5.94 | 0.51 | 2.26 | -2.25 | -12.07 | 7.57 | 0.65 | 5.01 |
| *Attributable Risk per 100,000 Person Years* | AR | 95% CI | | P-Value | SE | AR | 95% CI | | P-Value | SE | AR | 95% CI | | P-Value | SE |
|  | 12.53 | -12.22 | 37.28 | 0.32 | 12.63 | 13.12 | -25.49 | 51.72 | 0.51 | 19.69 | -19.62 | -105.31 | 66.07 | 0.65 | 43.72 |
| **HS** | | | | | | | | | | | | | | | |
| **1-21 Day Risk Window** | | | | | | | | | | | | | | | |
| ***Conditional Poisson Regression*** | | | | | | | | | | | | | | | |
| *Cases in Risk Window* | 36 | | | | | 47 | | | | | 24 | | | | |
| *Cases in Control Window* | 98 | | | | | 124 | | | | | 73 | | | | |
| *Risk Days* | 3,812 | | | | | 4,506 | | | | | 2,568 | | | | |
| *Control Days* | 7,458 | | | | | 8,615 | | | | | 4,030 | | | | |
| *Model Results* | RR | 95% CI | | P-Value | SE | RR | 95% CI | | P-Value | SE | RR | 95% CI | | P-Value | SE |
|  | 0.99 | 0.66 | 1.47 | 0.95 | 0.20 | 0.82 | 0.56 | 1.21 | 0.31 | 0.20 | 0.77 | 0.43 | 1.38 | 0.38 | 0.30 |
| ***Attributable Risk*** | | | | | | | | | | | | | | | |
| *# of Eligible Vaccinated* | 1,068,732 | | | | | 796,024 | | | | | 252,998 | | | | |
| *Person Years* | 44,817,385 | | | | | 33,374,011 | | | | | 10,586,931 | | | | |
| *Attributable Risk per 100,000 Doses* | AR | 95% CI | | P-Value | SE | AR | 95% CI | | P-Value | SE | AR | 95% CI | | P-Value | SE |
|  | -0.04 | -1.41 | 1.33 | 0.95 | 0.70 | -1.29 | -3.80 | 1.22 | 0.31 | 1.28 | -2.84 | -10.39 | 4.72 | 0.46 | 3.86 |
| *Attributable Risk per 100,000 Person Years* | AR | 95% CI | | P-Value | SE | AR | 95% CI | | P-Value | SE | AR | 95% CI | | P-Value | SE |
|  | -0.36 | -12.30 | 11.58 | 0.95 | 6.09 | -11.25 | -33.11 | 10.62 | 0.31 | 11.16 | -24.74 | -90.65 | 41.17 | 0.46 | 33.63 |
| **22-42 Day Risk Window** | | | | | | | | | | | | | | | |
| ***Conditional Poisson Regression*** | | | | | | | | | | | | | | | |
| *Cases in Risk Window* | 48 | | | | | 47 | | | | | 28 | | | | |
| *Cases in Control Window* | 98 | | | | | 124 | | | | | 73 | | | | |
| *Risk Days* | 3,689 | | | | | 4,263 | | | | | 2,325 | | | | |
| *Control Days* | 7,458 | | | | | 8,615 | | | | | 4,030 | | | | |
| *Model Results* | RR | 95% CI | | P-Value | SE | RR | 95% CI | | P-Value | SE | RR | 95% CI | | P-Value | SE |
|  | 1.15 | 0.79 | 1.67 | 0.46 | 0.19 | 0.89 | 0.62 | 1.27 | 0.52 | 0.18 | 0.85 | 0.51 | 1.43 | 0.55 | 0.26 |
| ***Attributable Risk*** | | | | | | | | | | | | | | | |
| *# of Eligible Vaccinated* | 1,068,732 | | | | | 796,024 | | | | | 252,998 | | | | |
| *Person Years* | 44,817,385 | | | | | 33,374,011 | | | | | 10,586,931 | | | | |
| *Attributable Risk per 100,000 Doses* | AR | 95% CI | | P-Value | SE | AR | 95% CI | | P-Value | SE | AR | 95% CI | | P-Value | SE |
|  | 0.59 | -0.97 | 2.15 | 0.46 | 0.80 | -0.74 | -2.89 | 1.42 | 0.50 | 1.10 | -1.91 | -8.34 | 4.53 | 0.56 | 3.28 |
| *Attributable Risk per 100,000 Person Years* | AR | 95% CI | | P-Value | SE | AR | 95% CI | | P-Value | SE | AR | 95% CI | | P-Value | SE |
|  | 5.14 | -8.47 | 18.75 | 0.46 | 6.95 | -6.41 | -25.19 | 12.37 | 0.50 | 9.58 | -16.64 | -72.78 | 39.50 | 0.56 | 28.64 |

**Abbreviations:** BNT162b2; WT/OMI BA.4/BA.5, Pfizer COVID-19 BivalentmRNA-1273.222, Moderna COVID-19 Bivalent;

HS, hemorrhagic stroke; NHS, non-hemorrhagic stroke; TIA, transient ischemic attacks

**eTable 17. Summary of Relative and Attributable Risk of AESI Following MRNA-1273.222 COVID-19 Vaccination in the Primary SCCS Analysis Stratified by Age, Adjusting for Event Dependent Observation Time**

|  | **MRNA-1273.222** | | | | | | | | | | | | | | |
| --- | --- | --- | --- | --- | --- | --- | --- | --- | --- | --- | --- | --- | --- | --- | --- |
|  | **65-74** | | | | | **75-84** | | | | | **85+** | | | | |
| **NHS** | | | | | | | | | | | | | | | |
| **1-21 Day Risk Window** | | | | | | | | | | | | | | | |
| ***Conditional Poisson Regression*** | | | | | | | | | | | | | | | |
| *Cases in Risk Window* | 65 | | | | | 89 | | | | | 61 | | | | |
| *Cases in Control Window* | 189 | | | | | 265 | | | | | 153 | | | | |
| *Risk Days* | 7,388 | | | | | 9,954 | | | | | 5,517 | | | | |
| *Control Days* | 16,066 | | | | | 21,484 | | | | | 11,599 | | | | |
| *Model Results* | RR | 95% CI | | P-Value | SE | RR | 95% CI | | P-Value | SE | RR | 95% CI | | P-Value | SE |
|  | 0.85 | 0.64 | 1.13 | 0.26 | 0.15 | 0.89 | 0.70 | 1.13 | 0.34 | 0.12 | 0.91 | 0.66 | 1.25 | 0.55 | 0.16 |
| ***Attributable Risk*** | | | | | | | | | | | | | | | |
| *# of Eligible Vaccinated* | 849,710 | | | | | 613,271 | | | | | 178,091 | | | | |
| *Person Years* | 35,637,049 | | | | | 25,713,934 | | | | | 7,455,719 | | | | |
| *Attributable Risk per 100,000 Doses* | AR | 95% CI | | P-Value | SE | AR | 95% CI | | P-Value | SE | AR | 95% CI | | P-Value | SE |
|  | -1.37 | -3.67 | 0.94 | 0.24 | 1.18 | -1.82 | -5.44 | 1.79 | 0.32 | 1.84 | -3.45 | -14.49 | 7.58 | 0.54 | 5.63 |
| *Attributable Risk per 100,000 Person Years* | AR | 95% CI | | P-Value | SE | AR | 95% CI | | P-Value | SE | AR | 95% CI | | P-Value | SE |
|  | -11.91 | -31.97 | 8.15 | 0.24 | 10.23 | -15.86 | -47.34 | 15.61 | 0.32 | 16.06 | -30.10 | -126.30 | 66.10 | 0.54 | 49.08 |
| **22-42 Day Risk Window** | | | | | | | | | | | | | | | |
| ***Conditional Poisson Regression*** | | | | | | | | | | | | | | | |
| *Cases in Risk Window* | 98 | | | | | 120 | | | | | 50 | | | | |
| *Cases in Control Window* | 189 | | | | | 265 | | | | | 153 | | | | |
| *Risk Days* | 7,315 | | | | | 9,923 | | | | | 5,357 | | | | |
| *Control Days* | 16,066 | | | | | 21,484 | | | | | 11,599 | | | | |
| *Model Results* | RR | 95% CI | | P-Value | SE | RR | 95% CI | | P-Value | SE | RR | 95% CI | | P-Value | SE |
|  | 1.24 | 0.96 | 1.58 | 0.09 | 0.13 | 1.11 | 0.89 | 1.38 | 0.36 | 0.11 | 0.77 | 0.55 | 1.06 | 0.11 | 0.17 |
| ***Attributable Risk*** | | | | | | | | | | | | | | | |
| *# of Eligible Vaccinated* | 849,710 | | | | | 613,271 | | | | | 178,091 | | | | |
| *Person Years* | 35,637,049 | | | | | 25,713,934 | | | | | 7,455,719 | | | | |
| *Attributable Risk per 100,000 Doses* | AR | 95% CI | | P-Value | SE | AR | 95% CI | | P-Value | SE | AR | 95% CI | | P-Value | SE |
|  | 2.21 | -0.43 | 4.85 | 0.10 | 1.35 | 1.91 | -2.20 | 6.01 | 0.36 | 2.10 | -8.61 | -18.65 | 1.43 | 0.09 | 5.12 |
| *Attributable Risk per 100,000 Person Years* | AR | 95% CI | | P-Value | SE | AR | 95% CI | | P-Value | SE | AR | 95% CI | | P-Value | SE |
|  | 19.20 | -3.78 | 42.19 | 0.10 | 11.73 | 16.59 | -19.16 | 52.34 | 0.36 | 18.24 | -75.08 | -162.60 | 12.45 | 0.09 | 44.66 |
| **TIA** |  |  |  |  |  |  |  |  |  |  |  |  |  |  |  |
| **1-21 Day Risk Window** | | | | | | | | | | | | | | | |
| ***Conditional Poisson Regression*** | | | | | | | | | | | | | | | |
| *Cases in Risk Window* | 70 | | | | | 119 | | | | | 56 | | | | |
| *Cases in Control Window* | 163 | | | | | 271 | | | | | 102 | | | | |
| *Risk Days* | 6,615 | | | | | 10,646 | | | | | 4,468 | | | | |
| *Control Days* | 14,900 | | | | | 24,106 | | | | | 9,952 | | | | |
| *Model Results* | RR | 95% CI | | P-Value | SE | RR | 95% CI | | P-Value | SE | RR | 95% CI | | P-Value | SE |
|  | 1.04 | 0.79 | 1.38 | 0.78 | 0.14 | 1.05 | 0.84 | 1.30 | 0.67 | 0.11 | 1.29 | 0.93 | 1.79 | 0.13 | 0.17 |
| ***Attributable Risk*** | | | | | | | | | | | | | | | |
| *# of Eligible Vaccinated* | 1,144,791 | | | | | 828,400 | | | | | 250,661 | | | | |
| *Person Years* | 48,013,563 | | | | | 34,734,511 | | | | | 10,493,283 | | | | |
| *Attributable Risk per 100,000 Doses* | AR | 95% CI | | P-Value | SE | AR | 95% CI | | P-Value | SE | AR | 95% CI | | P-Value | SE |
|  | 0.24 | -1.46 | 1.94 | 0.78 | 0.87 | 0.67 | -2.40 | 3.73 | 0.67 | 1.56 | 5.03 | -1.75 | 11.80 | 0.15 | 3.45 |
| *Attributable Risk per 100,000 Person Years* | AR | 95% CI | | P-Value | SE | AR | 95% CI | | P-Value | SE | AR | 95% CI | | P-Value | SE |
|  | 2.12 | -12.67 | 16.91 | 0.78 | 7.54 | 5.82 | -20.86 | 32.49 | 0.67 | 13.61 | 43.82 | -15.22 | 102.86 | 0.15 | 30.12 |
| **22-42 Day Risk Window** | | | | | | | | | | | | | | | |
| ***Conditional Poisson Regression*** | | | | | | | | | | | | | | | |
| *Cases in Risk Window* | 82 | | | | | 117 | | | | | 55 | | | | |
| *Cases in Control Window* | 163 | | | | | 271 | | | | | 102 | | | | |
| *Risk Days* | 6,610 | | | | | 10,626 | | | | | 4,437 | | | | |
| *Control Days* | 14,900 | | | | | 24,106 | | | | | 9,952 | | | | |
| *Model Results* | RR | 95% CI | | P-Value | SE | RR | 95% CI | | P-Value | SE | RR | 95% CI | | P-Value | SE |
|  | 1.21 | 0.93 | 1.58 | 0.16 | 0.14 | 1.03 | 0.83 | 1.27 | 0.81 | 0.11 | 1.27 | 0.92 | 1.77 | 0.15 | 0.17 |
| ***Attributable Risk*** | | | | | | | | | | | | | | | |
| *# of Eligible Vaccinated* | 1,144,791 | | | | | 828,400 | | | | | 250,661 | | | | |
| *Person Years* | 48,013,563 | | | | | 34,734,511 | | | | | 10,493,283 | | | | |
| *Attributable Risk per 100,000 Doses* | AR | 95% CI | | P-Value | SE | AR | 95% CI | | P-Value | SE | AR | 95% CI | | P-Value | SE |
|  | 1.25 | -0.54 | 3.05 | 0.17 | 0.91 | 0.36 | -2.63 | 3.35 | 0.81 | 1.53 | 4.71 | -1.83 | 11.24 | 0.16 | 3.33 |
| *Attributable Risk per 100,000 Person Years* | AR | 95% CI | | P-Value | SE | AR | 95% CI | | P-Value | SE | AR | 95% CI | | P-Value | SE |
|  | 10.91 | -4.69 | 26.52 | 0.17 | 7.96 | 3.15 | -22.89 | 29.19 | 0.81 | 13.29 | 41.03 | -15.95 | 98.01 | 0.16 | 29.07 |
| **NHS/TIA** | | | | | | | | | | | | | | | |
| **1-21 Day Risk Window** | | | | | | | | | | | | | | | |
| ***Conditional Poisson Regression*** | | | | | | | | | | | | | | | |
| *Cases in Risk Window* | 117 | | | | | 173 | | | | | 104 | | | | |
| *Cases in Control Window* | 306 | | | | | 453 | | | | | 218 | | | | |
| *Risk Days* | 12,155 | | | | | 17,430 | | | | | 8,410 | | | | |
| *Control Days* | 26,874 | | | | | 38,492 | | | | | 18,110 | | | | |
| *Model Results* | RR | 95% CI | | P-Value | SE | RR | 95% CI | | P-Value | SE | RR | 95% CI | | P-Value | SE |
|  | 0.94 | 0.76 | 1.17 | 0.58 | 0.11 | 0.95 | 0.80 | 1.14 | 0.60 | 0.09 | 1.10 | 0.87 | 1.41 | 0.43 | 0.12 |
| ***Attributable Risk*** | | | | | | | | | | | | | | | |
| *# of Eligible Vaccinated* | 849,710 | | | | | 613,271 | | | | | 178,091 | | | | |
| *Person Years* | 35,637,049 | | | | | 25,713,934 | | | | | 7,455,719 | | | | |
| *Attributable Risk per 100,000 Doses* | AR | 95% CI | | P-Value | SE | AR | 95% CI | | P-Value | SE | AR | 95% CI | | P-Value | SE |
|  | -0.87 | -3.92 | 2.18 | 0.58 | 1.56 | -1.34 | -6.31 | 3.63 | 0.60 | 2.54 | 5.49 | -8.19 | 19.17 | 0.43 | 6.98 |
| *Attributable Risk per 100,000 Person Years* | AR | 95% CI | | P-Value | SE | AR | 95% CI | | P-Value | SE | AR | 95% CI | | P-Value | SE |
|  | -7.57 | -34.12 | 18.97 | 0.58 | 13.54 | -11.66 | -54.96 | 31.64 | 0.60 | 22.09 | 47.86 | -71.41 | 167.13 | 0.43 | 60.86 |
| **22-42 Day Risk Window** | | | | | | | | | | | | | | | |
| ***Conditional Poisson Regression*** | | | | | | | | | | | | | | | |
| *Cases in Risk Window* | 156 | | | | | 204 | | | | | 80 | | | | |
| *Cases in Control Window* | 306 | | | | | 453 | | | | | 218 | | | | |
| *Risk Days* | 12,082 | | | | | 17,399 | | | | | 8,219 | | | | |
| *Control Days* | 26,874 | | | | | 38,492 | | | | | 18,110 | | | | |
| *Model Results* | RR | 95% CI | | P-Value | SE | RR | 95% CI | | P-Value | SE | RR | 95% CI | | P-Value | SE |
|  | 1.22 | 1.01 | 1.49 | 0.04 | 0.10 | 1.08 | 0.92 | 1.28 | 0.35 | 0.08 | 0.87 | 0.67 | 1.13 | 0.29 | 0.13 |
| ***Attributable Risk*** | | | | | | | | | | | | | | | |
| *# of Eligible Vaccinated* | 849,710 | | | | | 613,271 | | | | | 178,091 | | | | |
| *Person Years* | 35,637,049 | | | | | 25,713,934 | | | | | 7,455,719 | | | | |
| *Attributable Risk per 100,000 Doses* | AR | 95% CI | | P-Value | SE | AR | 95% CI | | P-Value | SE | AR | 95% CI | | P-Value | SE |
|  | 3.37 | 0.05 | 6.69 | 0.05 | 1.69 | 2.55 | -2.81 | 7.92 | 0.35 | 2.74 | -6.75 | -19.13 | 5.63 | 0.29 | 6.32 |
| *Attributable Risk per 100,000 Person Years* | AR | 95% CI | | P-Value | SE | AR | 95% CI | | P-Value | SE | AR | 95% CI | | P-Value | SE |
|  | 29.32 | 0.46 | 58.18 | 0.05 | 14.73 | 22.23 | -24.46 | 68.92 | 0.35 | 23.82 | -58.86 | -166.82 | 49.09 | 0.29 | 55.08 |
| **HS** | | | | | | | | | | | | | | | |
| **1-21 Day Risk Window** | | | | | | | | | | | | | | | |
| ***Conditional Poisson Regression*** | | | | | | | | | | | | | | | |
| *Cases in Risk Window* | 16 | | | | | 18 | | | | | 16 | | | | |
| *Cases in Control Window* | 67 | | | | | 66 | | | | | 39 | | | | |
| *Risk Days* | 2,214 | | | | | 2,139 | | | | | 1,538 | | | | |
| *Control Days* | 4,482 | | | | | 4,327 | | | | | 2,931 | | | | |
| *Model Results* | RR | 95% CI | | P-Value | SE | RR | 95% CI | | P-Value | SE | RR | 95% CI | | P-Value | SE |
|  | 0.56 | 0.32 | 1.00 | 0.05 | 0.29 | 0.69 | 0.39 | 1.21 | 0.20 | 0.29 | 1.19 | 0.64 | 2.24 | 0.58 | 0.32 |
| ***Attributable Risk*** | | | | | | | | | | | | | | |  |
| *# of Eligible Vaccinated* | 727,608 | | | | | 523,327 | | | | | 148,356 | | | | |
| *Person Years* | 30,516,235 | | | | | 21,943,175 | | | | | 6,211,289 | | | | |
| *Attributable Risk per 100,000 Doses* | AR | 95% CI | | P-Value | SE | AR | 95% CI | | P-Value | SE | AR | 95% CI | | P-Value | SE |
|  | -1.72 | -3.33 | -0.11 | 0.04 | 0.82 | -1.54 | -3.82 | 0.75 | 0.19 | 1.16 | 1.75 | -4.57 | 8.08 | 0.59 | 3.23 |
| *Attributable Risk per 100,000 Person Years* | AR | 95% CI | | P-Value | SE | AR | 95% CI | | P-Value | SE | AR | 95% CI | | P-Value | SE |
|  | -14.94 | -28.97 | -0.92 | 0.04 | 7.16 | -13.37 | -33.23 | 6.49 | 0.19 | 10.13 | 15.28 | -39.88 | 70.45 | 0.59 | 28.14 |
| **22-42 Day Risk Window** | | | | | | | | | | | | | | | |
| ***Conditional Poisson Regression*** | | | | | | | | | | | | | | | |
| *Cases in Risk Window* | 23 | | | | | 19 | | | | | 19 | | | | |
| *Cases in Control Window* | 67 | | | | | 66 | | | | | 39 | | | | |
| *Risk Days* | 2,150 | | | | | 2,090 | | | | | 1,491 | | | | |
| *Control Days* | 4,482 | | | | | 4,327 | | | | | 2,931 | | | | |
| *Model Results* | RR | 95% CI | | P-Value | SE | RR | 95% CI | | P-Value | SE | RR | 95% CI | | P-Value | SE |
|  | 0.73 | 0.44 | 1.21 | 0.22 | 0.26 | 0.69 | 0.40 | 1.17 | 0.17 | 0.27 | 1.24 | 0.69 | 2.22 | 0.48 | 0.30 |
| ***Attributable Risk*** | | | | | | | | | | | | | | | |
| *# of Eligible Vaccinated* | 727,608 | | | | | 523,327 | | | | | 148,356 | | | | |
| *Person Years* | 30,516,235 | | | | | 21,943,175 | | | | | 6,211,289 | | | | |
| *Attributable Risk per 100,000 Doses* | AR | 95% CI | | P-Value | SE | AR | 95% CI | | P-Value | SE | AR | 95% CI | | P-Value | SE |
|  | -1.19 | -3.06 | 0.69 | 0.22 | 0.96 | -1.67 | -4.06 | 0.73 | 0.17 | 1.22 | 2.45 | -4.26 | 9.16 | 0.47 | 3.42 |
| *Attributable Risk per 100,000 Person Years* | AR | 95% CI | | P-Value | SE | AR | 95% CI | | P-Value | SE | AR | 95% CI | | P-Value | SE |
|  | -10.32 | -26.64 | 6.00 | 0.22 | 8.33 | -14.51 | -35.35 | 6.33 | 0.17 | 10.63 | 21.34 | -37.16 | 79.83 | 0.47 | 29.85 |

**Abbreviations:** BNT162b2; WT/OMI BA.4/BA.5, Pfizer COVID-19 BivalentmRNA-1273.222, Moderna COVID-19 Bivalent;

HS, hemorrhagic stroke; NHS, non-hemorrhagic stroke; TIA, transient ischemic attacks

**eTable 18. Summary of Relative and Attributable Risk of AESI Following BNT162b2; WT/OMI BA.4/BA.5 COVID-19 Vaccination in the Primary SCCS Analysis Stratified by Concomitant Vaccination Status, Adjusting for Event Dependent Observation Time**

|  | **BNT162b2; WT/OMI BA.4/BA.5** | | | | | | | | | |
| --- | --- | --- | --- | --- | --- | --- | --- | --- | --- | --- |
|  | **Concomitant High Dose or Adjuvanted Influenza Vaccination** | | | | | **No Concomitant** | | | | |
| **NHS** | | | | | | | | | | |
| **1-21 Day Risk Window** | | | | | | | | | | |
| ***Conditional Poisson Regression*** | | | | | | | | | | |
| *Cases in Risk Window* | 201 | | | | | 215 | | | | |
| *Cases in Control Window* | 397 | | | | | 541 | | | | |
| *Risk Days* | 16,639 | | | | | 20,214 | | | | |
| *Control Days* | 35,068 | | | | | 43,153 | | | | |
| *Model Results* | RR | 95% CI | | P-Value | SE | RR | 95% CI | | P-Value | SE |
|  | 1.19 | 1.00 | 1.42 | 0.05 | 0.09 | 0.95 | 0.81 | 1.12 | 0.55 | 0.08 |
| ***Attributable Risk*** | | | | | | | | | | |
| *# of Eligible Vaccinated* | 1,042,059 | | | | | 1,377,536 | | | | |
| *Person Years* | 43,683,922 | | | | | 57,748,729 | | | | |
| *Attributable Risk per 100,000 Doses* | AR | 95% CI | | P-Value | SE | AR | 95% CI | | P-Value | SE |
|  | 3.13 | -0.04 | 6.30 | 0.05 | 1.62 | -0.81 | -3.43 | 1.80 | 0.54 | 1.34 |
| *Attributable Risk per 100,000 Person Years* | AR | 95% CI | | P-Value | SE | AR | 95% CI | | P-Value | SE |
|  | 27.28 | -0.31 | 54.87 | 0.05 | 14.08 | -7.09 | -29.88 | 15.70 | 0.54 | 11.63 |
| **22-42 Day Risk Window** | | | | | | | | | | |
| ***Conditional Poisson Regression*** | | | | | | | | | | |
| *Cases in Risk Window* | 199 | | | | | 211 | | | | |
| *Cases in Control Window* | 397 | | | | | 541 | | | | |
| *Risk Days* | 16,298 | | | | | 19,860 | | | | |
| *Control Days* | 35,068 | | | | | 43,153 | | | | |
| *Model Results* | RR | 95% CI | | P-Value | SE | RR | 95% CI | | P-Value | SE |
|  | 1.20 | 1.01 | 1.42 | 0.04 | 0.09 | 0.94 | 0.80 | 1.10 | 0.43 | 0.08 |
| ***Attributable Risk*** | | | | | | | | | | |
| *# of Eligible Vaccinated* | 1,042,059 | | | | | 1,377,536 | | | | |
| *Person Years* | 43,683,922 | | | | | 57,748,729 | | | | |
| *Attributable Risk per 100,000 Doses* | AR | 95% CI | | P-Value | SE | AR | 95% CI | | P-Value | SE |
|  | 3.13 | 0.05 | 6.22 | 0.05 | 1.57 | -1.03 | -3.54 | 1.48 | 0.42 | 1.28 |
| *Attributable Risk per 100,000 Person Years* | AR | 95% CI | | P-Value | SE | AR | 95% CI | | P-Value | SE |
|  | 27.29 | 0.44 | 54.14 | 0.05 | 13.70 | -8.99 | -30.84 | 12.87 | 0.42 | 11.15 |
| **TIA** | | | | | | | | | | |
| **1-21 Day Risk Window** | | | | | | | | | | |
| ***Conditional Poisson Regression*** | | | | | | | | | | |
| *Cases in Risk Window* | 169 | | | | | 214 | | | | |
| *Cases in Control Window* | 379 | | | | | 457 | | | | |
| *Risk Days* | 14,769 | | | | | 18,388 | | | | |
| *Control Days* | 33,339 | | | | | 41,548 | | | | |
| *Model Results* | RR | 95% CI | | P-Value | SE | RR | 95% CI | | P-Value | SE |
|  | 1.06 | 0.88 | 1.27 | 0.52 | 0.09 | 1.11 | 0.95 | 1.31 | 0.19 | 0.08 |
| ***Attributable Risk*** | | | | | | | | | | |
| *# of Eligible Vaccinated* | 1,294,240 | | | | | 1,839,657 | | | | |
| *Person Years* | 54,253,379 | | | | | 77,123,329 | | | | |
| *Attributable Risk per 100,000 Doses* | AR | 95% CI | | P-Value | SE | AR | 95% CI | | P-Value | SE |
|  | 0.75 | -1.59 | 3.09 | 0.53 | 1.19 | 1.19 | -0.64 | 3.02 | 0.20 | 0.93 |
| *Attributable Risk per 100,000 Person Years* | AR | 95% CI | | P-Value | SE | AR | 95% CI | | P-Value | SE |
|  | 6.55 | -13.83 | 26.94 | 0.53 | 10.40 | 10.35 | -5.60 | 26.30 | 0.20 | 8.14 |
| **22-42 Day Risk Window** | | | | | | | | | | |
| ***Conditional Poisson Regression*** | | | | | | | | | | |
| *Cases in Risk Window* | 156 | | | | | 205 | | | | |
| *Cases in Control Window* | 379 | | | | | 457 | | | | |
| *Risk Days* | 14,725 | | | | | 18,372 | | | | |
| *Control Days* | 33,339 | | | | | 41,548 | | | | |
| *Model Results* | RR | 95% CI | | P-Value | SE | RR | 95% CI | | P-Value | SE |
|  | 0.98 | 0.81 | 1.18 | 0.83 | 0.10 | 1.06 | 0.90 | 1.25 | 0.50 | 0.08 |
| ***Attributable Risk*** | | | | | | | | | | |
| *# of Eligible Vaccinated* | 1,294,240 | | | | | 1,839,657 | | | | |
| *Person Years* | 54,253,379 | | | | | 77,123,329 | | | | |
| *Attributable Risk per 100,000 Doses* | AR | 95% CI | | P-Value | SE | AR | 95% CI | | P-Value | SE |
|  | -0.25 | -2.47 | 1.97 | 0.83 | 1.13 | 0.62 | -1.19 | 2.43 | 0.50 | 0.93 |
| *Attributable Risk per 100,000 Person Years* | AR | 95% CI | | P-Value | SE | AR | 95% CI | | P-Value | SE |
|  | -2.18 | -21.54 | 17.18 | 0.83 | 9.88 | 5.40 | -10.40 | 21.20 | 0.50 | 8.06 |
| **NHS/TIA** | | | | | | | | | | |
| **1-21 Day Risk Window** | | | | | | | | | | |
| ***Conditional Poisson Regression*** | | | | | | | | | | |
| *Cases in Risk Window* | 329 | | | | | 372 | | | | |
| *Cases in Control Window* | 688 | | | | | 868 | | | | |
| *Risk Days* | 27,796 | | | | | 33,520 | | | | |
| *Control Days* | 60,307 | | | | | 73,439 | | | | |
| *Model Results* | RR | 95% CI | | P-Value | SE | RR | 95% CI | | P-Value | SE |
|  | 1.13 | 0.98 | 1.29 | 0.09 | 0.07 | 1.02 | 0.90 | 1.15 | 0.79 | 0.06 |
| ***Attributable Risk*** | | | | | | | | | | |
| *# of Eligible Vaccinated* | 1,042,059 | | | | | 1,377,536 | | | | |
| *Person Years* | 43,683,922 | | | | | 57,748,729 | | | | |
| *Attributable Risk per 100,000 Doses* | AR | 95% CI | | P-Value | SE | AR | 95% CI | | P-Value | SE |
|  | 3.52 | -0.53 | 7.57 | 0.09 | 2.07 | 0.45 | -2.86 | 3.76 | 0.79 | 1.69 |
| *Attributable Risk per 100,000 Person Years* | AR | 95% CI | | P-Value | SE | AR | 95% CI | | P-Value | SE |
|  | 30.65 | -4.65 | 65.95 | 0.09 | 18.01 | 3.94 | -24.88 | 32.77 | 0.79 | 14.71 |
| **22-42 Day Risk Window** | | | | | | | | | | |
| ***Conditional Poisson Regression*** | | | | | | | | | | |
| *Cases in Risk Window* | 312 | | | | | 361 | | | | |
| *Cases in Control Window* | 688 | | | | | 868 | | | | |
| *Risk Days* | 27,426 | | | | | 33,153 | | | | |
| *Control Days* | 60,307 | | | | | 73,439 | | | | |
| *Model Results* | RR | 95% CI | | P-Value | SE | RR | 95% CI | | P-Value | SE |
|  | 1.08 | 0.94 | 1.24 | 0.26 | 0.07 | 0.99 | 0.87 | 1.12 | 0.87 | 0.06 |
| ***Attributable Risk*** | | | | | | | | | | |
| *# of Eligible Vaccinated* | 1,042,059 | | | | | 1,377,536 | | | | |
| *Person Years* | 43,683,922 | | | | | 57,748,729 | | | | |
| *Attributable Risk per 100,000 Doses* | AR | 95% CI | | P-Value | SE | AR | 95% CI | | P-Value | SE |
|  | 2.23 | -1.78 | 6.25 | 0.28 | 2.05 | -0.28 | -3.55 | 2.98 | 0.87 | 1.67 |
| *Attributable Risk per 100,000 Person Years* | AR | 95% CI | | P-Value | SE | AR | 95% CI | | P-Value | SE |
|  | 19.46 | -15.49 | 54.40 | 0.28 | 17.83 | -2.45 | -30.88 | 25.99 | 0.87 | 14.51 |
| **HS** | | | | | | | | | | |
| **1-21 Day Risk Window** | | | | | | | | | | |
| ***Conditional Poisson Regression*** | | | | | | | | | | |
| *Cases in Risk Window* | 51 | | | | | 56 | | | | |
| *Cases in Control Window* | 128 | | | | | 160 | | | | |
| *Risk Days* | 5,064 | | | | | 5,633 | | | | |
| *Control Days* | 9,282 | | | | | 10,467 | | | | |
| *Model Results* | RR | 95% CI | | P-Value | SE | RR | 95% CI | | P-Value | SE |
|  | 0.96 | 0.67 | 1.39 | 0.85 | 0.19 | 0.85 | 0.60 | 1.20 | 0.35 | 0.17 |
| ***Attributable Risk*** | | | | | | | | | | |
| *# of Eligible Vaccinated* | 901,470 | | | | | 1,193,551 | | | | |
| *Person Years* | 37,790,852 | | | | | 50,035,935 | | | | |
| *Attributable Risk per 100,000 Doses* | AR | 95% CI | | P-Value | SE | AR | 95% CI | | P-Value | SE |
|  | -0.21 | -2.37 | 1.95 | 0.85 | 1.10 | -0.83 | -2.61 | 0.96 | 0.36 | 0.91 |
| *Attributable Risk per 100,000 Person Years* | AR | 95% CI | | P-Value | SE | AR | 95% CI | | P-Value | SE |
|  | -1.80 | -20.61 | 17.02 | 0.85 | 9.60 | -7.22 | -22.76 | 8.32 | 0.36 | 7.93 |
| **22-42 Day Risk Window** | | | | | | | | | | |
| ***Conditional Poisson Regression*** | | | | | | | | | | |
| *Cases in Risk Window* | 66 | | | | | 55 | | | | |
| *Cases in Control Window* | 128 | | | | | 160 | | | | |
| *Risk Days* | 4,762 | | | | | 5,345 | | | | |
| *Control Days* | 9,282 | | | | | 10,467 | | | | |
| *Model Results* | RR | 95% CI | | P-Value | SE | RR | 95% CI | | P-Value | SE |
|  | 1.26 | 0.92 | 1.74 | 0.16 | 0.16 | 0.76 | 0.54 | 1.06 | 0.11 | 0.17 |
| ***Attributable Risk*** | | | | | | | | | | |
| *# of Eligible Vaccinated* | 901,470 | | | | | 1,193,551 | | | | |
| *Person Years* | 37,790,852 | | | | | 50,035,935 | | | | |
| *Attributable Risk per 100,000 Doses* | AR | 95% CI | | P-Value | SE | AR | 95% CI | | P-Value | SE |
|  | 1.51 | -0.63 | 3.65 | 0.17 | 1.09 | -1.48 | -3.25 | 0.29 | 0.10 | 0.90 |
| *Attributable Risk per 100,000 Person Years* | AR | 95% CI | | P-Value | SE | AR | 95% CI | | P-Value | SE |
|  | 13.17 | -5.44 | 31.79 | 0.17 | 9.50 | -12.89 | -28.30 | 2.52 | 0.10 | 7.86 |

**Abbreviations:** BNT162b2; WT/OMI BA.4/BA.5, Pfizer COVID-19 BivalentmRNA-1273.222, Moderna COVID-19 Bivalent;

HS, hemorrhagic stroke; NHS, non-hemorrhagic stroke; TIA, transient ischemic attacks

**eTable 19. Summary of Relative and Attributable Risk of AESI Following MRNA-1273.222 COVID-19 Vaccination in the Primary SCCS Analysis Stratified by Concomitant Vaccination Status, Adjusting for Event Dependent Observation Time**

|  | **MRNA-1273.222** | | | | | | | | | |
| --- | --- | --- | --- | --- | --- | --- | --- | --- | --- | --- |
|  | **Concomitant High Dose or Adjuvanted Influenza Vaccination** | | | | | **No Concomitant** | | | | |
| **NHS** |  |  |  |  |  |  |  |  |  |  |
| **1-21 Day Risk Window** |  |  |  |  |  |  |  |  |  |  |
| ***Conditional Poisson Regression*** |  |  |  |  |  |  |  |  |  |  |
| *Cases in Risk Window* | 88 | | | | | 124 | | | | |
| *Cases in Control Window* | 243 | | | | | 354 | | | | |
| *Risk Days* | 9,054 | | | | | 13,490 | | | | |
| *Control Days* | 19,079 | | | | | 29,354 | | | | |
| *Model Results* | RR | 95% CI | | P-Value | SE | RR | 95% CI | | P-Value | SE |
|  | 0.89 | 0.69 | 1.14 | 0.35 | 0.13 | 0.88 | 0.71 | 1.08 | 0.22 | 0.11 |
| ***Attributable Risk*** |  |  |  |  |  |  |  |  |  |  |
| *# of Eligible Vaccinated* | 599,304 | | | | | 1,023,631 | | | | |
| *Person Years* | 25,127,676 | | | | | 42,919,658 | | | | |
| *Attributable Risk per 100,000 Doses* | AR | 95% CI | | P-Value | SE | AR | 95% CI | | P-Value | SE |
|  | -1.87 | -5.71 | 1.96 | 0.34 | 1.96 | -1.69 | -4.28 | 0.90 | 0.20 | 1.32 |
| *Attributable Risk per 100,000 Person Years* | AR | 95% CI | | P-Value | SE | AR | 95% CI | | P-Value | SE |
|  | -16.30 | -49.68 | 17.08 | 0.34 | 17.03 | -14.74 | -37.29 | 7.80 | 0.20 | 11.50 |
| **22-42 Day Risk Window** |  |  |  |  |  |  |  |  |  |  |
| ***Conditional Poisson Regression*** |  |  |  |  |  |  |  |  |  |  |
| *Cases in Risk Window* | 101 | | | | | 165 | | | | |
| *Cases in Control Window* | 243 | | | | | 354 | | | | |
| *Risk Days* | 8,896 | | | | | 13,384 | | | | |
| *Control Days* | 19,079 | | | | | 29,354 | | | | |
| *Model Results* | RR | 95% CI | | P-Value | SE | RR | 95% CI | | P-Value | SE |
|  | 1.00 | 0.79 | 1.27 | 0.98 | 0.12 | 1.11 | 0.92 | 1.34 | 0.26 | 0.10 |
| ***Attributable Risk*** |  |  |  |  |  |  |  |  |  |  |
| *# of Eligible Vaccinated* | 599,304 | | | | | 1,023,631 | | | | |
| *Person Years* | 25,127,676 | | | | | 42,919,658 | | | | |
| *Attributable Risk per 100,000 Doses* | AR | 95% CI | | P-Value | SE | AR | 95% CI | | P-Value | SE |
|  | 0.04 | -3.90 | 3.97 | 0.98 | 2.01 | 1.64 | -1.28 | 4.56 | 0.27 | 1.49 |
| *Attributable Risk per 100,000 Person Years* | AR | 95% CI | | P-Value | SE | AR | 95% CI | | P-Value | SE |
|  | 0.33 | -33.92 | 34.59 | 0.98 | 17.48 | 14.28 | -11.13 | 39.69 | 0.27 | 12.96 |
| **TIA** |  |  |  |  |  |  |  |  |  |  |
| **1-21 Day Risk Window** |  |  |  |  |  |  |  |  |  |  |
| ***Conditional Poisson Regression*** |  |  |  |  |  |  |  |  |  |  |
| *Cases in Risk Window* | 98 | | | | | 147 | | | | |
| *Cases in Control Window* | 175 | | | | | 355 | | | | |
| *Risk Days* | 7,602 | | | | | 13,938 | | | | |
| *Control Days* | 17,216 | | | | | 31,310 | | | | |
| *Model Results* | RR | 95% CI | | P-Value | SE | RR | 95% CI | | P-Value | SE |
|  | 1.35 | 1.06 | 1.74 | 0.02 | 0.13 | 0.99 | 0.82 | 1.20 | 0.92 | 0.10 |
| ***Attributable Risk*** |  |  |  |  |  |  |  |  |  |  |
| *# of Eligible Vaccinated* | 770,161 | | | | | 1,426,272 | | | | |
| *Person Years* | 32,290,897 | | | | | 59,802,347 | | | | |
| *Attributable Risk per 100,000 Doses* | AR | 95% CI | | P-Value | SE | AR | 95% CI | | P-Value | SE |
|  | 3.33 | 0.46 | 6.20 | 0.02 | 1.46 | -0.11 | -2.11 | 1.90 | 0.92 | 1.02 |
| *Attributable Risk per 100,000 Person Years* | AR | 95% CI | | P-Value | SE | AR | 95% CI | | P-Value | SE |
|  | 28.99 | 4.04 | 53.95 | 0.02 | 12.73 | -0.93 | -18.41 | 16.55 | 0.92 | 8.92 |
| **22-42 Day Risk Window** |  |  |  |  |  |  |  |  |  |  |
| ***Conditional Poisson Regression*** |  |  |  |  |  |  |  |  |  |  |
| *Cases in Risk Window* | 89 | | | | | 162 | | | | |
| *Cases in Control Window* | 175 | | | | | 355 | | | | |
| *Risk Days* | 7,602 | | | | | 13,882 | | | | |
| *Control Days* | 17,216 | | | | | 31,310 | | | | |
| *Model Results* | RR | 95% CI | | P-Value | SE | RR | 95% CI | | P-Value | SE |
|  | 1.21 | 0.94 | 1.56 | 0.14 | 0.13 | 1.09 | 0.90 | 1.31 | 0.37 | 0.09 |
| ***Attributable Risk*** |  |  |  |  |  |  |  |  |  |  |
| *# of Eligible Vaccinated* | 770,161 | | | | | 1,426,272 | | | | |
| *Person Years* | 32,290,897 | | | | | 59,802,347 | | | | |
| *Attributable Risk per 100,000 Doses* | AR | 95% CI | | P-Value | SE | AR | 95% CI | | P-Value | SE |
|  | 2.01 | -0.78 | 4.81 | 0.16 | 1.43 | 0.92 | -1.15 | 3.00 | 0.38 | 1.06 |
| *Attributable Risk per 100,000 Person Years* | AR | 95% CI | | P-Value | SE | AR | 95% CI | | P-Value | SE |
|  | 17.51 | -6.81 | 41.83 | 0.16 | 12.41 | 8.05 | -10.01 | 26.11 | 0.38 | 9.21 |
| **NHS/TIA** |  |  |  |  |  |  |  |  |  |  |
| **1-21 Day Risk Window** |  |  |  |  |  |  |  |  |  |  |
| ***Conditional Poisson Regression*** |  |  |  |  |  |  |  |  |  |  |
| *Cases in Risk Window* | 165 | | | | | 226 | | | | |
| *Cases in Control Window* | 373 | | | | | 592 | | | | |
| *Risk Days* | 14,703 | | | | | 22,872 | | | | |
| *Control Days* | 31,932 | | | | | 50,588 | | | | |
| *Model Results* | RR | 95% CI | | P-Value | SE | RR | 95% CI | | P-Value | SE |
|  | 1.09 | 0.90 | 1.31 | 0.38 | 0.10 | 0.94 | 0.80 | 1.09 | 0.40 | 0.08 |
| ***Attributable Risk*** |  |  |  |  |  |  |  |  |  |  |
| *# of Eligible Vaccinated* | 599,304 | | | | | 1,023,631 | | | | |
| *Person Years* | 25,127,676 | | | | | 42,919,658 | | | | |
| *Attributable Risk per 100,000 Doses* | AR | 95% CI | | P-Value | SE | AR | 95% CI | | P-Value | SE |
|  | 2.19 | -2.79 | 7.18 | 0.39 | 2.54 | -1.50 | -5.02 | 2.01 | 0.40 | 1.79 |
| *Attributable Risk per 100,000 Person Years* | AR | 95% CI | | P-Value | SE | AR | 95% CI | | P-Value | SE |
|  | 19.09 | -24.32 | 62.49 | 0.39 | 22.15 | -13.10 | -43.68 | 17.48 | 0.40 | 15.60 |
| **22-42 Day Risk Window** |  |  |  |  |  |  |  |  |  |  |
| ***Conditional Poisson Regression*** |  |  |  |  |  |  |  |  |  |  |
| *Cases in Risk Window* | 163 | | | | | 272 | | | | |
| *Cases in Control Window* | 373 | | | | | 592 | | | | |
| *Risk Days* | 14,545 | | | | | 22,735 | | | | |
| *Control Days* | 31,932 | | | | | 50,588 | | | | |
| *Model Results* | RR | 95% CI | | P-Value | SE | RR | 95% CI | | P-Value | SE |
|  | 1.06 | 0.88 | 1.27 | 0.57 | 0.09 | 1.10 | 0.95 | 1.27 | 0.20 | 0.07 |
| ***Attributable Risk*** |  |  |  |  |  |  |  |  |  |  |
| *# of Eligible Vaccinated* | 599,304 | | | | | 1,023,631 | | | | |
| *Person Years* | 25,127,676 | | | | | 42,919,658 | | | | |
| *Attributable Risk per 100,000 Doses* | AR | 95% CI | | P-Value | SE | AR | 95% CI | | P-Value | SE |
|  | 1.42 | -3.51 | 6.35 | 0.57 | 2.51 | 2.40 | -1.34 | 6.14 | 0.21 | 1.91 |
| *Attributable Risk per 100,000 Person Years* | AR | 95% CI | | P-Value | SE | AR | 95% CI | | P-Value | SE |
|  | 12.39 | -30.52 | 55.30 | 0.57 | 21.89 | 20.91 | -11.67 | 53.48 | 0.21 | 16.62 |
| **HS** |  |  |  |  |  |  |  |  |  |  |
| **1-21 Day Risk Window** |  |  |  |  |  |  |  |  |  |  |
| ***Conditional Poisson Regression*** |  |  |  |  |  |  |  |  |  |  |
| *Cases in Risk Window* | 18 | | | | | 32 | | | | |
| *Cases in Control Window* | 60 | | | | | 108 | | | | |
| *Risk Days* | 2,034 | | | | | 3,773 | | | | |
| *Control Days* | 4,019 | | | | | 7,529 | | | | |
| *Model Results* | RR | 95% CI | | P-Value | SE | RR | 95% CI | | P-Value | SE |
|  | 0.80 | 0.45 | 1.43 | 0.45 | 0.29 | 0.70 | 0.46 | 1.06 | 0.10 | 0.21 |
| ***Attributable Risk*** |  |  |  |  |  |  |  |  |  |  |
| *# of Eligible Vaccinated* | 510,908 | | | | | 874,210 | | | | |
| *Person Years* | 21,421,683 | | | | | 36,655,897 | | | | |
| *Attributable Risk per 100,000 Doses* | AR | 95% CI | | P-Value | SE | AR | 95% CI | | P-Value | SE |
|  | -0.87 | -3.08 | 1.34 | 0.44 | 1.13 | -1.58 | -3.29 | 0.14 | 0.07 | 0.88 |
| *Attributable Risk per 100,000 Person Years* | AR | 95% CI | | P-Value | SE | AR | 95% CI | | P-Value | SE |
|  | -7.56 | -26.81 | 11.70 | 0.44 | 9.82 | -13.71 | -28.66 | 1.24 | 0.07 | 7.63 |
| **22-42 Day Risk Window** |  |  |  |  |  |  |  |  |  |  |
| ***Conditional Poisson Regression*** |  |  |  |  |  |  |  |  |  |  |
| *Cases in Risk Window* | 20 | | | | | 41 | | | | |
| *Cases in Control Window* | 60 | | | | | 108 | | | | |
| *Risk Days* | 1,981 | | | | | 3,666 | | | | |
| *Control Days* | 4,019 | | | | | 7,529 | | | | |
| *Model Results* | RR | 95% CI | | P-Value | SE | RR | 95% CI | | P-Value | SE |
|  | 0.84 | 0.49 | 1.44 | 0.53 | 0.27 | 0.83 | 0.57 | 1.22 | 0.34 | 0.19 |
| ***Attributable Risk*** |  |  |  |  |  |  |  |  |  |  |
| *# of Eligible Vaccinated* | 510,908 | | | | | 874,210 | | | | |
| *Person Years* | 21,421,683 | | | | | 36,655,897 | | | | |
| *Attributable Risk per 100,000 Doses* | AR | 95% CI | | P-Value | SE | AR | 95% CI | | P-Value | SE |
|  | -0.73 | -3.01 | 1.56 | 0.53 | 1.17 | -0.95 | -2.88 | 0.98 | 0.33 | 0.98 |
| *Attributable Risk per 100,000 Person Years* | AR | 95% CI | | P-Value | SE | AR | 95% CI | | P-Value | SE |
|  | -6.32 | -26.24 | 13.59 | 0.53 | 10.16 | -8.29 | -25.07 | 8.49 | 0.33 | 8.56 |

**Abbreviations:** BNT162b2; WT/OMI BA.4/BA.5, Pfizer COVID-19 BivalentmRNA-1273.222, Moderna COVID-19 Bivalent;

HS, hemorrhagic stroke; NHS, non-hemorrhagic stroke; TIA, transient ischemic attacks

**eTable 20. Summary of Demographics, Socio-Economic Status, Residence, Health Status, and Healthcare Utilization of Influenza Vaccinated Medicare Beneficiaries with Stroke Outcomes in the Secondary Analysis**

| **Patient Characteristics** |  |  |  |  |
| --- | --- | --- | --- | --- |
|  | **NHS** | **TIA** | **NHS/TIA** | **HS** |
| **Total** | **5,497** | **4,871** | **9,065** | **1,498** |
| **Age (years)** | | | | |
| 65-74 | 1,675 (30.47) | 1,475 (30.28) | 2,726 (30.07) | 449 (29.97) |
| 75-84 | 2,332 (42.42) | 2,189 (44.94) | 3,957 (43.65) | 641 (42.79) |
| 85+ | 1,490 (27.11) | 1,207 (24.78) | 2,382 (26.28) | 408 (27.24) |
| Missing/Unknown | 0 (0.00) | 0 (0.00) | 0 (0.00) | 0 (0.00) |
| **Sex** | | | | |
| Female | 3,182 (57.89) | 2,897 (59.47) | 5,326 (58.75) | 771 (51.47) |
| Male | 2,315 (42.11) | 1,974 (40.53) | 3,739 (41.25) | 727 (48.53) |
| Missing/Unknown | 0 (0.00) | 0 (0.00) | 0 (0.00) | 0 (0.00) |
| **Race/Ethnicity** | | | | |
| Asian | 100 (1.82) | 59 (1.21) | 142 (1.57) | 35 (2.34) |
| Black | 300 (5.46) | 195 (4.00) | 446 (4.92) | 65 (4.34) |
| Hispanic | 39 (0.71) | 29 (0.60) | 63 (0.69) | *  (*) |
| Alaskan Native/Native American | 18 (0.33) | 15 (0.31) | 31 (0.34) | *  (*) |
| White | 4,846 (88.16) | 4,420 (90.74) | 8,072 (89.05) | 1,319 (88.05) |
| Other | 98 (1.78) | 59 (1.21) | 142 (1.57) | 39 (2.60) |
| Missing/Unknown | 96 (1.75) | 94 (1.93) | 169 (1.86) | 24 (1.60) |
| **Urban/Rural** | | | | |
| Urban | 4,471 (81.34) | 3,831 (78.65) | 7,297 (80.50) | 1,253 (83.64) |
| Rural | 1,011 (18.39) | 1,023 (21.00) | 1,742 (19.22) | *  (*) |
| Missing/Unknown | 15 (0.27) | 17 (0.35) | 26 (0.29) | *  (*) |
| **HHS Region** | | | | |
| Region 1 | 334 (6.08) | 329 (6.75) | 556 (6.13) | 92 (6.14) |
| Region 2 | 502 (9.13) | 391 (8.03) | 781 (8.62) | 136 (9.08) |
| Region 3 | 724 (13.17) | 607 (12.46) | 1,165 (12.85) | 206 (13.75) |
| Region 4 | 1,149 (20.90) | 1,115 (22.89) | 1,984 (21.89) | 286 (19.09) |
| Region 5 | 985 (17.92) | 773 (15.87) | 1,566 (17.28) | 265 (17.69) |
| Region 6 | 563 (10.24) | 516 (10.59) | 934 (10.30) | 131 (8.74) |
| Region 7 | 307 (5.58) | 257 (5.28) | 490 (5.41) | 78 (5.21) |
| Region 8 | *  (*) | 191 (3.92) | *  (*) | 45 (3.00) |
| Region 9 | 562 (10.22) | 501 (10.29) | 938 (10.35) | 198 (13.22) |
| Region 10 | 209 (3.80) | 187 (3.84) | 348 (3.84) | 61 (4.07) |
| Missing/Unknown | *  (*) | *  (*) | *  (*) | 0 (0.00) |
| **Dual-Eligibility Status**** | | | | |
| Dual-Eligible | 250 (4.55) | 186 (3.82) | 383 (4.23) | 63 (4.21) |
| Non-Dual-Eligible | 5,247 (95.45) | 4,685 (96.18) | 8,682 (95.77) | 1,435 (95.79) |
| **Area Deprivation Index (ADI) Rank** | | | | |
| 1-10 (lowest level of deprivation or disadvantage) | 754 (13.72) | 689 (14.14) | 1,248 (13.77) | 255 (17.02) |
| 11-20 | 762 (13.86) | 668 (13.71) | 1,270 (14.01) | 224 (14.95) |
| 21-30 | 761 (13.84) | 653 (13.41) | 1,250 (13.79) | 212 (14.15) |
| 31-40 | 688 (12.52) | 588 (12.07) | 1,124 (12.40) | 162 (10.81) |
| 41-50 | 622 (11.32) | 564 (11.58) | 1,013 (11.17) | 181 (12.08) |
| 51-60 | 555 (10.10) | 485 (9.96) | 905 (9.98) | 136 (9.08) |
| 61-70 | 445 (8.10) | 396 (8.13) | 734 (8.10) | 113 (7.54) |
| 71-80 | 335 (6.09) | 331 (6.80) | 588 (6.49) | 73 (4.87) |
| 81-90 | 261 (4.75) | 261 (5.36) | 452 (4.99) | 73 (4.87) |
| 91-100 | 163 (2.97) | 115 (2.36) | 244 (2.69) | 33 (2.20) |
| Missing/Unknown | 151 (2.75) | 121 (2.48) | 237 (2.61) | 36 (2.40) |
| **Medicare Status** | | | | |
| Aged-in without ESRD | 5,050 (91.87) | 4,532 (93.04) | 8,369 (92.32) | 1,394 (93.06) |
| Aged & Disabled with ESRD | 11 (0.20) | 11 (0.23) | 17 (0.19) | *  (*) |
| Disabled without ESRD | 436 (7.93) | 328 (6.73) | 679 (7.49) | 99 (6.61) |
| Missing/Unknown | 0 (0.00) | 0 (0.00) | 0 (0.00) | *  (*) |
| **Medical Conditions (0-365 days prior to vaccination date**) | | | | |
| Asthma | 400 (7.28) | 472 (9.69) | 771 (8.51) | 106 (7.08) |
| COPD | 756 (13.75) | 618 (12.69) | 1,222 (13.48) | 235 (15.69) |
| Chronic Kidney Disease | 1,565 (28.47) | 1,281 (26.30) | 2,486 (27.42) | 500 (33.38) |
| Depression | 1,024 (18.63) | 926 (19.01) | 1,721 (18.99) | 277 (18.49) |
| Gout | 345 (6.28) | 290 (5.95) | 553 (6.10) | 110 (7.34) |
| Heart Failure | 828 (15.06) | 645 (13.24) | 1,289 (14.22) | 344 (22.96) |
| Hypercholesterolemia | 1,161 (21.12) | 1,108 (22.75) | 1,954 (21.56) | 333 (22.23) |
| Hypothyroidism | 4,337 (78.90) | 3,802 (78.05) | 7,118 (78.52) | 1,213 (80.97) |
| Hypertension | 1,297 (23.59) | 1,254 (25.74) | 2,217 (24.46) | 356 (23.77) |
| ITP | 15 (0.27) | *  (*) | 24 (0.26) | *  (*) |
| Impaired Mobility | 30 (0.55) | *  (*) | 43 (0.47) | *  (*) |
| Ischemic Heart Disease | 321 (5.84) | 301 (6.18) | 536 (5.91) | 119 (7.94) |
| Nicotine Dependency | 1,290 (23.47) | 1,047 (21.49) | 2,037 (22.47) | 409 (27.30) |
| Obesity | 1,233 (22.43) | 1,091 (22.40) | 2,027 (22.36) | 353 (23.56) |
| **Charlson Comorbidity Index** | | | | |
| 0 | 1,180 (21.47) | 1,138 (23.36) | 2,006 (22.13) | 237 (15.82) |
| 1 | 997 (18.14) | 874 (17.94) | 1,622 (17.89) | 229 (15.29) |
| 2 | 1,010 (18.37) | 852 (17.49) | 1,662 (18.33) | 261 (17.42) |
| 3 | 694 (12.63) | 642 (13.18) | 1,155 (12.74) | 194 (12.95) |
| 4 | 575 (10.46) | 498 (10.22) | 944 (10.41) | 177 (11.82) |
| 5+ | 1,041 (18.94) | 867 (17.80) | 1,676 (18.49) | 400 (26.70) |
| **Prior COVID-19 diagnosis** | | | | |
| In the 30 days prior to outcome | 134 (2.44) | 101 (2.07) | 192 (2.12) | 24 (1.60) |
| In the 31-365 days prior to outcome | 705 (12.83) | 832 (17.08) | 1,298 (14.32) | 219 (14.62) |
| **Prior influenza diagnosis** | | | | |
| In the 30 days prior to outcome | 14 (0.25) | 20 (0.41) | 28 (0.31) | *  (*) |
| In the 31-365 days prior to outcome | 31 (0.56) | 34 (0.70) | 55 (0.61) | *  (*) |
| **Other vaccine (administered in risk/control intervals)** | | | | |
| Pneumococcal vaccine | 152 (2.77) | 146 (3.00) | 262 (2.89) | 31 (2.07) |

**Abbreviations:** BNT162b2; WT/OMI BA.4/BA.5, Pfizer COVID-19 BivalentmRNA-1273.222, Moderna COVID-19 Bivalent;

HS, hemorrhagic stroke; NHS, non-hemorrhagic stroke; TIA, transient ischemic attacks; ADI, area deprivations index; ESRD, end-stage renal disease; COPD, chronic obstructive pulmonary disease; COVID-19, coronavirus 2019

* Outcome counts of 10 or fewer and associated statistics are masked to protect the anonymity of the data.

** Reflects beneficiaries’ dual eligibility for Medicare and Medicaid plan benefits.

**eTable 21. Summary of Relative and Attributable Risk of AESI Following High Dose or Adjuvanted Influenza Vaccination in the Secondary SCCS Analysis, Adjusting for Event Dependent Observation Time**

| **High Dose or Adjuvanted Influenza Vaccine** | | | | | |
| --- | --- | --- | --- | --- | --- |
| **NHS** |  |  |  |  |  |
| **1-21 Day Risk Window** |  |  |  |  |  |
| ***Conditional Poisson Regression*** |  |  |  |  |  |
| *Cases in Risk Window* |  |  | 1,248 |  |  |
| *Cases in Control Window* |  |  | 2,916 |  |  |
| *Risk Days* |  |  | 115,001 |  |  |
| *Control Days* |  |  | 243,497 |  |  |
| *Model Results* | RR | 95% CI | | P-Value | SE |
|  | 1.02 | 0.96 | 1.10 | 0.49 | 0.04 |
| ***Attributable Risk*** |  |  |  |  |  |
| *# of Eligible Vaccinated* |  |  | 6,894,934 |  |  |
| *# of Vaccines* |  |  | 12,445,029 |  |  |
| *Person Years* |  |  | 288,759,941 |  |  |
| *Attributable Risk per 100,000 Doses* | AR | 95% CI | | P-Value | SE |
|  | 0.44 | -0.78 | 1.65 | 0.48 | 0.62 |
| *Attributable Risk per 100,000 Person Years* | AR | 95% CI | | P-Value | SE |
|  | 3.79 | -6.83 | 14.42 | 0.48 | 5.42 |
| **22-42 Day Risk Window** |  |  |  |  |  |
| ***Conditional Poisson Regression*** |  |  |  |  |  |
| *Cases in Risk Window* |  |  | 1,333 |  |  |
| *Cases in Control Window* |  |  | 2,916 |  |  |
| *Risk Days* |  |  | 112,913 |  |  |
| *Control Days* |  |  | 243,497 |  |  |
| *Model Results* | RR | 95% CI | | P-Value | SE |
|  | 1.09 | 1.02 | 1.17 | 0.01 | 0.03 |
| ***Attributable Risk*** |  |  |  |  |  |
| *# of Eligible Vaccinated* |  |  | 6,894,934 |  |  |
| *# of Vaccines* |  |  | 12,445,029 |  |  |
| *Person Years* |  |  | 288,759,941 |  |  |
| *Attributable Risk per 100,000 Doses* | AR | 95% CI | | P-Value | SE |
|  | 1.65 | 0.43 | 2.87 | 0.01 | 0.62 |
| *Attributable Risk per 100,000 Person Years* | AR | 95% CI | | P-Value | SE |
|  | 14.40 | 3.78 | 25.03 | 0.01 | 5.42 |
| **TIA** |  |  |  |  |  |
| **1-21 Day Risk Window** |  |  |  |  |  |
| ***Conditional Poisson Regression*** |  |  |  |  |  |
| *Cases in Risk Window* |  |  | 1,143 |  |  |
| *Cases in Control Window* |  |  | 2,630 |  |  |
| *Risk Days* |  |  | 102,268 |  |  |
| *Control Days* |  |  | 231,047 |  |  |
| *Model Results* | RR | 95% CI | | P-Value | SE |
|  | 1.03 | 0.96 | 1.11 | 0.38 | 0.04 |
| ***Attributable Risk*** |  |  |  |  |  |
| *# of Eligible Vaccinated* |  |  | 9,023,977 |  |  |
| *# of Vaccines* |  |  | 12,445,029 |  |  |
| *Person Years* |  |  | 377,969,554 |  |  |
| *Attributable Risk per 100,000 Doses* | AR | 95% CI | | P-Value | SE |
|  | 0.39 | -0.49 | 1.26 | 0.39 | 0.45 |
| *Attributable Risk per 100,000 Person Years* | AR | 95% CI | | P-Value | SE |
|  | 3.36 | -4.27 | 10.99 | 0.39 | 3.89 |
| **22-42 Day Risk Window** |  |  |  |  |  |
| ***Conditional Poisson Regression*** |  |  |  |  |  |
| *Cases in Risk Window* |  |  | 1,098 |  |  |
| *Cases in Control Window* |  |  | 2,630 |  |  |
| *Risk Days* |  |  | 102,123 |  |  |
| *Control Days* |  |  | 231,047 |  |  |
| *Model Results* | RR | 95% CI | | P-Value | SE |
|  | 0.99 | 0.92 | 1.06 | 0.76 | 0.04 |
| ***Attributable Risk*** |  |  |  |  |  |
| *# of Eligible Vaccinated* |  |  | 9,023,977 |  |  |
| *# of Vaccines* |  |  | 12,445,029 |  |  |
| *Person Years* |  |  | 377,969,554 |  |  |
| *Attributable Risk per 100,000 Doses* | AR | 95% CI | | P-Value | SE |
|  | -0.13 | -1.00 | 0.73 | 0.76 | 0.44 |
| *Attributable Risk per 100,000 Person Years* | AR | 95% CI | | P-Value | SE |
|  | -1.17 | -8.70 | 6.36 | 0.76 | 3.84 |
| **NHS/TIA** |  |  |  |  |  |
| **1-21 Day Risk Window** |  |  |  |  |  |
| ***Conditional Poisson Regression*** |  |  |  |  |  |
| *Cases in Risk Window* |  |  | 2,089 |  |  |
| *Cases in Control Window* |  |  | 4,844 |  |  |
| *Risk Days* |  |  | 189,906 |  |  |
| *Control Days* |  |  | 413,114 |  |  |
| *Model Results* | RR | 95% CI | | P-Value | SE |
|  | 1.03 | 0.98 | 1.09 | 0.25 | 0.03 |
| ***Attributable Risk*** |  |  |  |  |  |
| *# of Eligible Vaccinated* |  |  | 6,894,934 |  |  |
| *# of Vaccines* |  |  | 12,445,029 |  |  |
| *Person Years* |  |  | 288,759,941 |  |  |
| *Attributable Risk per 100,000 Doses* | AR | 95% CI | | P-Value | SE |
|  | 0.91 | -0.63 | 2.46 | 0.24 | 0.79 |
| *Attributable Risk per 100,000 Person Years* | AR | 95% CI | | P-Value | SE |
|  | 7.97 | -5.46 | 21.40 | 0.24 | 6.85 |
| **22-42 Day Risk Window** |  |  |  |  |  |
| ***Conditional Poisson Regression*** |  |  |  |  |  |
| *Cases in Risk Window* |  |  | 2,132 |  |  |
| *Cases in Control Window* |  |  | 4,844 |  |  |
| *Risk Days* |  |  | 187,708 |  |  |
| *Control Days* |  |  | 413,114 |  |  |
| *Model Results* | RR | 95% CI | | P-Value | SE |
|  | 1.05 | 1.00 | 1.11 | 0.06 | 0.03 |
| ***Attributable Risk*** |  |  |  |  |  |
| *# of Eligible Vaccinated* |  |  | 6,894,934 |  |  |
| *# of Vaccines* |  |  | 12,445,029 |  |  |
| *Person Years* |  |  | 288,759,941 |  |  |
| *Attributable Risk per 100,000 Doses* | AR | 95% CI | | P-Value | SE |
|  | 1.49 | -0.08 | 3.06 | 0.06 | 0.80 |
| *Attributable Risk per 100,000 Person Years* | AR | 95% CI | | P-Value | SE |
|  | 13.01 | -0.67 | 26.69 | 0.06 | 6.98 |
| **HS** |  |  |  |  |  |
| **1-21 Day Risk Window** |  |  |  |  |  |
| ***Conditional Poisson Regression*** |  |  |  |  |  |
| *Cases in Risk Window* |  |  | 338 |  |  |
| *Cases in Control Window* |  |  | 808 |  |  |
| *Risk Days* |  |  | 30,873 |  |  |
| *Control Days* |  |  | 56,104 |  |  |
| *Model Results* | RR | 95% CI | | P-Value | SE |
|  | 0.96 | 0.83 | 1.11 | 0.61 | 0.07 |
| ***Attributable Risk*** |  |  |  |  |  |
| *# of Eligible Vaccinated* |  |  | 5,781,464 |  |  |
| *# of Vaccines* |  |  | 12,445,029 |  |  |
| *Person Years* |  |  | 242,109,712 |  |  |
| *Attributable Risk per 100,000 Doses* | AR | 95% CI | | P-Value | SE |
|  | -0.23 | -1.09 | 0.63 | 0.60 | 0.44 |
| *Attributable Risk per 100,000 Person Years* | AR | 95% CI | | P-Value | SE |
|  | -1.99 | -9.47 | 5.50 | 0.60 | 3.82 |
| **22-42 Day Risk Window** |  |  |  |  |  |
| ***Conditional Poisson Regression*** |  |  |  |  |  |
| *Cases in Risk Window* |  |  | 352 |  |  |
| *Cases in Control Window* |  |  | 808 |  |  |
| *Risk Days* |  |  | 28,761 |  |  |
| *Control Days* |  |  | 56,104 |  |  |
| *Model Results* | RR | 95% CI | | P-Value | SE |
|  | 1.05 | 0.92 | 1.20 | 0.49 | 0.07 |
| ***Attributable Risk*** |  |  |  |  |  |
| *# of Eligible Vaccinated* |  |  | 5,781,464 |  |  |
| *# of Vaccines* |  |  | 12,445,029 |  |  |
| *Person Years* |  |  | 242,109,712 |  |  |
| *Attributable Risk per 100,000 Doses* | AR | 95% CI | | P-Value | SE |
|  | 0.29 | -0.52 | 1.09 | 0.49 | 0.41 |
| *Attributable Risk per 100,000 Person Years* | AR | 95% CI | | P-Value | SE |
|  | 2.50 | -4.53 | 9.53 | 0.49 | 3.59 |

**Abbreviations:** BNT162b2; WT/OMI BA.4/BA.5, Pfizer COVID-19 BivalentmRNA-1273.222, Moderna COVID-19 Bivalent;

HS, hemorrhagic stroke; NHS, non-hemorrhagic stroke; TIA, transient ischemic attacks

**eTable 22. Summary of Relative and Attributable Risk of AESI Following High Dose or Adjuvanted Influenza Vaccination in the Secondary SCCS Analysis, Adjusting for Seasonality and Event Dependent Observation Time**

| **High Dose or Adjuvanted Influenza Vaccine** | | | | | |
| --- | --- | --- | --- | --- | --- |
| **NHS** |  |  |  |  |  |
| **1-21 Day Risk Window** |  |  |  |  |  |
| ***Conditional Poisson Regression*** |  |  |  |  |  |
| *Cases in Risk Window* |  |  | 1,248 |  |  |
| *Cases in Control Window* |  |  | 2,916 |  |  |
| *Risk Days* |  |  | 115,001 |  |  |
| *Control Days* |  |  | 243,497 |  |  |
| *Model Results* | RR | 95% CI | | P-Value | SE |
|  | 1.05 | 0.98 | 1.13 | 0.16 | 0.04 |
| ***Attributable Risk*** |  |  |  |  |  |
| *# of Eligible Vaccinated* |  |  | 6,894,934 |  |  |
| *# of Vaccines* |  |  | 12,445,029 |  |  |
| *Person Years* |  |  | 288,759,941 |  |  |
| *Attributable Risk per 100,000 Doses* | AR | 95% CI | | P-Value | SE |
|  | 0.88 | -0.34 | 2.10 | 0.16 | 0.62 |
| *Attributable Risk per 100,000 Person Years* | AR | 95% CI | | P-Value | SE |
|  | 7.68 | -2.95 | 18.32 | 0.16 | 5.43 |
| **22-42 Day Risk Window** |  |  |  |  |  |
| ***Conditional Poisson Regression*** |  |  |  |  |  |
| *Cases in Risk Window* |  |  | 1,333 |  |  |
| *Cases in Control Window* |  |  | 2,916 |  |  |
| *Risk Days* |  |  | 112,913 |  |  |
| *Control Days* |  |  | 243,497 |  |  |
| *Model Results* | RR | 95% CI | | P-Value | SE |
|  | 1.10 | 1.03 | 1.17 | 0.01 | 0.03 |
| ***Attributable Risk*** |  |  |  |  |  |
| *# of Eligible Vaccinated* |  |  | 6,894,934 |  |  |
| *# of Vaccines* |  |  | 12,445,029 |  |  |
| *Person Years* |  |  | 288,759,941 |  |  |
| *Attributable Risk per 100,000 Doses* | AR | 95% CI | | P-Value | SE |
|  | 1.72 | 0.49 | 2.95 | 0.01 | 0.63 |
| *Attributable Risk per 100,000 Person Years* | AR | 95% CI | | P-Value | SE |
|  | 15.00 | 4.26 | 25.73 | 0.01 | 5.47 |
| **TIA** |  |  |  |  |  |
| **1-21 Day Risk Window** |  |  |  |  |  |
| ***Conditional Poisson Regression*** |  |  |  |  |  |
| *Cases in Risk Window* |  |  | 1,143 |  |  |
| *Cases in Control Window* |  |  | 2,630 |  |  |
| *Risk Days* |  |  | 102,268 |  |  |
| *Control Days* |  |  | 231,047 |  |  |
| *Model Results* | RR | 95% CI | | P-Value | SE |
|  | 1.05 | 0.98 | 1.12 | 0.21 | 0.04 |
| ***Attributable Risk*** |  |  |  |  |  |
| *# of Eligible Vaccinated* |  |  | 9,023,977 |  |  |
| *# of Vaccines* |  |  | 12,445,029 |  |  |
| *Person Years* |  |  | 377,969,554 |  |  |
| *Attributable Risk per 100,000 Doses* | AR | 95% CI | | P-Value | SE |
|  | 0.56 | -0.32 | 1.44 | 0.21 | 0.45 |
| *Attributable Risk per 100,000 Person Years* | AR | 95% CI | | P-Value | SE |
|  | 4.87 | -2.80 | 12.54 | 0.21 | 3.91 |
| **22-42 Day Risk Window** |  |  |  |  |  |
| ***Conditional Poisson Regression*** |  |  |  |  |  |
| *Cases in Risk Window* |  |  | 1,098 |  |  |
| *Cases in Control Window* |  |  | 2,630 |  |  |
| *Risk Days* |  |  | 102,123 |  |  |
| *Control Days* |  |  | 231,047 |  |  |
| *Model Results* | RR | 95% CI | | P-Value | SE |
|  | 0.98 | 0.91 | 1.05 | 0.59 | 0.04 |
| ***Attributable Risk*** |  |  |  |  |  |
| *# of Eligible Vaccinated* |  |  | 9,023,977 |  |  |
| *# of Vaccines* |  |  | 12,445,029 |  |  |
| *Person Years* |  |  | 377,969,554 |  |  |
| *Attributable Risk per 100,000 Doses* | AR | 95% CI | | P-Value | SE |
|  | -0.24 | -1.12 | 0.63 | 0.59 | 0.45 |
| *Attributable Risk per 100,000 Person Years* | AR | 95% CI | | P-Value | SE |
|  | -2.10 | -9.74 | 5.53 | 0.59 | 3.90 |
| **NHS/TIA** |  |  |  |  |  |
| **1-21 Day Risk Window** |  |  |  |  |  |
| ***Conditional Poisson Regression*** |  |  |  |  |  |
| *Cases in Risk Window* |  |  | 2,089 |  |  |
| *Cases in Control Window* |  |  | 4,844 |  |  |
| *Risk Days* |  |  | 189,906 |  |  |
| *Control Days* |  |  | 413,114 |  |  |
| *Model Results* | RR | 95% CI | | P-Value | SE |
|  | 1.06 | 1.00 | 1.12 | 0.04 | 0.03 |
| ***Attributable Risk*** |  |  |  |  |  |
| *# of Eligible Vaccinated* |  |  | 6,894,934 |  |  |
| *# of Vaccines* |  |  | 12,445,029 |  |  |
| *Person Years* |  |  | 288,759,941 |  |  |
| *Attributable Risk per 100,000 Doses* | AR | 95% CI | | P-Value | SE |
|  | 1.65 | 0.11 | 3.20 | 0.04 | 0.79 |
| *Attributable Risk per 100,000 Person Years* | AR | 95% CI | | P-Value | SE |
|  | 14.42 | 0.97 | 27.87 | 0.04 | 6.86 |
| **22-42 Day Risk Window** |  |  |  |  |  |
| ***Conditional Poisson Regression*** |  |  |  |  |  |
| *Cases in Risk Window* |  |  | 2,132 |  |  |
| *Cases in Control Window* |  |  | 4,844 |  |  |
| *Risk Days* |  |  | 187,708 |  |  |
| *Control Days* |  |  | 413,114 |  |  |
| *Model Results* | RR | 95% CI | | P-Value | SE |
|  | 1.05 | 1.00 | 1.11 | 0.05 | 0.03 |
| ***Attributable Risk*** |  |  |  |  |  |
| *# of Eligible Vaccinated* |  |  | 6,894,934 |  |  |
| *# of Vaccines* |  |  | 12,445,029 |  |  |
| *Person Years* |  |  | 288,759,941 |  |  |
| *Attributable Risk per 100,000 Doses* | AR | 95% CI | | P-Value | SE |
|  | 1.60 | 0.02 | 3.18 | 0.05 | 0.81 |
| *Attributable Risk per 100,000 Person Years* | AR | 95% CI | | P-Value | SE |
|  | 13.94 | 0.14 | 27.74 | 0.05 | 7.04 |
| **HS** |  |  |  |  |  |
| **1-21 Day Risk Window** |  |  |  |  |  |
| ***Conditional Poisson Regression*** |  |  |  |  |  |
| *Cases in Risk Window* |  |  | 338 |  |  |
| *Cases in Control Window* |  |  | 808 |  |  |
| *Risk Days* |  |  | 30,873 |  |  |
| *Control Days* |  |  | 56,104 |  |  |
| *Model Results* | RR | 95% CI | | P-Value | SE |
|  | 1.01 | 0.88 | 1.17 | 0.85 | 0.07 |
| ***Attributable Risk*** |  |  |  |  |  |
| *# of Eligible Vaccinated* |  |  | 5,781,464 |  |  |
| *# of Vaccines* |  |  | 12,445,029 |  |  |
| *Person Years* |  |  | 242,109,712 |  |  |
| *Attributable Risk per 100,000 Doses* | AR | 95% CI | | P-Value | SE |
|  | 0.08 | -0.76 | 0.92 | 0.85 | 0.43 |
| *Attributable Risk per 100,000 Person Years* | AR | 95% CI | | P-Value | SE |
|  | 0.71 | -6.61 | 8.03 | 0.85 | 3.73 |
| **22-42 Day Risk Window** |  |  |  |  |  |
| ***Conditional Poisson Regression*** |  |  |  |  |  |
| *Cases in Risk Window* |  |  | 352 |  |  |
| *Cases in Control Window* |  |  | 808 |  |  |
| *Risk Days* |  |  | 28,761 |  |  |
| *Control Days* |  |  | 56,104 |  |  |
| *Model Results* | RR | 95% CI | | P-Value | SE |
|  | 1.07 | 0.93 | 1.23 | 0.34 | 0.07 |
| ***Attributable Risk*** |  |  |  |  |  |
| *# of Eligible Vaccinated* |  |  | 5,781,464 |  |  |
| *# of Vaccines* |  |  | 12,445,029 |  |  |
| *Person Years* |  |  | 242,109,712 |  |  |
| *Attributable Risk per 100,000 Doses* | AR | 95% CI | | P-Value | SE |
|  | 0.40 | -0.41 | 1.20 | 0.34 | 0.41 |
| *Attributable Risk per 100,000 Person Years* | AR | 95% CI | | P-Value | SE |
|  | 3.45 | -3.60 | 10.50 | 0.34 | 3.60 |

**Abbreviations:** BNT162b2; WT/OMI BA.4/BA.5, Pfizer COVID-19 BivalentmRNA-1273.222, Moderna COVID-19 Bivalent;

HS, hemorrhagic stroke; NHS, non-hemorrhagic stroke; TIA, transient ischemic attacks

**eTable 23. Summary of Relative and Attributable Risk of NHS Following High Dose or Adjuvanted Influenza Vaccination in the Secondary SCCS Analysis, Adjusting for PPV and Event Dependent Observation Time**

| **High Dose or Adjuvanted Influenza Vaccine** | | | | | |
| --- | --- | --- | --- | --- | --- |
| **NHS** |  |  |  |  |  |
| ***1-21 Day Risk Window*** |  |  |  |  |  |
| ***Conditional Poisson Regression*** |  |  |  |  |  |
| *Cases in Risk Window (1-21days)* | 1,005 | | | | |
| *Cases in Control Window (43-90 days)* | 2,347 | | | | |
| *Risk Days* | 92,574 | | | | |
| *Control Days* | 196,008 | | | | |
| *Model Results* | IRR | 95% CI | | P-Value | SE |
|  | 1.03 | 0.94 | 1.12 | 0.53 | 0.04 |
| ***Attributable Risk*** |  |  |  |  |  |
| *# of Eligible Vaccinated* |  |  | 6,894,934 |  |  |
| *# of Vaccines* |  |  | 12,445,029 |  |  |
| *Person Days* |  |  | 288,759,941 |  |  |
| *Attributable Risk per 100,000 Doses* | AR | 95% CI | | P-Value | SE |
|  | 0.39 | -0.86 | 1.53 | NA | NA |
| *Attributable Risk per 100,000 Person Years* | AR | 95% CI | | P-Value | SE |
|  | 3.36 | -7.46 | 13.30 | NA | NA |
| ***22-42 Day Risk Window*** |  |  |  |  |  |
| ***Conditional Poisson Regression*** |  |  |  |  |  |
| *Cases in Risk Window (22-42days)* |  |  | 1,073 |  |  |
| *Cases in Control Window (43-90 days)* |  |  | 2,347 |  |  |
| *Risk Days* |  |  | 90,892 |  |  |
| *Control Days* |  |  | 196,008 |  |  |
| *Model Results* | IRR | 95% CI | | P-Value | SE |
|  | 1.09 | 1.01 | 1.19 | 0.03 | 0.04 |
| ***Attributable Risk*** |  |  |  |  |  |
| *# of Eligible Vaccinated* |  |  | 6,894,934 |  |  |
| *# of Vaccines* |  |  | 12,445,029 |  |  |
| *Person Days* |  |  | 288,759,941 |  |  |
| *Attributable Risk per 100,000 Doses* | AR | 95% CI | | P-Value | SE |
|  | 1.35 | 0.17 | 2.44 | NA | NA |
| *Attributable Risk per 100,000 Person Years* | AR | 95% CI | | P-Value | SE |
|  | 11.76 | 1.47 | 21.26 | NA | NA |

**Abbreviations:** BNT162b2; WT/OMI BA.4/BA.5, Pfizer COVID-19 BivalentmRNA-1273.222, Moderna COVID-19 Bivalent;

HS, hemorrhagic stroke; NHS, non-hemorrhagic stroke; TIA, transient ischemic attacks

**eTable 24. Summary of Relative and Attributable Risk of AESI Following High Dose or Adjuvanted Influenza Vaccination in the Secondary SCCS Analysis Stratified by Age, Adjusting for Event Dependent Observation Time**

|  | **High Dose or Adjuvanted Influenza Vaccine** | | | | | | | | | | | | | | |
| --- | --- | --- | --- | --- | --- | --- | --- | --- | --- | --- | --- | --- | --- | --- | --- |
|  | **65-74** | | | | | **75-84** | | | | | **85+** | | | | |
| **NHS** | | | | | | | | | | | | | | | |
| **1-21 Day Risk Window** | | | | | | | | | | | | | | | |
| ***Conditional Poisson Regression*** | | | | | | | | | | | | | | | |
| *Cases in Risk Window* | 393 | | | | | 504 | | | | | 351 | | | | |
| *Cases in Control Window* | 868 | | | | | 1,277 | | | | | 771 | | | | |
| *Risk Days* | 35,095 | | | | | 48,791 | | | | | 31,115 | | | | |
| *Control Days* | 75,863 | | | | | 104,221 | | | | | 63,413 | | | | |
| *Model Results* | RR | 95% CI | | P-Value | SE | RR | 95% CI | | P-Value | SE | RR | 95% CI | | P-Value | SE |
|  | 1.10 | 0.97 | 1.24 | 0.12 | 0.06 | 0.95 | 0.86 | 1.06 | 0.36 | 0.05 | 1.08 | 0.94 | 1.23 | 0.28 | 0.07 |
| ***Attributable Risk*** | | | | | | | | | | | | | | | |
| *# of Eligible Vaccinated* | 3,309,164 | | | | | 2,651,977 | | | | | 933,793 | | | | |
| *Person Years* | 138,682,579 | | | | | 111,076,846 | | | | | 39,000,516 | | | | |
| *Attributable Risk per 100,000 Doses* | AR | 95% CI | | P-Value | SE | AR | 95% CI | | P-Value | SE | AR | 95% CI | | P-Value | SE |
|  | 1.09 | -0.32 | 2.50 | 0.13 | 0.72 | -0.97 | -3.00 | 1.06 | 0.35 | 1.03 | 2.69 | -2.17 | 7.56 | 0.28 | 2.48 |
| *Attributable Risk per 100,000 Person Years* | AR | 95% CI | | P-Value | SE | AR | 95% CI | | P-Value | SE | AR | 95% CI | | P-Value | SE |
|  | 9.52 | -2.76 | 21.80 | 0.13 | 6.27 | -8.46 | -26.13 | 9.22 | 0.35 | 9.02 | 23.54 | -18.94 | 66.03 | 0.28 | 21.68 |
| **22-42 Day Risk Window** | | | | | | | | | | | | | | | |
| ***Conditional Poisson Regression*** | | | | | | | | | | | | | | | |
| *Cases in Risk Window* | 414 | | | | | 551 | | | | | 368 | | | | |
| *Cases in Control Window* | 868 | | | | | 1,277 | | | | | 771 | | | | |
| *Risk Days* | 34,620 | | | | | 48,108 | | | | | 30,185 | | | | |
| *Control Days* | 75,863 | | | | | 104,221 | | | | | 63,413 | | | | |
| *Model Results* | RR | 95% CI | | P-Value | SE | RR | 95% CI | | P-Value | SE | RR | 95% CI | | P-Value | SE |
|  | 1.16 | 1.03 | 1.30 | 0.02 | 0.06 | 1.03 | 0.93 | 1.14 | 0.58 | 0.05 | 1.14 | 1.01 | 1.30 | 0.04 | 0.06 |
| ***Attributable Risk*** | | | | | | | | | | | | | | | |
| *# of Eligible Vaccinated* | 3,309,164 | | | | | 2,651,977 | | | | | 933,793 | | | | |
| *Person Years* | 138,682,579 | | | | | 111,076,846 | | | | | 39,000,516 | | | | |
| *Attributable Risk per 100,000 Doses* | AR | 95% CI | | P-Value | SE | AR | 95% CI | | P-Value | SE | AR | 95% CI | | P-Value | SE |
|  | 1.68 | 0.29 | 3.07 | 0.02 | 0.71 | 0.58 | -1.46 | 2.63 | 0.58 | 1.04 | 4.95 | 0.21 | 9.69 | 0.04 | 2.42 |
| *Attributable Risk per 100,000 Person Years* | AR | 95% CI | | P-Value | SE | AR | 95% CI | | P-Value | SE | AR | 95% CI | | P-Value | SE |
|  | 14.66 | 2.54 | 26.77 | 0.02 | 6.18 | 5.07 | -12.75 | 22.88 | 0.58 | 9.09 | 43.25 | 1.81 | 84.68 | 0.04 | 21.14 |
| **TIA** | | | | | | | | | | | | | | | |
| **1-21 Day Risk Window** | | | | | | | | | | | | | | | |
| ***Conditional Poisson Regression*** | | | | | | | | | | | | | | | |
| *Cases in Risk Window* | 357 | | | | | 474 | | | | | 312 | | | | |
| *Cases in Control Window* | 759 | | | | | 1,230 | | | | | 641 | | | | |
| *Risk Days* | 30,963 | | | | | 45,969 | | | | | 25,336 | | | | |
| *Control Days* | 70,082 | | | | | 103,980 | | | | | 56,985 | | | | |
| *Model Results* | RR | 95% CI | | P-Value | SE | RR | 95% CI | | P-Value | SE | RR | 95% CI | | P-Value | SE |
|  | 1.12 | 0.99 | 1.28 | 0.07 | 0.06 | 0.92 | 0.83 | 1.02 | 0.13 | 0.05 | 1.17 | 1.02 | 1.34 | 0.02 | 0.07 |
| ***Attributable Risk*** | | | | | | | | | | | | | | | |
| *# of Eligible Vaccinated* | 4,393,338 | | | | | 3,427,001 | | | | | 1,203,638 | | | | |
| *Person Years* | 184,138,259 | | | | | 143,554,539 | | | | | 50,276,756 | | | | |
| *Attributable Risk per 100,000 Doses* | AR | 95% CI | | P-Value | SE | AR | 95% CI | | P-Value | SE | AR | 95% CI | | P-Value | SE |
|  | 0.90 | -0.08 | 1.87 | 0.07 | 0.50 | -1.19 | -2.68 | 0.30 | 0.12 | 0.76 | 3.77 | 0.40 | 7.15 | 0.03 | 1.72 |
| *Attributable Risk per 100,000 Person Years* | AR | 95% CI | | P-Value | SE | AR | 95% CI | | P-Value | SE | AR | 95% CI | | P-Value | SE |
|  | 7.82 | -0.68 | 16.33 | 0.07 | 4.34 | -10.39 | -23.38 | 2.59 | 0.12 | 6.63 | 32.95 | 3.46 | 62.44 | 0.03 | 15.05 |
| **22-42 Day Risk Window** | | | | | | | | | | | | | | | |
| ***Conditional Poisson Regression*** | | | | | | | | | | | | | | | |
| *Cases in Risk Window* | 359 | | | | | 485 | | | | | 254 | | | | |
| *Cases in Control Window* | 759 | | | | | 1,230 | | | | | 641 | | | | |
| *Risk Days* | 30,953 | | | | | 45,881 | | | | | 25,289 | | | | |
| *Control Days* | 70,082 | | | | | 103,980 | | | | | 56,985 | | | | |
| *Model Results* | RR | 95% CI | | P-Value | SE | RR | 95% CI | | P-Value | SE | RR | 95% CI | | P-Value | SE |
|  | 1.12 | 0.99 | 1.27 | 0.08 | 0.06 | 0.94 | 0.84 | 1.04 | 0.22 | 0.05 | 0.94 | 0.81 | 1.09 | 0.42 | 0.07 |
| ***Attributable Risk*** | | | | | | | | | | | | | | | |
| *# of Eligible Vaccinated* | 4,393,338 | | | | | 3,427,001 | | | | | 1,203,638 | | | | |
| *Person Years* | 184,138,259 | | | | | 143,554,539 | | | | | 50,276,756 | | | | |
| *Attributable Risk per 100,000 Doses* | AR | 95% CI | | P-Value | SE | AR | 95% CI | | P-Value | SE | AR | 95% CI | | P-Value | SE |
|  | 0.87 | -0.12 | 1.86 | 0.08 | 0.50 | -0.96 | -2.47 | 0.55 | 0.21 | 0.77 | -1.30 | -4.45 | 1.85 | 0.42 | 1.61 |
| *Attributable Risk per 100,000 Person Years* | AR | 95% CI | | P-Value | SE | AR | 95% CI | | P-Value | SE | AR | 95% CI | | P-Value | SE |
|  | 7.60 | -1.02 | 16.21 | 0.08 | 4.40 | -8.40 | -21.55 | 4.75 | 0.21 | 6.71 | -11.35 | -38.88 | 16.18 | 0.42 | 14.04 |
| **NHS/TIA** | | | | | | | | | | | | | | | |
| **1-21 Day Risk Window** | | | | | | | | | | | | | | | |
| ***Conditional Poisson Regression*** | | | | | | | | | | | | | | | |
| *Cases in Risk Window* | 642 | | | | | 860 | | | | | 587 | | | | |
| *Cases in Control Window* | 1,404 | | | | | 2,198 | | | | | 1,242 | | | | |
| *Risk Days* | 57,154 | | | | | 82,916 | | | | | 49,836 | | | | |
| *Control Days* | 125,918 | | | | | 181,480 | | | | | 105,716 | | | | |
| *Model Results* | RR | 95% CI | | P-Value | SE | RR | 95% CI | | P-Value | SE | RR | 95% CI | | P-Value | SE |
|  | 1.10 | 1.00 | 1.21 | 0.05 | 0.05 | 0.94 | 0.87 | 1.02 | 0.12 | 0.04 | 1.12 | 1.01 | 1.24 | 0.03 | 0.05 |
| ***Attributable Risk*** | | | | | | | | | | | | | | | |
| *# of Eligible Vaccinated* | 3,309,164 | | | | | 2,651,977 | | | | | 933,793 | | | | |
| *Person Years* | 138,682,579 | | | | | 111,076,846 | | | | | 39,000,516 | | | | |
| *Attributable Risk per 100,000 Doses* | AR | 95% CI | | P-Value | SE | AR | 95% CI | | P-Value | SE | AR | 95% CI | | P-Value | SE |
|  | 1.79 | 0.02 | 3.56 | 0.05 | 0.90 | -2.13 | -4.76 | 0.50 | 0.11 | 1.34 | 6.57 | 0.49 | 12.66 | 0.03 | 3.11 |
| *Attributable Risk per 100,000 Person Years* | AR | 95% CI | | P-Value | SE | AR | 95% CI | | P-Value | SE | AR | 95% CI | | P-Value | SE |
|  | 15.60 | 0.19 | 31.00 | 0.05 | 7.86 | -18.57 | -41.52 | 4.39 | 0.11 | 11.71 | 57.44 | 4.25 | 110.63 | 0.03 | 27.14 |
| **22-42 Day Risk Window** | | | | | | | | | | | | | | | |
| ***Conditional Poisson Regression*** | | | | | | | | | | | | | | | |
| *Cases in Risk Window* | 680 | | | | | 899 | | | | | 553 | | | | |
| *Cases in Control Window* | 1,404 | | | | | 2,198 | | | | | 1,242 | | | | |
| *Risk Days* | 56,670 | | | | | 82,164 | | | | | 48,874 | | | | |
| *Control Days* | 125,918 | | | | | 181,480 | | | | | 105,716 | | | | |
| *Model Results* | RR | 95% CI | | P-Value | SE | RR | 95% CI | | P-Value | SE | RR | 95% CI | | P-Value | SE |
|  | 1.16 | 1.06 | 1.28 | 0.00 | 0.05 | 0.97 | 0.90 | 1.05 | 0.51 | 0.04 | 1.06 | 0.96 | 1.17 | 0.27 | 0.05 |
| ***Attributable Risk*** | | | | | | | | | | | | | | | |
| *# of Eligible Vaccinated* | 3,309,164 | | | | | 2,651,977 | | | | | 933,793 | | | | |
| *Person Years* | 138,682,579 | | | | | 111,076,846 | | | | | 39,000,516 | | | | |
| *Attributable Risk per 100,000 Doses* | AR | 95% CI | | P-Value | SE | AR | 95% CI | | P-Value | SE | AR | 95% CI | | P-Value | SE |
|  | 2.88 | 1.09 | 4.68 | 0.00 | 0.92 | -0.90 | -3.53 | 1.73 | 0.50 | 1.34 | 3.31 | -2.54 | 9.15 | 0.27 | 2.98 |
| *Attributable Risk per 100,000 Person Years* | AR | 95% CI | | P-Value | SE | AR | 95% CI | | P-Value | SE | AR | 95% CI | | P-Value | SE |
|  | 25.12 | 9.46 | 40.77 | 0.00 | 7.99 | -7.86 | -30.78 | 15.07 | 0.50 | 11.70 | 28.90 | -22.20 | 80.01 | 0.27 | 26.07 |
| **HS** | | | | | | | | | | | | | | | |
| **1-21 Day Risk Window** | | | | | | | | | | | | | | | |
| ***Conditional Poisson Regression*** | | | | | | | | | | | | | | | |
| *Cases in Risk Window* | 101 | | | | | 138 | | | | | 99 | | | | |
| *Cases in Control Window* | 224 | | | | | 360 | | | | | 224 | | | | |
| *Risk Days* | 9,283 | | | | | 13,206 | | | | | 8,384 | | | | |
| *Control Days* | 17,925 | | | | | 24,294 | | | | | 13,885 | | | | |
| *Model Results* | RR | 95% CI | | P-Value | SE | RR | 95% CI | | P-Value | SE | RR | 95% CI | | P-Value | SE |
|  | 1.05 | 0.81 | 1.36 | 0.71 | 0.13 | 0.84 | 0.67 | 1.05 | 0.13 | 0.11 | 1.08 | 0.81 | 1.44 | 0.60 | 0.15 |
| ***Attributable Risk*** | | | | | | | | | | | | | | | |
| *# of Eligible Vaccinated* | 2,764,550 | | | | | 2,234,060 | | | | | 782,854 | | | | |
| *Person Years* | 115,852,292 | | | | | 93,564,395 | | | | | 32,693,025 | | | | |
| *Attributable Risk per 100,000 Doses* | AR | 95% CI | | P-Value | SE | AR | 95% CI | | P-Value | SE | AR | 95% CI | | P-Value | SE |
|  | 0.17 | -0.76 | 1.10 | 0.72 | 0.47 | -1.15 | -2.63 | 0.32 | 0.12 | 0.75 | 0.92 | -2.50 | 4.35 | 0.60 | 1.75 |
| *Attributable Risk per 100,000 Person Years* | AR | 95% CI | | P-Value | SE | AR | 95% CI | | P-Value | SE | AR | 95% CI | | P-Value | SE |
|  | 1.50 | -6.60 | 9.60 | 0.72 | 4.13 | -10.06 | -22.91 | 2.79 | 0.12 | 6.56 | 8.08 | -21.86 | 38.02 | 0.60 | 15.28 |
| **22-42 Day Risk Window** | | | | | | | | | | | | | | | |
| ***Conditional Poisson Regression*** | | | | | | | | | | | | | | | |
| *Cases in Risk Window* | 124 | | | | | 143 | | | | | 85 | | | | |
| *Cases in Control Window* | 224 | | | | | 360 | | | | | 224 | | | | |
| *Risk Days* | 8,837 | | | | | 12,354 | | | | | 7,570 | | | | |
| *Control Days* | 17,925 | | | | | 24,294 | | | | | 13,885 | | | | |
| *Model Results* | RR | 95% CI | | P-Value | SE | RR | 95% CI | | P-Value | SE | RR | 95% CI | | P-Value | SE |
|  | 1.31 | 1.04 | 1.65 | 0.02 | 0.12 | 0.88 | 0.71 | 1.09 | 0.25 | 0.11 | 1.06 | 0.81 | 1.40 | 0.66 | 0.14 |
| ***Attributable Risk*** | | | | | | | | | | | | | | | |
| *# of Eligible Vaccinated* | 2,764,550 | | | | | 2,234,060 | | | | | 782,854 | | | | |
| *Person Years* | 115,852,292 | | | | | 93,564,395 | | | | | 32,693,025 | | | | |
| *Attributable Risk per 100,000 Doses* | AR | 95% CI | | P-Value | SE | AR | 95% CI | | P-Value | SE | AR | 95% CI | | P-Value | SE |
|  | 1.06 | 0.13 | 1.98 | 0.03 | 0.47 | -0.85 | -2.29 | 0.59 | 0.25 | 0.74 | 0.64 | -2.20 | 3.47 | 0.66 | 1.45 |
| *Attributable Risk per 100,000 Person Years* | AR | 95% CI | | P-Value | SE | AR | 95% CI | | P-Value | SE | AR | 95% CI | | P-Value | SE |
|  | 9.19 | 1.10 | 17.28 | 0.03 | 4.13 | -7.41 | -19.97 | 5.16 | 0.25 | 6.41 | 5.57 | -19.22 | 30.37 | 0.66 | 12.65 |

**Abbreviations:** BNT162b2; WT/OMI BA.4/BA.5, Pfizer COVID-19 BivalentmRNA-1273.222, Moderna COVID-19 Bivalent;

HS, hemorrhagic stroke; NHS, non-hemorrhagic stroke; TIA, transient ischemic attacks

**eTable 25. Summary of Relative and Attributable Risk of AESI Following High Dose or Adjuvanted Influenza Vaccination in the Secondary SCCS Analysis Stratified by Concomitant Vaccination Status, Adjusting for Event Dependent Observation Time**

|  | **High Dose or Adjuvanted Influenza Vaccine** | | | | | | | | | |
| --- | --- | --- | --- | --- | --- | --- | --- | --- | --- | --- |
|  | **Concomitant COVAX** | | | | | **No Concomitant COVAX** | | | | |
| **NHS** |  |  |  |  |  |  |  |  |  |  |
| **1-21 Day Risk Window** |  |  |  |  |  |  |  |  |  |  |
| ***Conditional Poisson Regression*** |  |  |  |  |  |  |  |  |  |  |
| *Cases in Risk Window* |  |  | 288 |  |  |  |  | 960 |  |  |
| *Cases in Control Window* |  |  | 649 |  |  |  |  | 2,267 |  |  |
| *Risk Days* |  |  | 26,029 |  |  |  |  | 88,972 |  |  |
| *Control Days* |  |  | 54,786 |  |  |  |  | 188,711 |  |  |
| *Model Results* | RR | 95% CI | | P-Value | SE | RR | 95% CI | | P-Value | SE |
|  | 1.07 | 0.93 | 1.24 | 0.34 | 0.07 | 1.01 | 0.94 | 1.10 | 0.74 | 0.04 |
| ***Attributable Risk*** |  |  |  |  |  |  |  |  |  |  |
| *# of Eligible Vaccinated* |  |  | 1,639,550 |  |  |  |  | 5,255,384 |  |  |
| *Person Years* |  |  | 68,713,401 |  |  |  |  | 220,046,540 |  |  |
| *Attributable Risk per 100,000 Doses* | AR | 95% CI | | P-Value | SE | AR | 95% CI | | P-Value | SE |
|  | 1.19 | -1.25 | 3.63 | 0.34 | 1.24 | 0.24 | -1.16 | 1.65 | 0.74 | 0.72 |
| *Attributable Risk per 100,000 Person Years* | AR | 95% CI | | P-Value | SE | AR | 95% CI | | P-Value | SE |
|  | 10.33 | -10.91 | 31.58 | 0.34 | 10.84 | 2.11 | -10.13 | 14.34 | 0.74 | 6.24 |
| **22-42 Day Risk Window** |  |  |  |  |  |  |  |  |  |  |
| ***Conditional Poisson Regression*** |  |  |  |  |  |  |  |  |  |  |
| *Cases in Risk Window* |  |  | 308 |  |  |  |  | 1,025 |  |  |
| *Cases in Control Window* |  |  | 649 |  |  |  |  | 2,267 |  |  |
| *Risk Days* |  |  | 25,547 |  |  |  |  | 87,366 |  |  |
| *Control Days* |  |  | 54,786 |  |  |  |  | 188,711 |  |  |
| *Model Results* | RR | 95% CI | | P-Value | SE | RR | 95% CI | | P-Value | SE |
|  | 1.14 | 0.99 | 1.31 | 0.06 | 0.07 | 1.08 | 1.00 | 1.17 | 0.04 | 0.04 |
| ***Attributable Risk*** |  |  |  |  |  |  |  |  |  |  |
| *# of Eligible Vaccinated* |  |  | 1,639,550 |  |  |  |  | 5,255,384 |  |  |
| *Person Years* |  |  | 68,713,401 |  |  |  |  | 220,046,540 |  |  |
| *Attributable Risk per 100,000 Doses* | AR | 95% CI | | P-Value | SE | AR | 95% CI | | P-Value | SE |
|  | 2.29 | -0.18 | 4.77 | 0.07 | 1.26 | 1.47 | 0.06 | 2.88 | 0.04 | 0.72 |
| *Attributable Risk per 100,000 Person Years* | AR | 95% CI | | P-Value | SE | AR | 95% CI | | P-Value | SE |
|  | 19.98 | -1.58 | 41.54 | 0.07 | 11.00 | 12.83 | 0.54 | 25.11 | 0.04 | 6.27 |
| **TIA** |  |  |  |  |  |  |  |  |  |  |
| **1-21 Day Risk Window** |  |  |  |  |  |  |  |  |  |  |
| ***Conditional Poisson Regression*** |  |  |  |  |  |  |  |  |  |  |
| *Cases in Risk Window* |  |  | 272 |  |  |  |  | 871 |  |  |
| *Cases in Control Window* |  |  | 562 |  |  |  |  | 2,068 |  |  |
| *Risk Days* |  |  | 22,686 |  |  |  |  | 79,582 |  |  |
| *Control Days* |  |  | 51,298 |  |  |  |  | 179,749 |  |  |
| *Model Results* | RR | 95% CI | | P-Value | SE | RR | 95% CI | | P-Value | SE |
|  | 1.16 | 1.00 | 1.34 | 0.05 | 0.07 | 1.01 | 0.93 | 1.09 | 0.87 | 0.04 |
| ***Attributable Risk*** |  |  |  |  |  |  |  |  |  |  |
| *# of Eligible Vaccinated* |  |  | 2,061,687 |  |  |  |  | 6,962,290 |  |  |
| *Person Years* |  |  | 86,406,033 |  |  |  |  | 291,563,521 |  |  |
| *Attributable Risk per 100,000 Doses* | AR | 95% CI | | P-Value | SE | AR | 95% CI | | P-Value | SE |
|  | 1.80 | -0.04 | 3.64 | 0.06 | 0.94 | 0.08 | -0.90 | 1.07 | 0.87 | 0.50 |
| *Attributable Risk per 100,000 Person Years* | AR | 95% CI | | P-Value | SE | AR | 95% CI | | P-Value | SE |
|  | 15.65 | -0.38 | 31.68 | 0.06 | 8.18 | 0.71 | -7.87 | 9.30 | 0.87 | 4.38 |
| **22-42 Day Risk Window** |  |  |  |  |  |  |  |  |  |  |
| ***Conditional Poisson Regression*** |  |  |  |  |  |  |  |  |  |  |
| *Cases in Risk Window* |  |  | 247 |  |  |  |  | 851 |  |  |
| *Cases in Control Window* |  |  | 562 |  |  |  |  | 2,068 |  |  |
| *Risk Days* |  |  | 22,642 |  |  |  |  | 79,481 |  |  |
| *Control Days* |  |  | 51,298 |  |  |  |  | 179,749 |  |  |
| *Model Results* | RR | 95% CI | | P-Value | SE | RR | 95% CI | | P-Value | SE |
|  | 1.05 | 0.90 | 1.21 | 0.56 | 0.08 | 0.97 | 0.90 | 1.06 | 0.52 | 0.04 |
| ***Attributable Risk*** |  |  |  |  |  |  |  |  |  |  |
| *# of Eligible Vaccinated* |  |  | 2,061,687 |  |  |  |  | 6,962,290 |  |  |
| *Person Years* |  |  | 86,406,033 |  |  |  |  | 291,563,521 |  |  |
| *Attributable Risk per 100,000 Doses* | AR | 95% CI | | P-Value | SE | AR | 95% CI | | P-Value | SE |
|  | 0.52 | -1.26 | 2.31 | 0.56 | 0.91 | -0.32 | -1.31 | 0.66 | 0.52 | 0.50 |
| *Attributable Risk per 100,000 Person Years* | AR | 95% CI | | P-Value | SE | AR | 95% CI | | P-Value | SE |
|  | 4.57 | -10.98 | 20.11 | 0.56 | 7.93 | -2.81 | -11.41 | 5.78 | 0.52 | 4.38 |
| **NHS/TIA** |  |  |  |  |  |  |  |  |  |  |
| **1-21 Day Risk Window** |  |  |  |  |  |  |  |  |  |  |
| ***Conditional Poisson Regression*** |  |  |  |  |  |  |  |  |  |  |
| *Cases in Risk Window* |  |  | 498 |  |  |  |  | 1,591 |  |  |
| *Cases in Control Window* |  |  | 1,074 |  |  |  |  | 3,770 |  |  |
| *Risk Days* |  |  | 43,024 |  |  |  |  | 146,882 |  |  |
| *Control Days* |  |  | 93,333 |  |  |  |  | 319,781 |  |  |
| *Model Results* | RR | 95% CI | | P-Value | SE | RR | 95% CI | | P-Value | SE |
|  | 1.11 | 1.00 | 1.24 | 0.06 | 0.06 | 1.01 | 0.95 | 1.07 | 0.76 | 0.03 |
| ***Attributable Risk*** |  |  |  |  |  |  |  |  |  |  |
| *# of Eligible Vaccinated* |  |  | 1,639,550 |  |  |  |  | 5,255,384 |  |  |
| *Person Years* |  |  | 68,713,401 |  |  |  |  | 220,046,540 |  |  |
| *Attributable Risk per 100,000 Doses* | AR | 95% CI | | P-Value | SE | AR | 95% CI | | P-Value | SE |
|  | 3.05 | -0.13 | 6.23 | 0.06 | 1.62 | 0.28 | -1.51 | 2.07 | 0.76 | 0.91 |
| *Attributable Risk per 100,000 Person Years* | AR | 95% CI | | P-Value | SE | AR | 95% CI | | P-Value | SE |
|  | 26.59 | -1.12 | 54.30 | 0.06 | 14.14 | 2.44 | -13.15 | 18.03 | 0.76 | 7.95 |
| **22-42 Day Risk Window** |  |  |  |  |  |  |  |  |  |  |
| ***Conditional Poisson Regression*** |  |  |  |  |  |  |  |  |  |  |
| *Cases in Risk Window* |  |  | 483 |  |  |  |  | 1,649 |  |  |
| *Cases in Control Window* |  |  | 1,074 |  |  |  |  | 3,770 |  |  |
| *Risk Days* |  |  | 42,513 |  |  |  |  | 145,195 |  |  |
| *Control Days* |  |  | 93,333 |  |  |  |  | 319,781 |  |  |
| *Model Results* | RR | 95% CI | | P-Value | SE | RR | 95% CI | | P-Value | SE |
|  | 1.08 | 0.97 | 1.20 | 0.18 | 0.06 | 1.04 | 0.98 | 1.11 | 0.15 | 0.03 |
| ***Attributable Risk*** |  |  |  |  |  |  |  |  |  |  |
| *# of Eligible Vaccinated* |  |  | 1,639,550 |  |  |  |  | 5,255,384 |  |  |
| *Person Years* |  |  | 68,713,401 |  |  |  |  | 220,046,540 |  |  |
| *Attributable Risk per 100,000 Doses* | AR | 95% CI | | P-Value | SE | AR | 95% CI | | P-Value | SE |
|  | 2.11 | -1.00 | 5.22 | 0.18 | 1.59 | 1.33 | -0.46 | 3.12 | 0.14 | 0.91 |
| *Attributable Risk per 100,000 Person Years* | AR | 95% CI | | P-Value | SE | AR | 95% CI | | P-Value | SE |
|  | 18.35 | -8.74 | 45.43 | 0.18 | 13.82 | 11.59 | -3.97 | 27.16 | 0.14 | 7.94 |
| **HS** |  |  |  |  |  |  |  |  |  |  |
| **1-21 Day Risk Window** |  |  |  |  |  |  |  |  |  |  |
| ***Conditional Poisson Regression*** |  |  |  |  |  |  |  |  |  |  |
| *Cases in Risk Window* |  |  | 68 |  |  |  |  | 270 |  |  |
| *Cases in Control Window* |  |  | 191 |  |  |  |  | 617 |  |  |
| *Risk Days* |  |  | 7,161 |  |  |  |  | 23,712 |  |  |
| *Control Days* |  |  | 13,379 |  |  |  |  | 42,725 |  |  |
| *Model Results* | RR | 95% CI | | P-Value | SE | RR | 95% CI | | P-Value | SE |
|  | 0.88 | 0.64 | 1.20 | 0.42 | 0.16 | 0.99 | 0.84 | 1.17 | 0.92 | 0.08 |
| ***Attributable Risk*** |  |  |  |  |  |  |  |  |  |  |
| *# of Eligible Vaccinated* |  |  | 1,410,945 |  |  |  |  | 4,370,519 |  |  |
| *Person Years* |  |  | 59,131,517 |  |  |  |  | 182,978,195 |  |  |
| *Attributable Risk per 100,000 Doses* | AR | 95% CI | | P-Value | SE | AR | 95% CI | | P-Value | SE |
|  | -0.66 | -2.27 | 0.95 | 0.42 | 0.82 | -0.05 | -1.06 | 0.96 | 0.92 | 0.52 |
| *Attributable Risk per 100,000 Person Years* | AR | 95% CI | | P-Value | SE | AR | 95% CI | | P-Value | SE |
|  | -5.74 | -19.73 | 8.25 | 0.42 | 7.14 | -0.45 | -9.27 | 8.38 | 0.92 | 4.50 |
| **22-42 Day Risk Window** |  |  |  |  |  |  |  |  |  |  |
| ***Conditional Poisson Regression*** |  |  |  |  |  |  |  |  |  |  |
| *Cases in Risk Window* |  |  | 87 |  |  |  |  | 265 |  |  |
| *Cases in Control Window* |  |  | 191 |  |  |  |  | 617 |  |  |
| *Risk Days* |  |  | 6,792 |  |  |  |  | 21,969 |  |  |
| *Control Days* |  |  | 13,379 |  |  |  |  | 42,725 |  |  |
| *Model Results* | RR | 95% CI | | P-Value | SE | RR | 95% CI | | P-Value | SE |
|  | 1.12 | 0.85 | 1.47 | 0.41 | 0.14 | 1.04 | 0.89 | 1.21 | 0.64 | 0.08 |
| ***Attributable Risk*** |  |  |  |  |  |  |  |  |  |  |
| *# of Eligible Vaccinated* |  |  | 1,410,945 |  |  |  |  | 4,370,519 |  |  |
| *Person Years* |  |  | 59,131,517 |  |  |  |  | 182,978,195 |  |  |
| *Attributable Risk per 100,000 Doses* | AR | 95% CI | | P-Value | SE | AR | 95% CI | | P-Value | SE |
|  | 0.67 | -0.93 | 2.26 | 0.41 | 0.81 | 0.22 | -0.70 | 1.14 | 0.64 | 0.47 |
| *Attributable Risk per 100,000 Person Years* | AR | 95% CI | | P-Value | SE | AR | 95% CI | | P-Value | SE |
|  | 5.79 | -8.08 | 19.66 | 0.41 | 7.08 | 1.92 | -6.10 | 9.94 | 0.64 | 4.09 |

**Abbreviations:** BNT162b2; WT/OMI BA.4/BA.5, Pfizer COVID-19 BivalentmRNA-1273.222, Moderna COVID-19 Bivalent;

HS, hemorrhagic stroke; NHS, non-hemorrhagic stroke; TIA, transient ischemic attacks

**eFigure 1. Monthly Incidence Rates of NHS, TIA, NHS/TIA, and HS per 100,000 Person-Years at Risk**

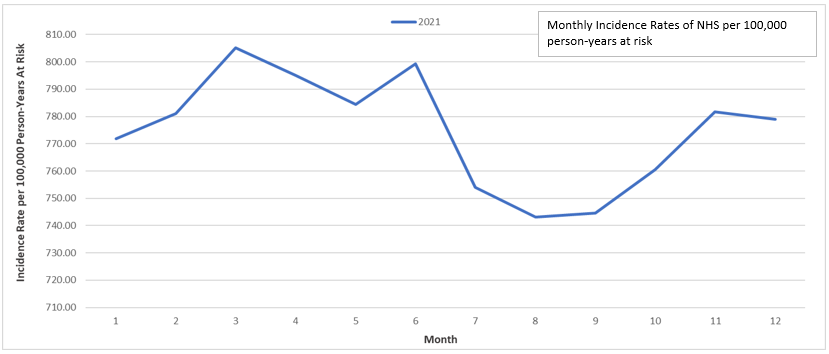

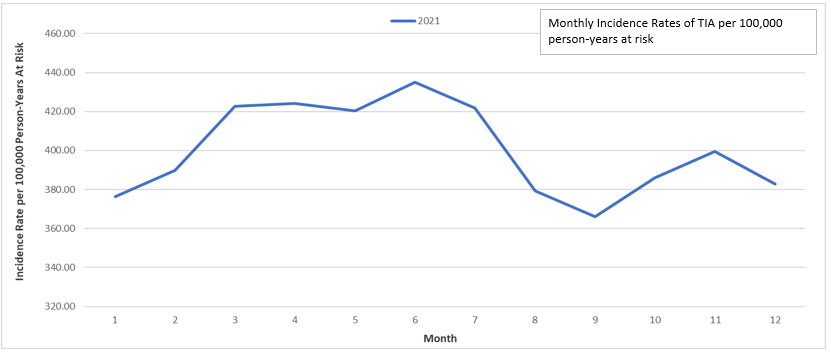

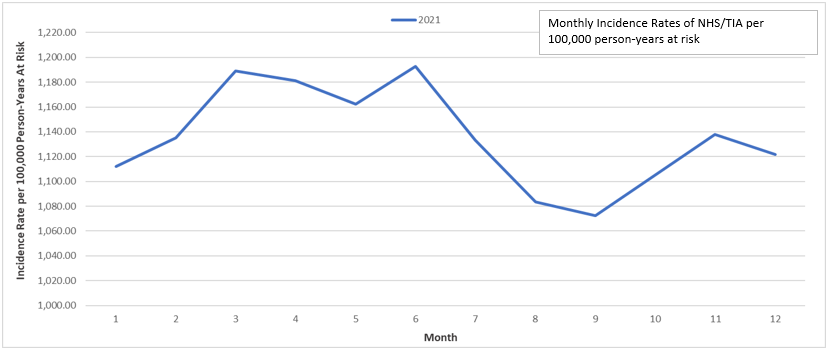

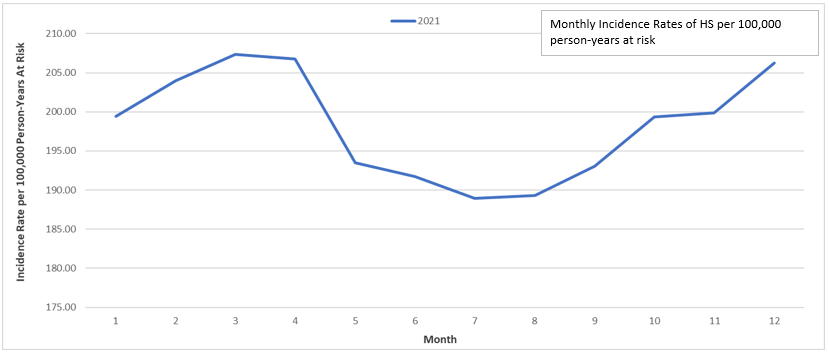

**Abbreviations**: HS, hemorrhagic stroke; NHS, non-hemorrhagic stroke; TIA, transient ischemic attacks

**eFigure 2. Temporal Scan Plots**

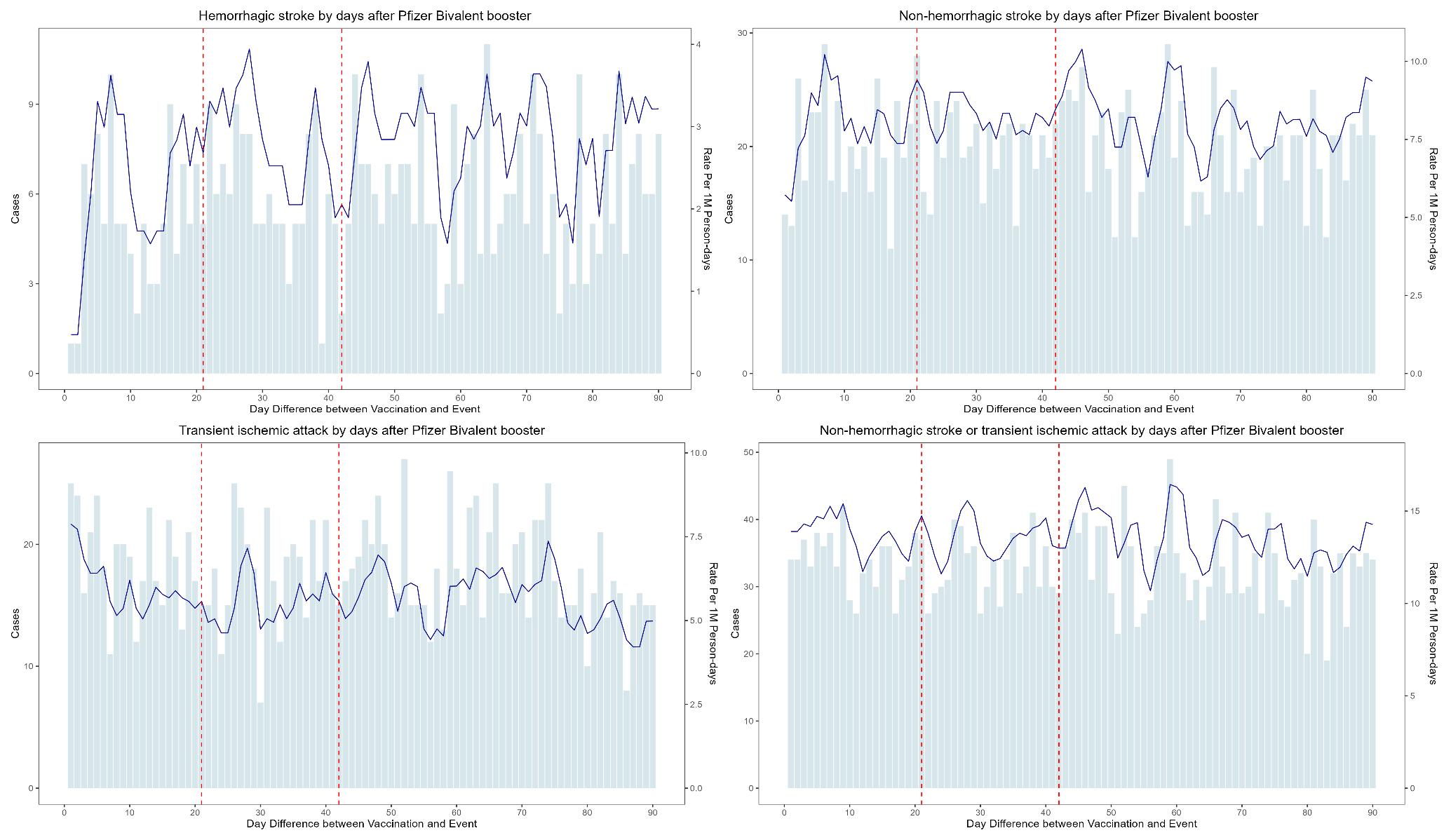

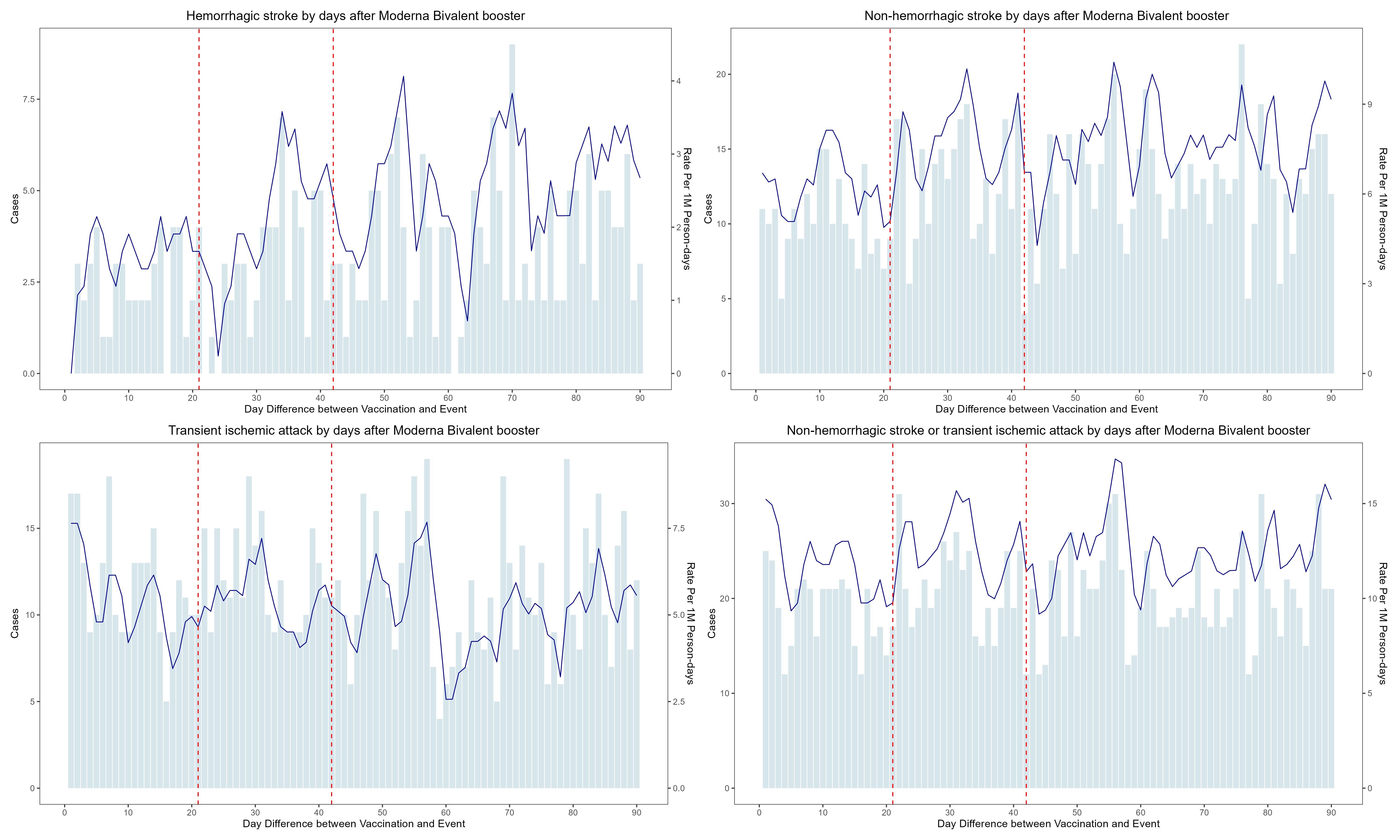

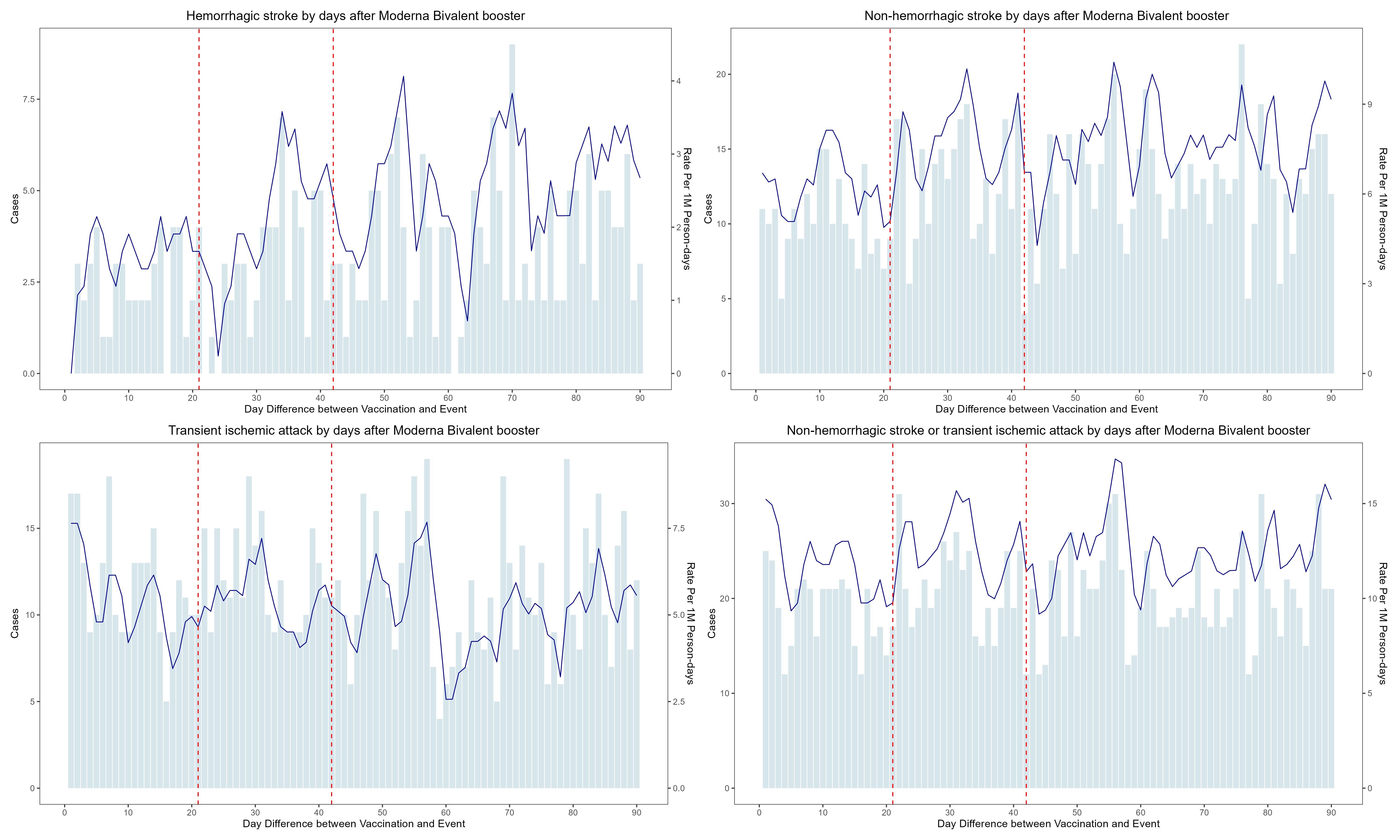
